# Sodium-glucose cotransporter 2 inhibitors versus dipeptidyl peptidase 4 inhibitors on new-onset overall cancer in type 2 diabetes mellitus: a population-based study

**DOI:** 10.1101/2022.07.21.22277915

**Authors:** Cheuk To Chung, Ishan Lakhani, Oscar Hou-In Chou, Teddy Tai Loy Lee, Christopher Dee, Kendrick Ng, Wing Tak Wong, Tong Liu, Sharen Lee, Qingpeng Zhang, Bernard Man Yung Cheung, Gary Tse, Jiandong Zhou

**Affiliations:** Diabetes Research Unit, Cardiovascular Analytics Group, Hong Kong, China-UK Collaboration; Division of Clinical Pharmacology and Therapeutics, Department of Medicine, LKS Faculty of Medicine, The University of Hong Kong, Hong Kong, China; Department of Radiation Oncology, Memorial Sloan Kettering Cancer Center, New York, NY, USA; Department of Medical Oncology, University College London Hospitals NHS Foundation Trust, London, United Kingdom; School of Life Sciences, Chinese University of Hong Kong, Hong Kong, China; Tianjin Key Laboratory of Ionic-Molecular Function of Cardiovascular Disease, Department of Cardiology, Tianjin Institute of Cardiology, Second Hospital of Tianjin Medical University, Tianjin 300211, China; School of Data Science, City University of Hong Kong, Hong Kong, China; Kent and Medway Medical School, Canterbury, Kent, CT2 7NT, United Kingdom; Nuffield Department of Medicine, University of Oxford, Oxford, United Kingdom

## Abstract

**Background:** There is much uncertainty regarding the comparative risks of cancer for T2DM patients on SGLT2I versus DPP4I.

**Methods:** This population-based cohort study patients included T2DM patients who were administered with either SGLT2I or DPP4I between January 1st, 2015, to December 31st, 2020 in Hong Kong.

**Results:** Amongst 60112 T2DM patients (mean baseline age: 62.1±12.4 years, male: 56.36%), 18167 patients were SGLT2I users and 41945 patients were DPP4I users. Multivariate cox regression analysis revealed that SGLT2I usage was associated with a decreased risk of all-cause mortality (HR:0.92; 95%CI:0.84-0.99; P=0.04), cancer-related mortality (HR:0.58; 95%CI:0.42-0.80; P≤0.001) and a 30% risk reduction of new-onset overall cancer (HR:0.70; 95%CI:0.59-0.84; P≤0.001). Dapagliflozin and ertugliflozin both demonstrated superiority in relation to new-onset cancer development, with the former demonstrating a lowered risk of breast cancer (HR:0.48; 95%CI:0.27-0.83; P=0.001).

**Conclusion:** SGLT2I was associated with lower risk of all-cause mortality, cancer-related mortality and new-onset overall cancer compared to DPP4I.

## Introduction

The burden of cancer incidence has drastically increased over the years and is currently the second leading cause of death globally. In 2020, the Global Cancer Observatory estimated a total of 19.3 million new cancer cases and 10 million cancer deaths (1). Despite efforts to advance preventive interventions, the asymptomatic nature of the disease during its early stages poses a challenge for cancer diagnosis (2, 3). Although the aetiology of some cancer types still requires further exploration, currently established risk factors include but are not limited to type 2 diabetes mellitus (T2DM), hypertension and smoking (4). Numerous epidemiological studies have found supporting evidence for the association between T2DM and many different types of cancer, such as liver cancer, breast cancer and colorectal cancer (5, 6). As such, this has instigated growing interest into anti-diabetic medications as a potential adjuvant in the clinical management of cancer.

Metformin, in multiple pre-clinical studies, has been described to be useful in the treatment of various types of malignancies (7-9). However, current evidence presents conflicting results regarding the use of novel anti-diabetic agents such as sodium-glucose cotransporter 2 inhibitors (SGLT2I) and dipeptidyl peptidase 4 inhibitors (DPP4I). A systematic review and meta-analysis revealed canagliflozin had protective effects against gastrointestinal cancers, while empagliflozin was found to have increased risks of bladder cancer (10). Similarly, previous studies have reported increased risks of liver, kidney and bladder cancer and melanoma in T2DM patients using DPP4I (11). In stark contrast, there is also evidence to suggest an absence of any association between these medications and malignancy, even when stratified by different subtypes of DPP4I (12). Regarding SGLT2I, a retrospective study from Taiwan found SGLT2I usage was associated with lower risks of cancer-related mortality relative to DPP4I (13). Likewise, another investigation comparing the risk of urinary tract and haematological malignancies amongst SGLT2I and DPP4I users demonstrated superiority of the former (14).

Despite the aforementioned findings, there is still much uncertainty regarding the comparative associations between SGLT2I and DPP4I with different types of new-onset overall cancer. Given the prevalence with which these medications are used, the present study aims to assess the effects of SGLT2I versus DPP4I on the risk of new-onset overall cancer and pre-specified cancers in T2DM patients from Hong Kong

## Method

### Study population

This population-based, retrospective study has assessed integrated medical records of patients through the Clinical Data Analysis and Reporting System (CDARS), including disease diagnosis, laboratory results, past comorbidities, medication prescription details and clinical characteristics. The system has also been used by our team in previous epidemiological research in Hong Kong (15-17). Patients who were diagnosed with T2DM and were administered either SGLT2 or DPP4 inhibitors, between January 1st, 2015, to December 31st, 2020 in centres under the Hong Kong Hospital Authority were included in the study cohort. The exclusion criteria for the cohort were as follows: (1) Patients who died within 30 days after initial drug exposure; (2) Patients under 18 years old; (3) Patients with prior all-cause malignancies; (4) Patients with new-onset all-cause malignancies development less than 1 year after drug exposure and (5) Patients with both DPP4I and SGLT2I prescription. The study has also received clearance from The Joint Chinese University of Hong Kong-New Territories East Cluster Clinical Research Ethics Committee.

### Clinical and Biochemical Data Collection

Biochemical and clinical data were extracted for this cohort. Patient’s demographic information includes sex, baseline age and date of initial drug use. Past comorbidities include diabetes mellitus disease duration, hyperlipidemia, obesity, hypertension, alcoholism, liver diseases, autoimmune diseases, HIV, carcinogen pathogens, previous irradiation, COPD, gastrointestinal diseases, cardiovascular diseases, ischemic stroke, diabetic eye diseases and renal diseases. Moreover, Charlson’s standard comorbidity index was also calculated. Renal function was represented by using the abbreviated modification of diet in renal disease (MDRD) formula (PMID: 23181772).

Moreover, anti-diabetic and non-SGLT2I/DPP4 medications and baseline laboratory data results were also extracted. Data on the following medications were extracted: sulphonylurea, insulin, metformin, thiazolidinedione, acarbose, glucagon-like peptide-1 receptor agonists, statins and fibrates, ACEI, ARB, anti-depressant drugs, antihypertensive drugs, anti-hepatitic drugs, anticoagulants, diuretics, nitrates, beta-blockers, calcium channel blockers and non-steroidal anti-inflammatory drugs. The extracted laboratory data include lipid profiles, complete blood count, renal function test, biochemical test and glycaemic profiles.

### Outcome and Statistical analysis

The primary outcome of this study was new-onset all-cause cancer incidence, all-cause cancer-related mortality and all-cause mortality. Mortality data were extracted from the Hong Kong Death Registry, an official government registry linked with CDARS that registers death records of all Hong Kong citizens. Study outcomes and comorbidities were documented using the ICD-9 codes, whilst mortality outcomes were recorded using the ICD-10 coding system. ICD-10 codes C00-C97 were used to identify all-cause cancer mortality. The ICD-9 and ICD-10 codes are summarised in **Supplementary Table 1**.

Descriptive statistics were used to summarise baseline characteristics for this cohort. Mean and standard deviation (SD) was used to represent continuous variables, while a number and percentage were used to represent categorical variables. Propensity score matching with a 1:1 ratio between SGLT2I and DPP4I users and patients with and without new-onset overall cancer risk based on demographics, prior comorbidities, laboratory data, medication usage, Charlson comorbidity index and abbreviated MDRD were performed using the nearest neighbour strategy with the calliper as 0.1. Univariate and multivariate Cox proportional regression was performed for both before and after matching to identify significant predictors of new-onset all-cause cancer occurrence and mortality. This is further corroborated by the inverse probability of treatment-weighting using propensity scores and calculating incidence rate ratios. Cumulative incidence curves were also calculated to visually depict the difference in the time-to-adverse event by comparing the SGLT2I and DPP4I groups. Values with a p-value of less than 0.05 were considered statistically significant. Statistical analyses and propensity score matching was performed with RStudio software (version: 1.1.456) and Stata software (version 13.0), respectively.

## Results

### Baseline characteristics

This study included 60112 T2DM patients (mean baseline age: 62.1 ± 12.4 years, male: 56.36%, mean diabetes mellitus disease duration to baseline date: 640.6 ± 1264.0 days), of which 18167 patients were SGLT2I users and 41945 patients were DPP4I users. In the SGLT2I subgroup, the corresponding number of patients on individual SGLT2Is is as follows: 4523 (24.89%) on canagliflozin, 10556 (58.10%) on dapagliflozin, 3780 (20.80%) on empagliflozin and 2527 (13.90%) on ertugliflozin. During the follow-up period, 1533 patients developed new-onset overall cancer, 3033 patients died from any cause, of which 506 patients died due to cancer-related causes. Data on specific types of new-onset overall cancers were also extracted: 249 patients developed new-onset lung cancer, 817 patients developed new-onset gastrointestinal cancer, 201 patients developed new-onset breast cancer, 261 patients developed new-onset genitourinary cancer and 97 patients developed new-onset bladder cancer (**Figure 1**). The baseline characteristics for continuous and discrete variables of demographics, laboratory and medication histories for patients before and after matching are shown in **Table 1, Supplementary Table 3A, Supplementary Table 3B and Supplementary Table 3C**. The method of variability (standard deviation) calculation is shown in **Supplementary Table 2**.

**Figure 1.**
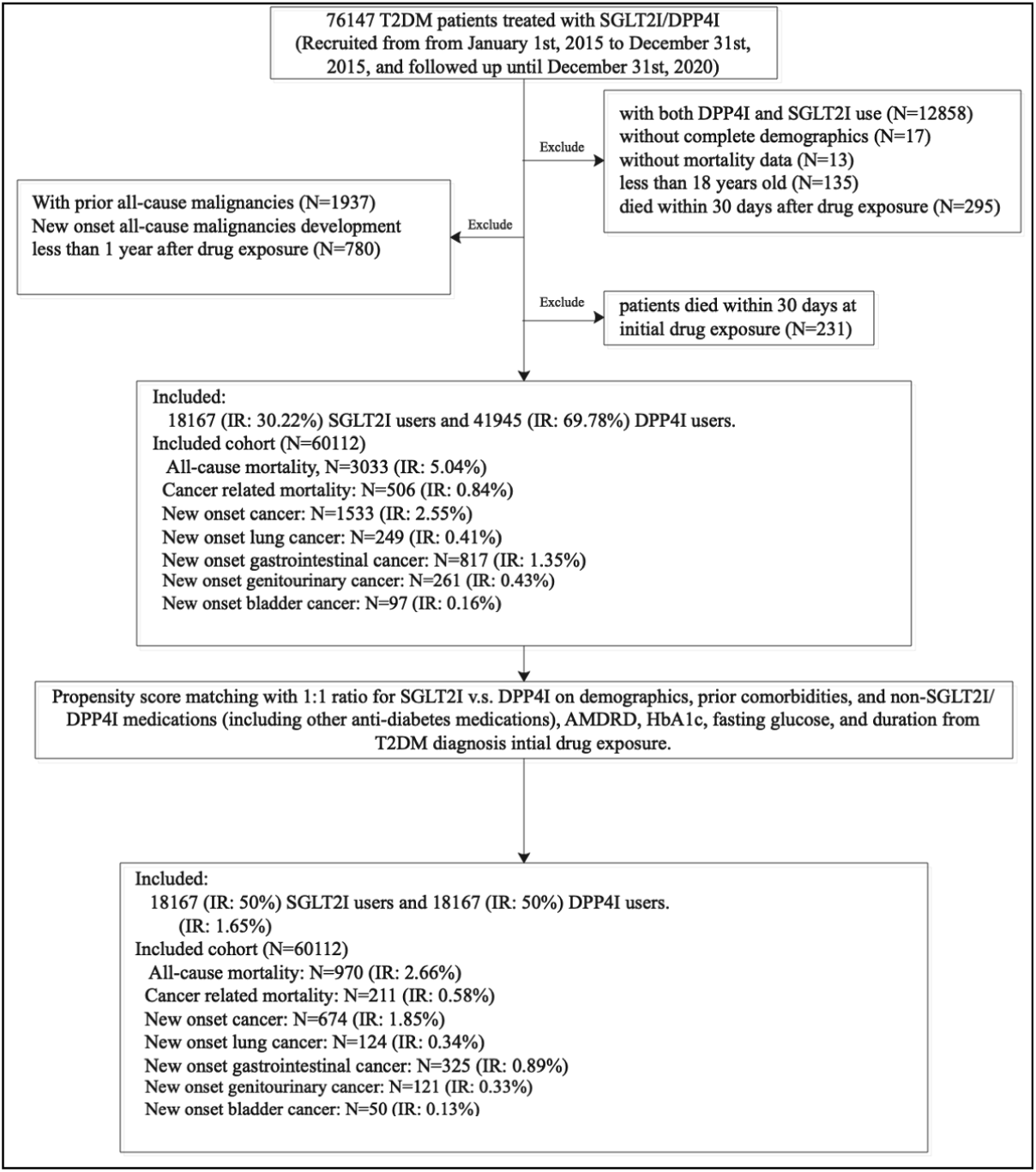
Procedures of data processing for the study cohort. IR: Incidence rate; SGLT2I: Sodium-glucose cotransporter-2 inhibitors; DPP4I: Dipeptidyl peptidase-4 inhibitors.

**Table 1.** Baseline and clinical characteristics of patients with SGLT2I v.s. DPP4I use before and after propensity score matching (1:1). * for SMD≥0.2; SD: standard deviation; SGLT2I: sodium glucose cotransporter-2 inhibitor; DPP4I: dipeptidyl peptidase-4 inhibitor; SD: standard deviation; SGLT2I: sodium glucose cotransporter-2 inhibitor; DPP4I: dipeptidyl peptidase-4 inhibitor; MDRD: modification of diet in renal disease; # indicated the difference between SGLT2I users and DPP4I users.

| Characteristics | Before matching |  |  |  | After matching |  |  |  |
| --- | --- | --- | --- | --- | --- | --- | --- | --- |
|  | All (N=60112)<br>Mean(SD);N or<br>Count(%) | SGLT2I v.s. DPP4I<br>(N=18167)<br>Mean(SD);N or<br>Count(%) | No SGLT2I v.s.<br>DPP4I (N=41945)<br>Mean(SD);N or<br>Count(%) | SMD | All (N=36334)<br>Mean(SD);N or<br>Count(%) | SGLT2I v.s. DPP4I<br>(N=18167)<br>Mean(SD);N or<br>Count(%) | No SGLT2I v.s.<br>DPP4I (N=18167)<br>Mean(SD);N or<br>Count(%) | SMD |
| Demographics |  |  |  |  |  |  |  |  |
| Male gender | 33883(56.36%) | 11138(61.30%) | 22745(54.22%) | 0.14 | 22288(61.34%) | 11147(61.35%) | 11141(61.32%) | <0.01 |
| Female gender | 26229(43.63%) | 7029(38.69%) | 19200(45.77%) | 0.14 | 14046(38.65%) | 7020(38.64%) | 7026(38.67%) | <0.01 |
| Baseline age, years | 62.1(12.4);n=60112 | 57.8(11.2);n=18167 | 63.9(12.5);n=41945 | 0.51* | 58.2(11.0);n=36334 | 57.8(11.2);n=18167 | 58.7(10.9);n=18167 | 0.08 |
| 18-50 | 9057(15.06%) | 3878(21.34%) | 5179(12.34%) | 0.24* | 7212(19.84%) | 3888(21.40%) | 3324(18.29%) | 0.08 |
| 50-60 | 17550(29.19%) | 6524(35.91%) | 11026(26.28%) | 0.21* | 13285(36.56%) | 6543(36.01%) | 6742(37.11%) | 0.02 |
| 60-70 | 18029(29.99%) | 5464(30.07%) | 12565(29.95%) | <0.01 | 11194(30.80%) | 5457(30.03%) | 5737(31.57%) | 0.03 |
| 70-80 | 10338(17.19%) | 1883(10.36%) | 8455(20.15%) | 0.27* | 3766(10.36%) | 1870(10.29%) | 1896(10.43%) | <0.01 |
| >80 | 5145(8.55%) | 421(2.31%) | 4724(11.26%) | 0.36* | 881(2.42%) | 412(2.26%) | 469(2.58%) | 0.02 |
| Past comorbidities |  |  |  |  |  |  |  |  |
| Charlson standard comorbidity index | 1.9(1.4);n=60112 | 1.5(1.2);n=18167 | 2.1(1.4);n=41945 | 0.45* | 1.5(1.2);n=36334 | 1.52(1.17);n=18167 | 1.55(1.14);n=18167 | 0.02 |
| Duration from earliest diabetes mellitus date to baseline date, day | 619.9(1341.9);n=60112 | 616.9(1352.3);n=18167 | 621.2(1337.4);n=41945 | <0.01 | 590.9(1323.1);n=36334 | 618.7(1354.2);n=18167 | 563.1(1290.6);n=18167 | 0.04 |
| Hypertension | 13376(22.25%) | 4284(23.58%) | 9092(21.67%) | 0.05 | 8325(22.91%) | 4283(23.57%) | 4042(22.24%) | 0.03 |
| Hyperlipidaemia | 1620(2.69%) | 662(3.64%) | 958(2.28%) | 0.08 | 1265(3.48%) | 659(3.62%) | 606(3.33%) | 0.02 |
| Hypotension | 350(0.58%) | 76(0.41%) | 274(0.65%) | 0.03 | 152(0.41%) | 75(0.41%) | 77(0.42%) | <0.01 |
| Overweight, obesity and hyperalimentation | 432(0.71%) | 297(1.63%) | 135(0.32%) | 0.13 | 576(1.58%) | 300(1.65%) | 276(1.51%) | 0.01 |
| Gout | 1510(2.51%) | 394(2.16%) | 1116(2.66%) | 0.03 | 782(2.15%) | 394(2.16%) | 388(2.13%) | <0.01 |
| Heart failure | 1629(2.70%) | 447(2.46%) | 1182(2.81%) | 0.02 | 874(2.40%) | 447(2.46%) | 427(2.35%) | 0.01 |
| Acute myocardial infarction | 1521(2.53%) | 616(3.39%) | 905(2.15%) | 0.08 | 1210(3.33%) | 613(3.37%) | 597(3.28%) | <0.01 |
| Ischemic heart disease | 5680(9.44%) | 2342(12.89%) | 3338(7.95%) | 0.16 | 4466(12.29%) | 2333(12.84%) | 2133(11.74%) | 0.03 |
| Peripheral vascular disease | 375(0.62%) | 97(0.53%) | 278(0.66%) | 0.02 | 194(0.53%) | 99(0.54%) | 95(0.52%) | <0.01 |
| Stroke/transient ischemic attack | 1777(2.95%) | 474(2.60%) | 1303(3.10%) | 0.03 | 940(2.58%) | 477(2.62%) | 463(2.54%) | <0.01 |
| Atrial fibrillation | 1325(2.20%) | 388(2.13%) | 937(2.23%) | 0.01 | 766(2.10%) | 386(2.12%) | 380(2.09%) | <0.01 |
| Diabetic eye disease | 4045(6.72%) | 1322(7.27%) | 2723(6.49%) | 0.03 | 2459(6.76%) | 1321(7.27%) | 1138(6.26%) | 0.04 |
| Alcohol dependence | 119(0.19%) | 22(0.12%) | 97(0.23%) | 0.03 | 44(0.12%) | 23(0.12%) | 21(0.11%) | <0.01 |
| Chronic liver disease and cirrhosis | 1195(1.98%) | 513(2.82%) | 682(1.62%) | 0.08 | 995(2.73%) | 515(2.83%) | 480(2.64%) | 0.01 |
| Viral hepatitis | 629(1.04%) | 209(1.15%) | 420(1.00%) | 0.01 | 414(1.13%) | 211(1.16%) | 203(1.11%) | <0.01 |
| History of acute liver injury | 159(0.26%) | 36(0.19%) | 123(0.29%) | 0.02 | 72(0.19%) | 36(0.19%) | 36(0.19%) | <0.01 |
| Other liver disease | 612(1.01%) | 163(0.89%) | 449(1.07%) | 0.02 | 324(0.89%) | 160(0.88%) | 164(0.90%) | <0.01 |
| Autoimmune disease tissue | 620(1.03%) | 198(1.08%) | 422(1.00%) | 0.01 | 386(1.06%) | 200(1.10%) | 186(1.02%) | 0.01 |
| Other carcinogens pathogen | 13(0.02%) | 6(0.03%) | 7(0.01%) | 0.01 | 12(0.03%) | 6(0.03%) | 6(0.03%) | <0.01 |
| Chronic obstructive pulmonary disease | 69(0.11%) | 12(0.06%) | 57(0.13%) | 0.02 | 24(0.06%) | 12(0.06%) | 12(0.06%) | <0.01 |
| Gastrointestinal disease | 1353(2.25%) | 349(1.92%) | 1004(2.39%) | 0.03 | 690(1.89%) | 350(1.92%) | 340(1.87%) | <0.01 |
| Medications |  |  |  |  |  |  |  |  |
| SGLT2I v.s. DPP4I | 18167(30.22%) | 18167(100.00%) | 0(0.00%) | inf* | 18167(50.00%) | 18167(100.00%) | 0(0.00%) | inf* |
| SGLT2I frequency | 7.4(10.1);n=18167 | 7.4(10.1);n=18167 | - | - | 7.3(10.0);n=18167 | 7.3(10.0);n=18167 | - | - |
| DPP4I frequency | 5.5(7.2);n=41945 | - | 5.5(7.2);n=41945 | - | 6.3(8.0);n=18167 | - | 6.3(8.0);n=18167 | - |
| SGLT2I duration, days | 538.1(674.5);n=18167 | 538.1(674.5);n=18167 | - | - | 538.5(673.6);n=18167 | 538.5(673.6);n=18167 | - | - |
| DPP4I duration, days | 528.9(301.0);n=41945 | - | 528.9(301.0);n=41945 | - | 575.3(360.3);n=18167 | - | 575.3(360.3);n=18167 | - |
| Metformin | 54407(90.50%) | 16917(93.11%) | 37490(89.37%) | 0.13 | 33945(93.42%) | 16923(93.15%) | 17022(93.69%) | 0.02 |
| Sulphonylurea | 46521(77.39%) | 12899(71.00%) | 33622(80.15%) | 0.21* | 26376(72.59%) | 12889(70.94%) | 13487(74.23%) | 0.07 |
| Insulin | 29349(48.82%) | 9517(52.38%) | 19832(47.28%) | 0.1 | 19074(52.49%) | 9509(52.34%) | 9565(52.65%) | 0.01 |
| Acarbose | 1566(2.60%) | 751(4.13%) | 815(1.94%) | 0.13 | 1429(3.93%) | 744(4.09%) | 685(3.77%) | 0.02 |
| Thiozolidinedone | 12046(20.03%) | 5130(28.23%) | 6916(16.48%) | 0.28* | 9569(26.33%) | 5134(28.26%) | 4435(24.41%) | 0.09 |
| Glucagon-like peptide-1 receptor agonists | 2044(3.40%) | 1505(8.28%) | 539(1.28%) | 0.33* | 2797(7.69%) | 1506(8.28%) | 1291(7.10%) | 0.04 |
| ACEI/ARB | 18249(30.35%) | 11149(61.36%) | 7100(16.92%) | 1.02* | 22352(61.51%) | 11152(61.38%) | 11200(61.65%) | 0.01 |
| Antidepressants | 2742(4.56%) | 1665(9.16%) | 1077(2.56%) | 0.28* | 2971(8.17%) | 1661(9.14%) | 1310(7.21%) | 0.07 |
| Antihypertensive drugs | 2311(3.84%) | 1681(9.25%) | 630(1.50%) | 0.35* | 3051(8.39%) | 1680(9.24%) | 1371(7.54%) | 0.06 |
| Antihepatitis | 809(1.34%) | 336(1.84%) | 473(1.12%) | 0.06 | 664(1.82%) | 337(1.85%) | 327(1.79%) | <0.01 |
| Anticoagulants | 29401(48.91%) | 18166(99.99%) | 11235(26.78%) | 2.34* | 36332(99.99%) | 18166(99.99%) | 18166(99.99%) | <0.01 |
| Antiplatelets | 10115(16.82%) | 5981(32.92%) | 4134(9.85%) | 0.59* | 11686(32.16%) | 5975(32.88%) | 5711(31.43%) | 0.03 |
| Statins and fibrates | 34307(57.07%) | 13773(75.81%) | 20534(48.95%) | 0.58* | 27749(76.37%) | 13780(75.85%) | 13969(76.89%) | 0.02 |
| Nitrates | 4652(7.73%) | 2688(14.79%) | 1964(4.68%) | 0.35* | 5278(14.52%) | 2684(14.77%) | 2594(14.27%) | 0.01 |
| Non-steroidal anti-inflammatory drugs | 9719(16.16%) | 5734(31.56%) | 3985(9.50%) | 0.57* | 11369(31.29%) | 5730(31.54%) | 5639(31.03%) | 0.01 |
| Diuretics | 10179(16.93%) | 5633(31.00%) | 4546(10.83%) | 0.51* | 11095(30.53%) | 5628(30.97%) | 5467(30.09%) | 0.02 |
| Beta-blockers | 8075(13.43%) | 4696(25.84%) | 3379(8.05%) | 0.49* | 9261(25.48%) | 4694(25.83%) | 4567(25.13%) | 0.02 |
| Calcium channel blockers | 14154(23.54%) | 8107(44.62%) | 6047(14.41%) | 0.70* | 16462(45.30%) | 8102(44.59%) | 8360(46.01%) | 0.03 |
| Subclinical biomarkers |  |  |  |  |  |  |  |  |
| Abbreviated MDRD, mL/min/1.73m<sup>2</sup> | 81.4(27.9);n=49633 | 90.1(23.8);n=15520 | 77.4(28.8);n=34113 | 0.48* | 88.6(23.5);n=29687 | 90.1(23.8);n=15514 | 86.9(23.1);n=14173 | 0.14 |
| Most severe renal damage (<15 mL/min/1.73m<sup>2</sup>) | 436.0(0.72%) | 16.0(0.08%) | 420.0(1.00%) | 0.12 | 40.0(0.11%) | 16.0(0.08%) | 24.0(0.13%) | 0.01 |
| Severe renal damage ([15, 30) mL/min/1.73m<sup>2</sup>) | 1108.0(1.84%) | 41.0(0.22%) | 1067.0(2.54%) | 0.2 | 111.0(0.30%) | 39.0(0.21%) | 72.0(0.39%) | 0.03 |
| Moderate to severe renal damage ([30, 45) mL/min/1.73m<sup>2</sup>) | 3548.0(5.90%) | 265.0(1.45%) | 3283.0(7.82%) | 0.31* | 657.0(1.80%) | 261.0(1.43%) | 396.0(2.17%) | 0.06 |
| Mild to moderate renal damage ([45, 60) mL/min/1.73m<sup>2</sup>) | 5816.0(9.67%) | 999.0(5.49%) | 4817.0(11.48%) | 0.22* | 2222.0(6.11%) | 1000.0(5.50%) | 1222.0(6.72%) | 0.05 |
| Mild renal damage ([60, 90] mL/min/1.73m<sup>2</sup>) | 19920.0(33.13%) | 6788.0(37.36%) | 13132.0(31.30%) | 0.13 | 12997.0(35.77%) | 6783.0(37.33%) | 6214.0(34.20%) | 0.07 |
| Chronic kidney disease (>90 mL/min/1.73m<sup>2</sup>) | 18805.0(31.28%) | 7411.0(40.79%) | 11394.0(27.16%) | 0.29* | 13660.0(37.59%) | 7415.0(40.81%) | 6245.0(34.37%) | 0.13 |
| Neutrophil-to-lymphocyte ratio | 3.4(4.5);n=23978 | 3.0(3.9);n=8133 | 3.6(4.8);n=15845 | 0.15 | 3.1(3.8);n=15970 | 3.0(3.9);n=8146 | 3.2(3.7);n=7824 | 0.05 |
| Platelet-to-lymphocyte ratio | 142.0(148.2);n=23976 | 133.5(168.7);n=8131 | 146.4(136.3);n=15845 | 0.08 | 133.6(137.8);n=15969 | 133.3(168.5);n=8144 | 133.9(95.8);n=7825 | <0.01 |
| Neutrophil-to-high-density lipoprotein ratio | 0.3(0.2);n=21968 | 0.27(0.16);n=7755 | 0.27(0.2);n=14213 | 0.01 | 0.3(0.2);n=15080 | 0.27(0.16);n=7770 | 0.27(0.16);n=7310 | 0.01 |
| Low density lipoprotein ratio-to-high density lipoprotein ratio | 2.1(0.9);n=46116 | 2.14(0.83);n=14654 | 2.1(0.86);n=31462 | 0.05 | 2.1(0.8);n=27933 | 2.14(0.83);n=14653 | 2.13(0.84);n=13280 | 0.01 |
| Triglyceride-glucose index | 7.6(0.7);n=42025 | 7.6(0.7);n=13571 | 7.5(0.7);n=28454 | 0.15 | 7.6(0.7);n=25514 | 7.64(0.72);n=13576 | 7.64(0.73);n=11938 | 0.01 |
| Protein-to-creatinine ratio | 3.0(1.5);n=29872 | 3.4(1.5);n=10496 | 2.8(1.5);n=19376 | 0.34* | 3.3(1.5);n=19746 | 3.4(1.5);n=10502 | 3.2(1.4);n=9244 | 0.13 |
| Aspartate aminotransferase-to-alanine transaminase ratio | 1.1(3.2);n=8773 | 0.9(1.0);n=3284 | 1.2(3.9);n=5489 | 0.08 | 0.9(0.8);n=6377 | 0.92(0.99);n=3279 | 0.94(0.5);n=3098 | 0.03 |
| Complete blood counts, renal and liver functions |  |  |  |  |  |  |  |  |
| Mean corpuscular volume, fL | 87.1(7.5);n=29998 | 86.6(7.2);n=10523 | 87.4(7.6);n=19475 | 0.1 | 86.7(7.2);n=19809 | 86.6(7.2);n=10529 | 86.8(7.3);n=9280 | 0.03 |
| Potassium, mmol/L | 4.3(0.5);n=49471 | 4.3(0.4);n=15489 | 4.4(0.5);n=33982 | 0.1 | 4.3(0.4);n=29604 | 4.31(0.43);n=15483 | 4.27(0.45);n=14121 | 0.1 |
| Albumin, g/L | 41.9(3.8);n=37720 | 42.5(3.3);n=13085 | 41.5(4.0);n=24635 | 0.27* | 42.5(3.3);n=24595 | 42.5(3.3);n=13089 | 42.4(3.4);n=11506 | 0.03 |
| Sodium, mmol/L | 139.3(2.9);n=49494 | 139.1(2.7);n=15492 | 139.3(3.0);n=34002 | 0.07 | 139.3(2.7);n=29612 | 139.1(2.7);n=15486 | 139.4(2.7);n=14126 | 0.08 |
| Urea, mmol/L | 6.4(3.3);n=49483 | 5.7(2.0);n=15486 | 6.8(3.7);n=33997 | 0.34* | 5.8(2.1);n=29607 | 5.7(2.0);n=15480 | 5.8(2.2);n=14127 | 0.05 |
| Protein, g/L | 73.9(5.4);n=35490 | 74.3(4.9);n=12386 | 73.8(5.6);n=23104 | 0.1 | 74.4(5.0);n=23401 | 74.3(4.9);n=12387 | 74.5(5.0);n=11014 | 0.05 |
| Creatinine, umol/L | 92.5(71.5);n=49633 | 79.0(28.9);n=15520 | 98.6(83.2);n=34113 | 0.31* | 80.4(33.0);n=29687 | 79.0(28.9);n=15514 | 82.0(36.9);n=14173 | 0.09 |
| Alkaline phosphatase, U/L | 76.2(30.1);n=37836 | 73.5(26.1);n=13091 | 77.6(31.9);n=24745 | 0.14 | 74.8(27.2);n=24597 | 73.5(26.1);n=13095 | 76.2(28.3);n=11502 | 0.1 |
| Aspartate transaminase, U/L | 28.2(53.4);n=15006 | 28.1(26.9);n=5161 | 28.2(63.0);n=9845 | <0.01 | 28.8(30.5);n=10477 | 28.2(26.9);n=5162 | 29.4(33.6);n=5315 | 0.04 |
| Alanine transaminase, U/L | 29.4(33.6);n=32037 | 32.1(28.1);n=11145 | 28.0(36.1);n=20892 | 0.13 | 32.3(27.5);n=20438 | 32.1(28.2);n=11145 | 32.4(26.6);n=9293 | 0.01 |
| Bilirubin, umol/L | 11.3(6.7);n=37645 | 11.4(5.6);n=13060 | 11.2(7.1);n=24585 | 0.04 | 11.4(5.7);n=24545 | 11.4(5.6);n=13064 | 11.3(5.8);n=11481 | 0.01 |
| Lipid profiles |  |  |  |  |  |  |  |  |
| Triglyceride, mmol/L | 1.7(1.6);n=46943 | 1.8(1.7);n=14910 | 1.7(1.5);n=32033 | 0.06 | 1.8(1.8);n=28423 | 1.8(1.75);n=14910 | 1.82(1.84);n=13513 | 0.01 |
| Low-density lipoprotein, mmol/L | 2.4(0.8);n=46121 | 2.37(0.8);n=14658 | 2.4(0.8);n=31463 | 0.04 | 2.4(0.8);n=27937 | 2.37(0.8);n=14657 | 2.39(0.79);n=13280 | 0.03 |
| High-density lipoprotein, mmol/L | 1.2(0.3);n=46872 | 1.16(0.31);n=14885 | 1.21(0.33);n=31987 | 0.15 | 1.2(0.3);n=28370 | 1.16(0.31);n=14885 | 1.18(0.32);n=13485 | 0.06 |
| Total cholesterol, mmol/L | 4.3(1.0);n=46986 | 4.3(1.0);n=14929 | 4.4(1.0);n=32057 | 0.04 | 4.3(1.0);n=28445 | 4.3(1.0);n=14929 | 4.4(1.0);n=13516 | 0.05 |
| Hemoglobin A1C, % | 8.1(1.5);n=48920 | 8.3(1.6);n=15353 | 8.0(1.5);n=33567 | 0.21* | 8.3(1.6);n=29345 | 8.3(1.57);n=15352 | 8.26(1.61);n=13993 | 0.03 |
| Fasting glucose, mmol/L | 9.9(3.3);n=34323 | 10.1(3.2);n=11828 | 9.8(3.4);n=22495 | 0.08 | 10.1(3.3);n=22141 | 10.1(3.2);n=11841 | 10.0(3.5);n=10300 | 0.02 |

The cumulative incidence of primary and secondary outcomes after propensity score matching is shown in **Table 2A**. The cumulative incidences of these outcomes stratified by initial drug exposure age, drug use, the combination of gender and drug exposure and combination of age and drug exposure effects are summarised by cumulative incidence curves **Figures 2A** and **2B**. Gender-based and age-based trends in the incidence of the different outcomes are shown in **Supplementary Figures 3A** and **3B**. Furthermore, summary figures of comparing annual incidence ratios with 95% CIs of different adverse events stratified by drug use are presented in **Figure 3A**.

**Table 2A.** Annualized incidence rate (IR) per 1000 person-year of primary and secondary cancer outcomes, all-cause mortality and cancer related mortality in the cohort before and after 1:1 propensity score matching.

| Before matching |  |  |  | After 1:1 propensity score matching |  |  |  |
| --- | --- | --- | --- | --- | --- | --- | --- |
| All-cause mortality |  |  |  |  |  |  |  |
| Overall | Person-year | Events | IR [95% CI] | Overall | Person-year | Events | IR [95% CI] |
|  | 3.29 x10^5 | 3033 | 9.2 [8.6-9.6] |  | 2.01 x10^5 | 970 | 4.8 [4.5-5.1] |
| Cancer-related mortality |  |  |  |  |  |  |  |
| Overall | Person-year | Events | IR [95% CI] | Overall | Person-year | Events | IR [95% CI] |
|  | 3.29 x10^5 | 506 | 1.5 [1.4-1.7] |  | 2.01 x10^5 | 211 | 1.1 [0.9-1.2] |
| New-onset all-cause cancer |  |  |  |  |  |  |  |
| Overall | Person-year | Events | IR [95% CI] | Overall | Person-year | Events | IR [95% CI] |
|  | 3.26 x10^5 | 1533 | 4.7 [4.5-4.9] |  | 2.00 x10^5 | 674 | 3.4 [3.1-3.6] |
| New-onset lung cancer |  |  |  |  |  |  |  |
| Overall | Person-year | Events | IR [95% CI] | Overall | Person-year | Events | IR [95% CI] |
|  | 3.28 x10^5 | 249 | 0.8 [0.7-0.9] |  | 2.01 x10^5 | 124 | 0.6 [0.5-0.7] |
| New-onset gastrointestinal cancer |  |  |  |  |  |  |  |
| Overall | Person-year | Events | IR [95% CI] | Overall | Person-year | Events | IR [95% CI] |
|  | 3.27 x10^5 | 817 | 2.5 [2.3-2.7] |  | 2.00 x10^5 | 325 | 1.6 [1.5-1.8] |
| New-onset bladder cancer |  |  |  |  |  |  |  |
| Overall | Person-year | Events | IR [95% CI] | Overall | Person-year | Events | IR [95% CI] |
|  | 3.29 x10^5 | 97 | 0.3 [0.2-0.4] |  | 2.01 x10^5 | 50 | 0.2 [0.2-0.3] |
| New-onset genitourinary cancer |  |  |  |  |  |  |  |
| Overall | Person-year | Events | IR [95% CI] | Overall | Person-year | Events | IR [95% CI] |
|  | 3.28 x10^5 | 261 | 0.8 [0.7-0.9] |  | 2.01 x10^5 | 121 | 0.6 [0.5-0.7] |

**Figure 2A.**
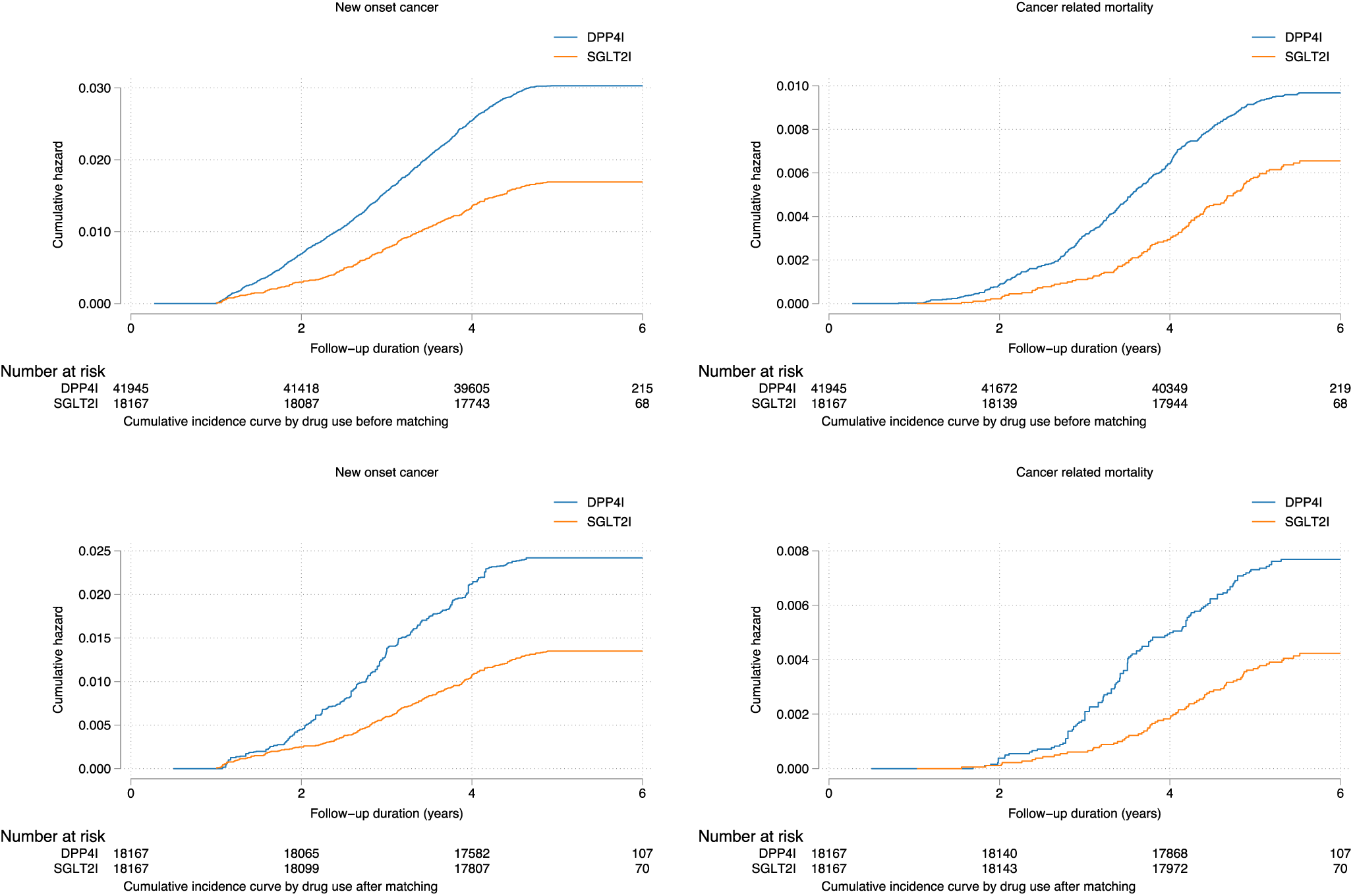
Cumulative incidence curves for new onset cancer and cancer related mortality stratified by drug exposure effects of SGLT2I and DPP4I before and after propensity score matching (1:1).

**Figure 2B.**
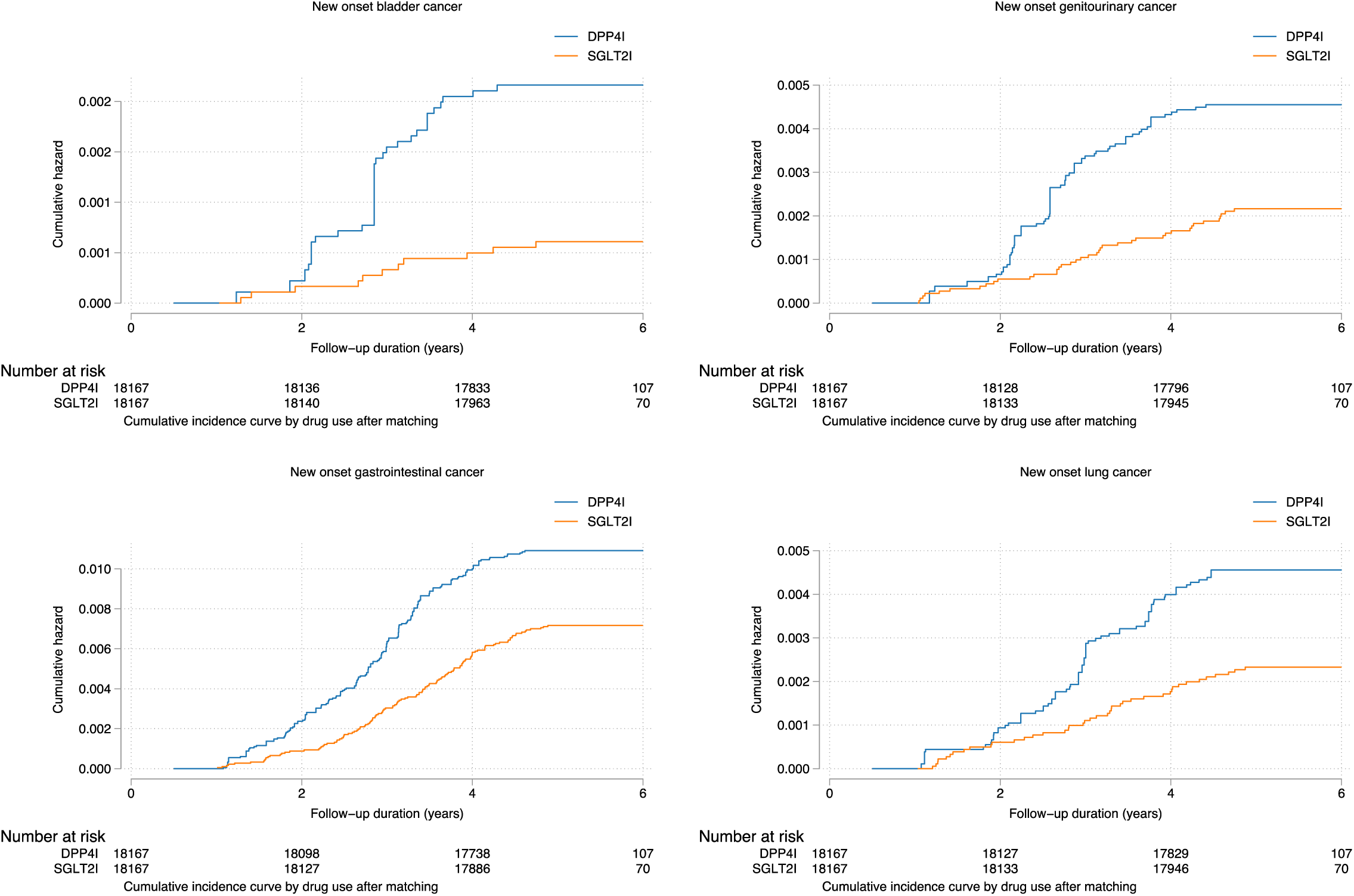
Cumulative incidence curves for different new onset cancer outcomes stratified by drug exposure effects of SGLT2I and DPP4I in the matched cohort.

**Figure 3A.**
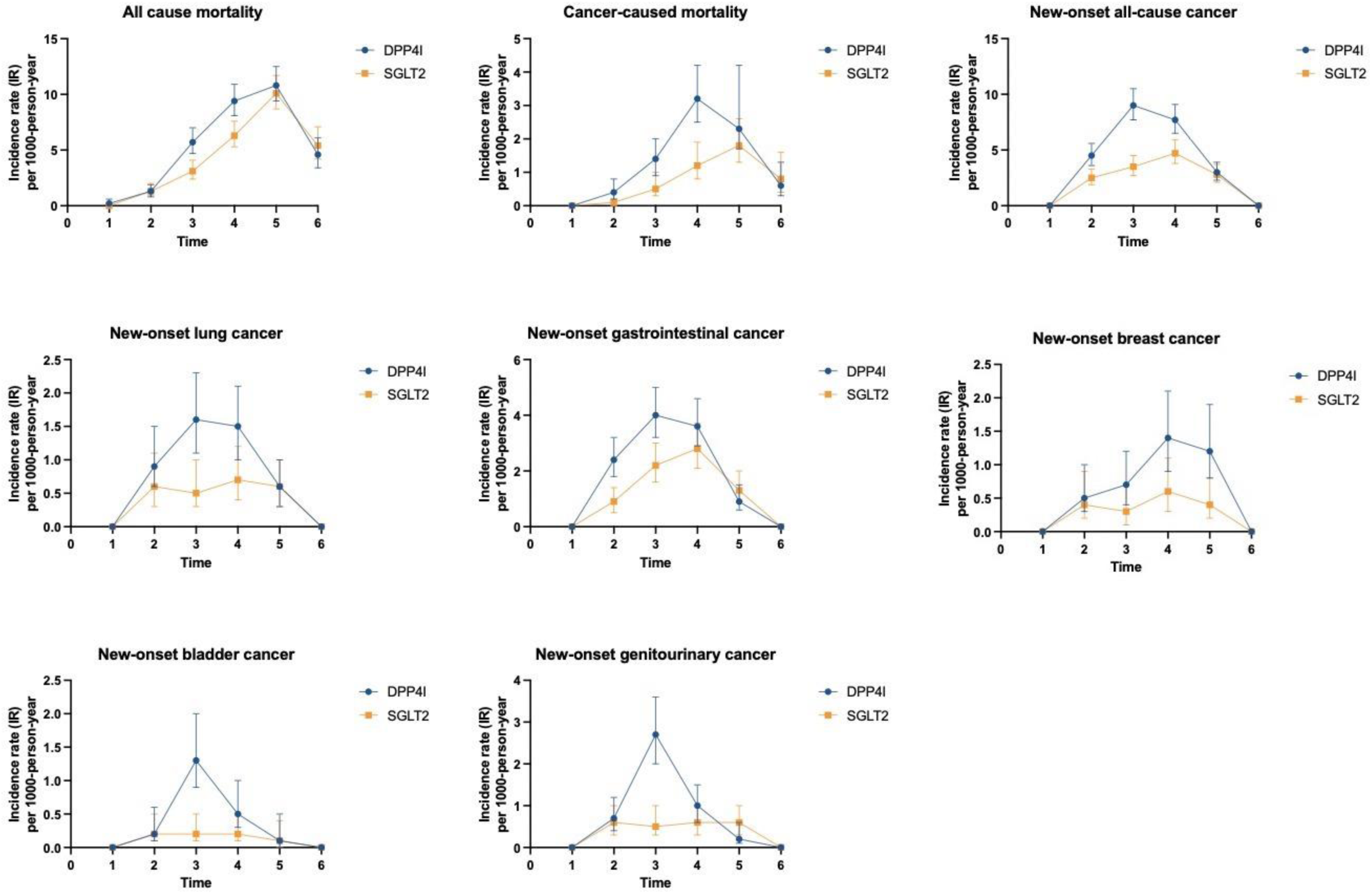
Summary figures of comparing annual incidence ratios with 95% CIs of different adverse events stratified by drug use.

### Cox regression

The results of univariate Cox regression analysis for predicting general and pre-specified cancer risk are displayed in **Supplementary Table 4A** and **Supplementary Table 4B**. Significant variables in univariate regression were subsequently incorporated into multivariate models to evaluate the relationship between SGLT2I and DPP4I with malignancy. As shown in **Table 2B**, after adjustment for significant demographics, past comorbidities, non-SGLT2I/DPP4I medications, abbreviated MDRD, fasting glucose, HbA1c, and duration from earliest diabetes mellitus date to initial drug exposure date, SGLT2I were associated with a comparatively decreased risk of all-cause mortality (HR: 0.92; 95% CI: 0.84-0.99; P = 0.04), cancer-related mortality (HR: 0.58; 95% CI: 0.42-0.80; P = < 0.001), as well as a 30% reduction in the risk of new-onset overall cancer (HR: 0.70; 95% CI: 0.59-0.84; P = < 0.001). When stratified by cancer subtype, SGLT2I were related to a lower risk of new-onset breast cancer (HR: 0.51; 95% CI: 0.32-0.80; P = < 0.001), but not with other malignancies. With subgroup analysis comparing DPP4I to different subtypes of SGLT2I, dapagliflozin (HR: 0.78; 95% CI: 0.64-0.95; P = 0.01) and ertugliflozin (HR: 0.65; 95% CI: 0.43-0.98; P = 0.04) both demonstrated superiority in relation to new-onset overall cancer development, with the former also presenting with a relatively lower risk of breast cancer (HR: 0.48; 95% CI: 0.27-0.83; P = 0.001). There were no observable differences when comparing the use of either Canagliflozin or Empagliflozin with DPP4I in terms of overall or specific cancer risk.

**Table 2B.**
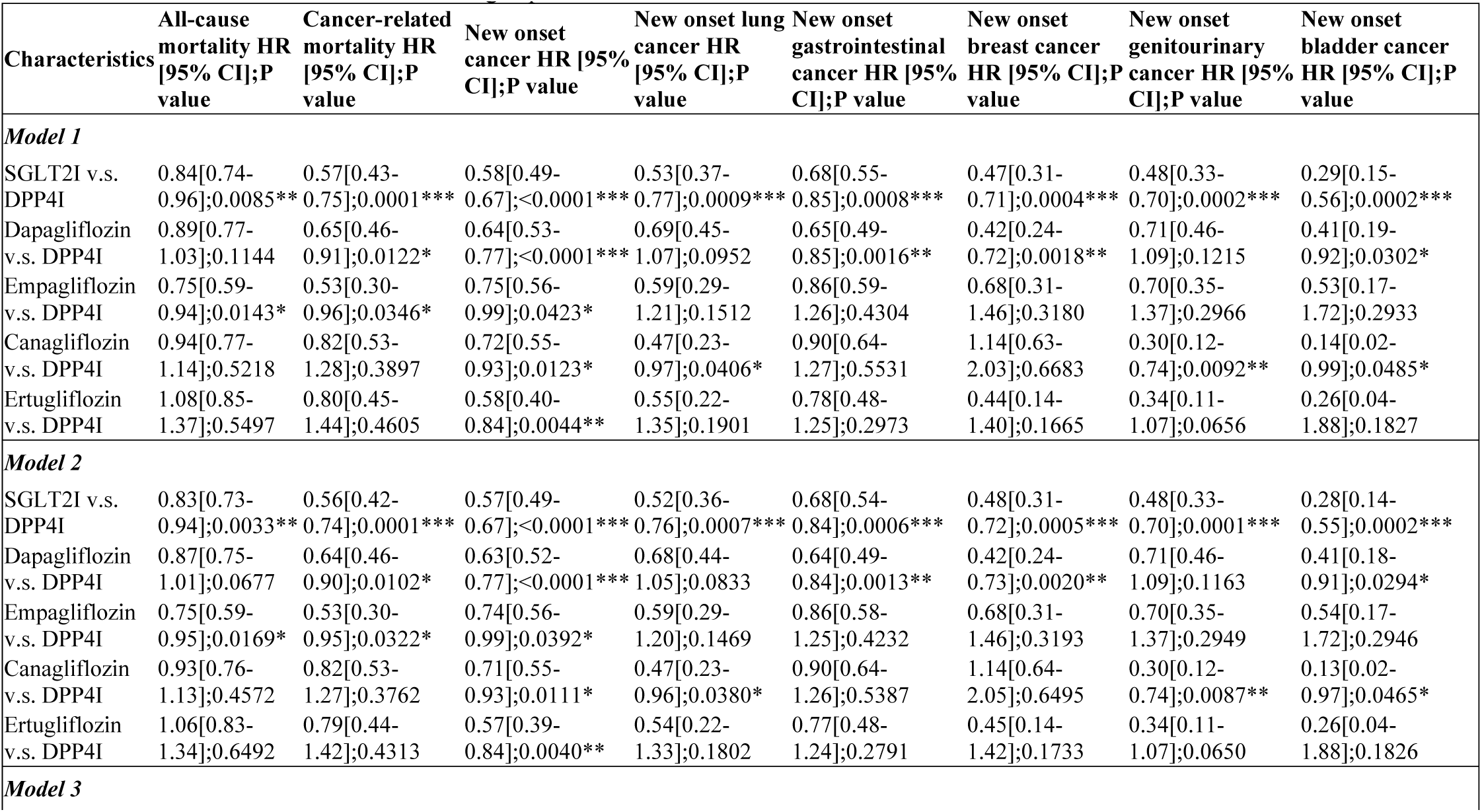
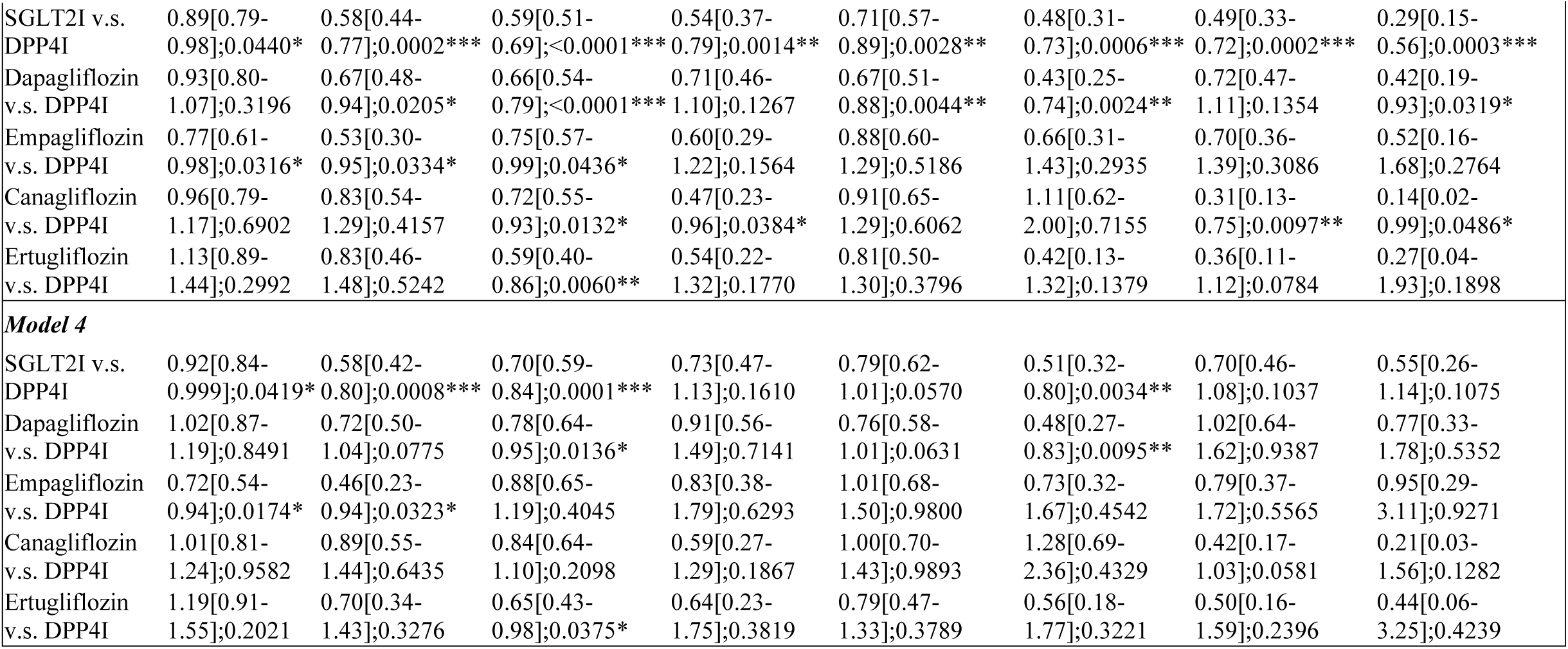
Multivariate Cox regression models with adjustments to predict new all-cause cancers in the matched cohort. * for p≤ 0.05, ** for p ≤ 0.01, *** for p ≤ 0.001; HR: hazard ratio; CI: confidence interval; SGLT2I: sodium glucose cotransporter-2 inhibitor; DPP4I: dipeptidyl peptidase-4 inhibitor. Model 1 adjusted for significant demographics. Model 2 adjusted for significant demographics, and past comorbidities. Model 3 adjusted for significant demographics, past comorbidities, and non-SGLT2I/DPP4I medications. Model 4 adjusted for significant demographics, past comorbidities, non-SGLT2I/DPP4I medications, abbreviated MDRD, fasting glucose, HbA1c, and duration from earliest diabetes mellitus date to initial drug exposure date.

### Sensitivity analysis

To assess the predictivity of the models, sensitivity analysis was conducted to evaluate the effect of matching on the results, namely with inverse probability of treatment weighting (**Supplementary Table 5)**. The findings confirmed those of univariate cox regression, that SGLT2I administration was still associated with a lower risk of all-cause mortality, cancer-related mortality, new-onset overall cancer as well as all pre-specified cancers (lung, breast, gastrointestinal, genitourinary and bladder) when compared to DPP4I usage.

## Discussion

To the best of our knowledge, this is the first territory-wide study that does a direct comparison of the effect of SGLT2I and DPP4I on overall and pre-specified cancer risk in a cohort of Asian patients. The main findings of this study are as follows: in comparison to DPP4I, i) SGLT2I were associated with a lower risk of all-cause mortality, cancer-related mortality and new-onset overall cancer; ii) SGLT2I were related to a lower risk of new-onset breast cancer; iii) when stratified according to the medication subtype, dapagliflozin and ertugliflozin both demonstrated a reduced risk of new-onset malignancy, with the former also presenting with a lower risk of breast cancer.

Antidiabetic medications are among the most commonly prescribed drugs in the world, with the indications of some expanding beyond T2DM to other non-diabetic cardiovascular and chronic kidney conditions (18, 19). The clinical practicality of these medications, coupled with their multifaceted systemic effects, warrants a thorough assessment of the safety of their long-term usage, which has raised some important concerns in recent years. This is of specific importance concerning the comparatively newer classes of oral hypoglycemic drugs, namely DPP4 inhibitors (DPP4I) and SGLT2 inhibitors (SGLT2I), the first of which were marketed in 2006 (Sitagliptin) and 2013 (Canagliflozin), respectively (20). Given the chronicity with which these medications are taken, a particularly significant outcome that is evaluated, unsurprisingly, is a cancer risk.

The majority of the comparative studies available in existing literature have evaluated cancer risk across older diabetic drugs. Liu *et al*. performed a retrospective case-controlled prognostic assessment for different anti-diabetic medications, including metformin, thiazolidinediones, sulfonylureas, meglitinides, acarbose as well as insulin and its analogues, in turn revealing that aside from pioglitazone and insulin, the other therapies failed to show an association with cancer incidence. This relationship was maintained when stratifying outcomes by cancer type namely for pancreatic, liver and lung cancer (21). In addition to this, certain investigations have demonstrated the protective effect of some of the older classes against cancer, most notably with metformin, which has demonstrated either a reduced association with cancer (22, 23), or a lower incidence of cancer on follow-up relative to other antidiabetic medications (24).

Despite this, it should be noted that there is much more uncertainty about the malignancy risk of the somewhat newer anti-diabetic medications. Regarding DPP4I, a meta-analysis compiled by Zhao *et al*. did not report any association between these medications and malignancy, even when stratified by different subtypes of DPP4I (12). Similarly, the findings of another meta-analysis lend further credence to this notion by not only failing to show a relationship with malignancy development but also purporting a potential protective effect of DPP4I against colorectal cancer (25). However, although these results may reflect much of the current school of thought concerning DPP4I, there have been some recent investigations that have suggested the possible existence of either a dose-dependent or cancer-type-dependent correlation. As to the former, Chou *et al*. presented a higher incidence of colorectal cancer in patients on DPP4I who were receiving a high cumulative daily dose, but a corresponding lower risk of colorectal cancer amongst low cumulative dose users (26). As it pertains to the latter, there is evidence to suggest that whilst a relationship between DPP4I and overall cancer risk may not exist, these drugs are associated with specific cancer types when categorized, namely bladder, kidney and liver cancer as well as melanoma (11).

Likewise, very much akin to that of DPP4I, the data centred around SGLT2I is also controversial. Most recently, a meta-analysis performed by Benedetti *et al*. proposed a reduced cancer risk of SGLT2I when compared to placebo, with particular efficacy for dapagliflozin and ertugliflozin (27). These results are in line with that of the present study, which also demonstrated the superiority of dapagliflozin and ertugliflozin in relation to cancer risk. Such findings are further emphasized by that of Pelletier et al, which also failed to display an increased cancer risk with SGLT2I users, regardless of cancer type (28). However, the obscurity in the findings concerning SGLT2I primarily resides in the fact that the malignancy risk varies depending on the SGLT2I and cancer subtypes. Tang *et al*. showed that although the overall cancer incidence is lower with SGLT2I relative to other comparator drugs when analysing pre-specified cancers, empagliflozin demonstrated a higher risk of bladder cancer whilst canagliflozin exhibited protective effects against gastrointestinal cancers (10). The ambiguity regarding SGLT2I is further compounded with other contrarian evidence suggesting a reduced risk of malignancy with empagliflozin relative to other oral hypoglycaemic agents, but instead, an increased risk when compared to placebo (29).

Given the relatively newer status of DPP4I and SGLT2I, there is a paucity of literature comparing the non-diabetic outcomes associated with these medications. Au *et al*. showcased a reduced incidence of pneumonia and pneumonia-related mortality with SGLT2I relative to DPP4I in patients from Hong Kong (30). In a Taiwanese cohort, SGLT2I similarly exhibited superiority in the risk of gout development when compared to DPP4I (13). Moreover, one retrospective study in a Taiwanese cohort demonstrated that SGLT2I usage was associated with a lower risk of cancer-related mortality compared to DPP4I, akin to the results to our investigations (12). To date, in addition to our study, there is only one other that has directly assessed these two classes of medication and their respective cancer risks. The findings from this study indicated that the risk of a urinary tract and haematological malignancy with SGLT2I was half that of with DPP4I, albeit there were no other differences amongst other cancer subtypes (14). These findings are supported by that of this study, which has likewise showcased that the use of SGLT2I is associated with a 30% reduction in new-onset overall cancer risk in comparison to DPP4I, though there were no observable differences in genitourinary or bladder malignancy development between the two drug classes.

## Limitations

There are certain limitations present in this population-based study. Firstly, due to the observational nature of this study, acquired results may be susceptible to information bias due to missing data, coding errors or under coding. Secondly, the retrospective nature of the study suggests that all derived findings regarding the relationship between SGLT2I, DPP4I and new-onset overall cancer were correlational in nature. Thirdly, information on drug exposure could not be directly obtained, and was instead determined indirectly through prescription refills, which may pose a liability concern. Fourthly, as the drug exposure duration could not be standardised, this may have influenced the primary and secondary outcomes of the study. Finally, due to the lack of codes in CDARS, information regarding medical history, such as smoking status, were unattainable and could have been a confounding variable to cancer risk.

## Conclusions

In conclusion, SGLT2I was associated with lower risk of all-cause mortality and cancer-related mortality, as well as a 30% reduction in the risk of new-onset overall cancer when compared to DPP4I. When stratified by cancer subtype, SGLT2I demonstrated superiority in relation to new-onset breast cancer risk, but not other types of malignancies. These findings, coupled with the aforementioned drug subtype specific associations demonstrated, suggests the potential importance of using SGLT2I over DPP4I for patients with high risk factors for cancer development. However, further cohort studies of a prospective nature are still needed to not only validate these results, but also guide risk stratification and management in the clinical setting.

## Data Availability

All data produced in the present study are available upon reasonable request to the authors

## Availability of data and materials

An anonymised version of the dataset without identifiable or personal information is available from the corresponding authors upon reasonable request for research purposes.

## Acknowledgements

None.

## Author’s Contribution

CTC, IL, OHIC: study conception, data analysis, data extraction, data interpretation, statistical analysis, manuscript drafting, critical revision of manuscript

TL, CD, KN, WTW, TL, SL, QZ, BC: data acquisition, data interpretation, critical revision of manuscript

JD, GT: study conception, study supervision, project planning, data interpretation, statistical analysis, manuscript drafting, critical revision of manuscript

