## Supplementary Table and Figure for "Sodium-glucose cotransporter 2 inhibitors versus dipeptidyl peptidase 4 inhibitors on new-onset overall cancer in type 2 diabetes mellitus: a population-based study"

**Supplementary Appendix**

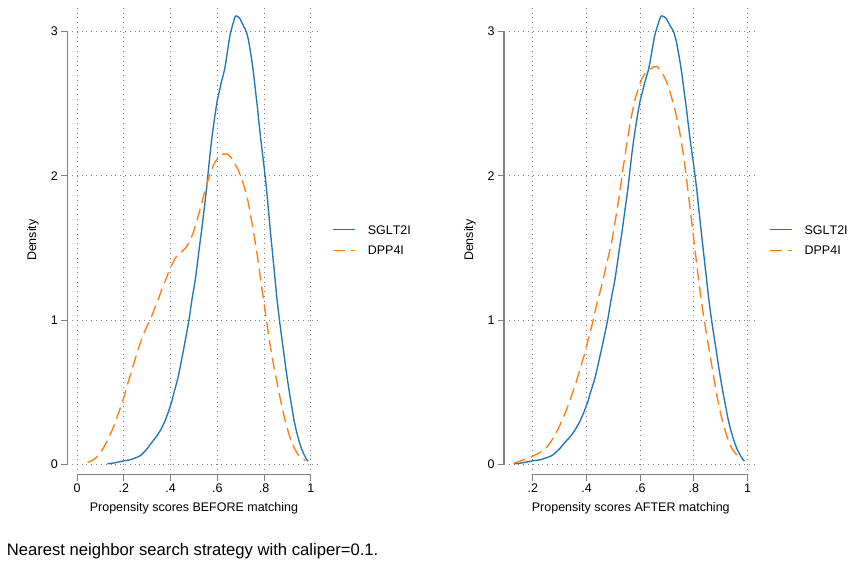

**Supplementary Figure 1. Propensity score matching comparisons and proportional hazard assumption checking with parallel lines for SGLT2I v.s. DPP4I before and after 1:1 matching with nearest neighbor search strategy with caliper of 0.1**

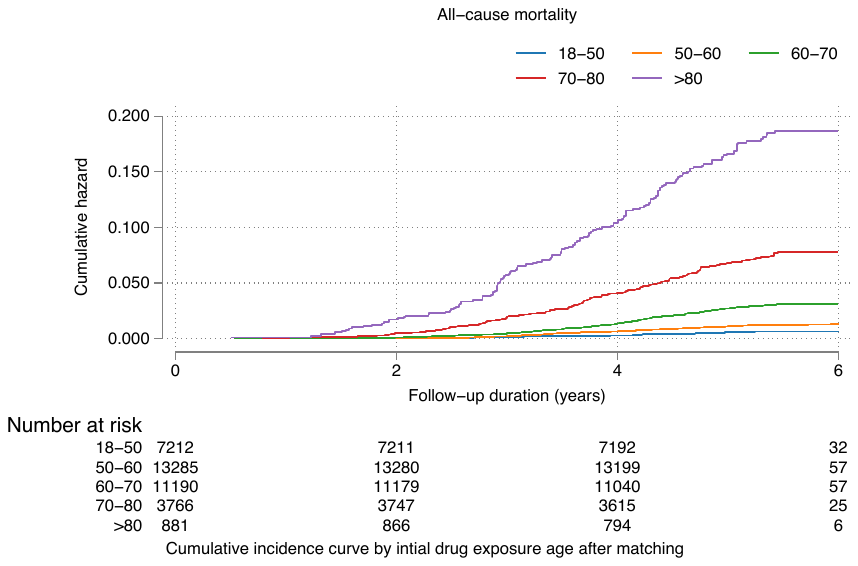

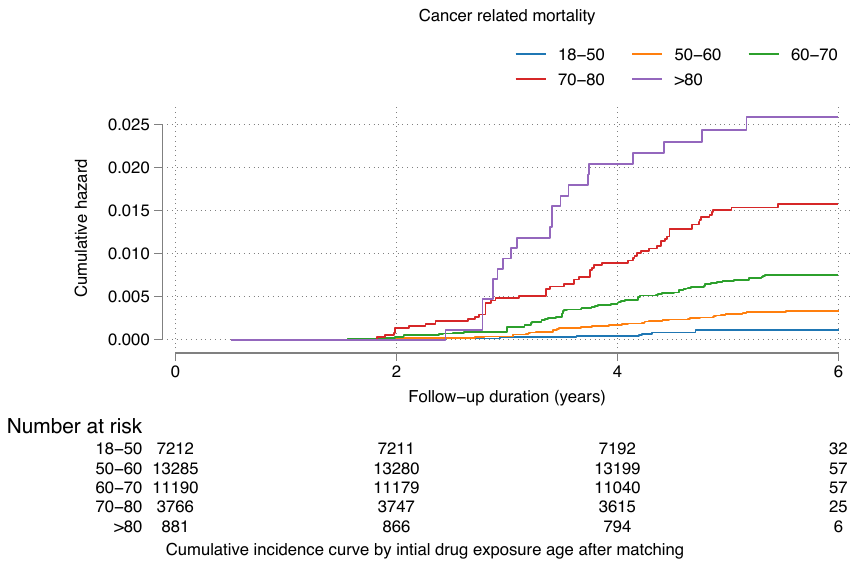

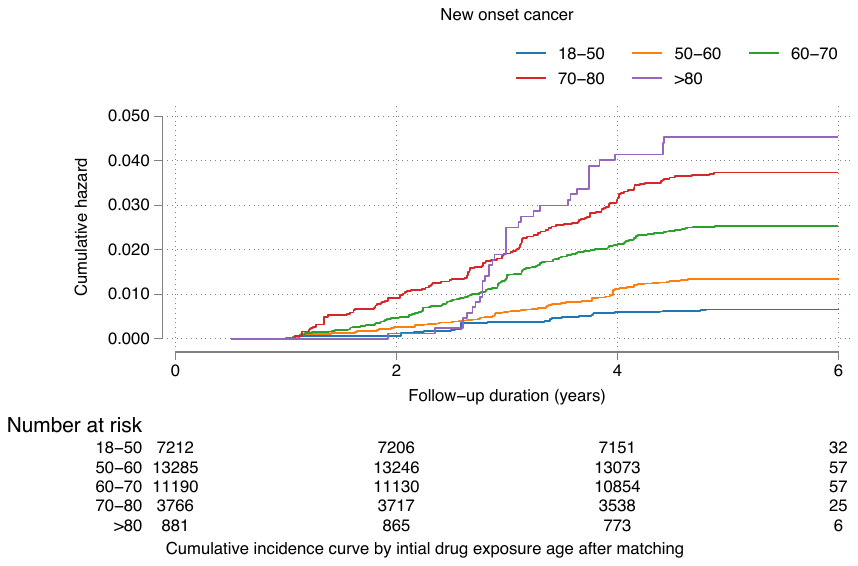

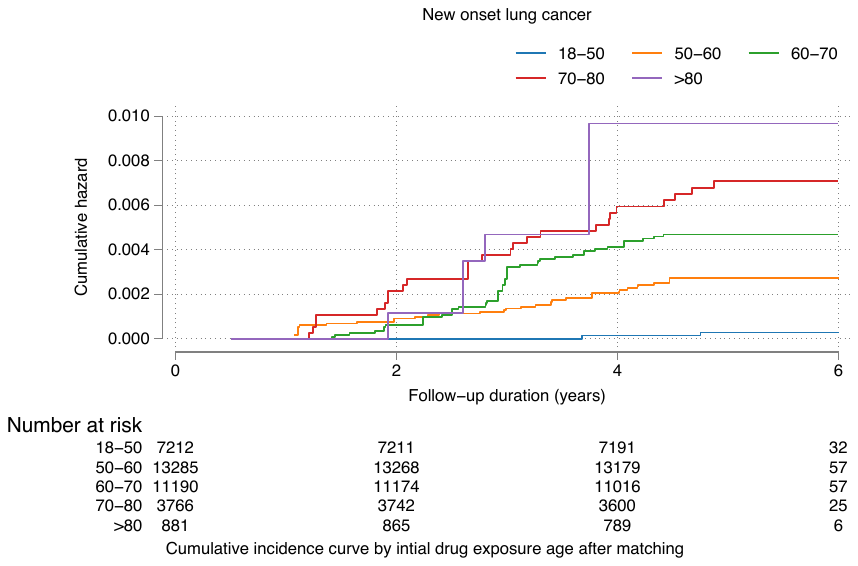

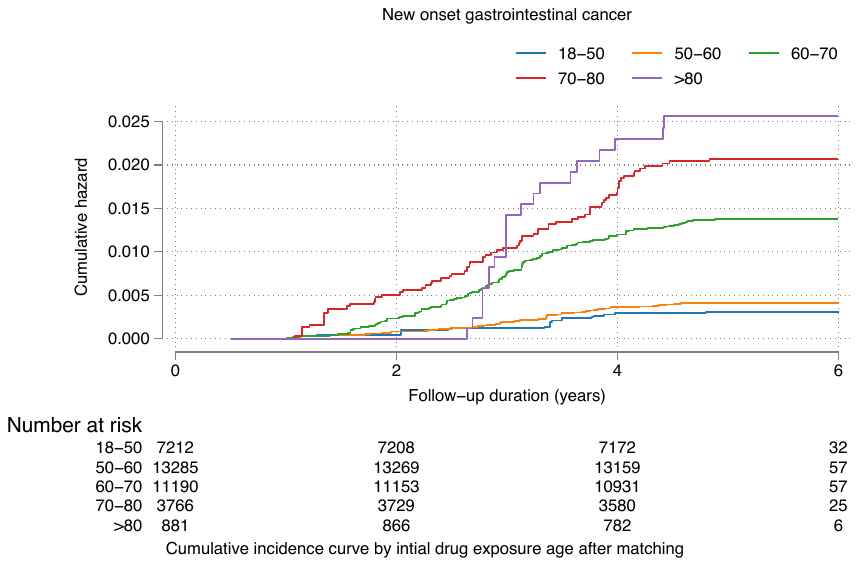

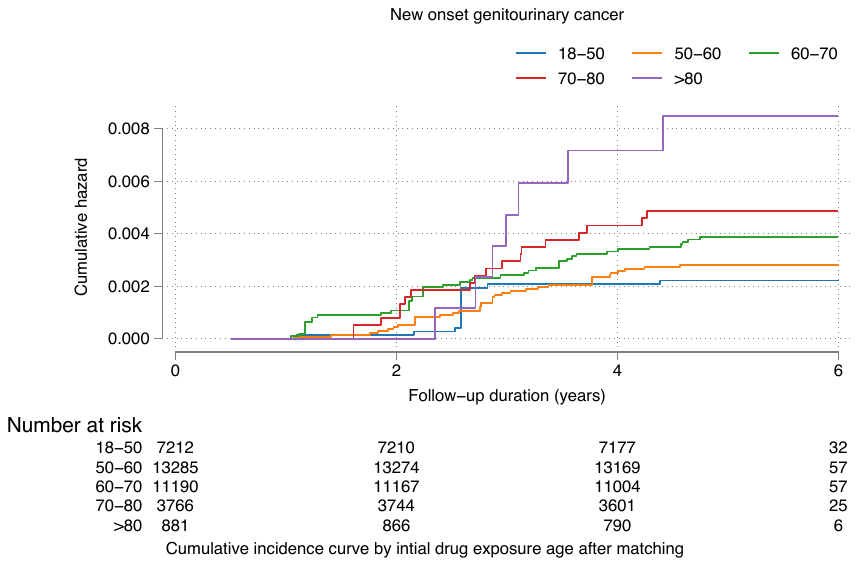

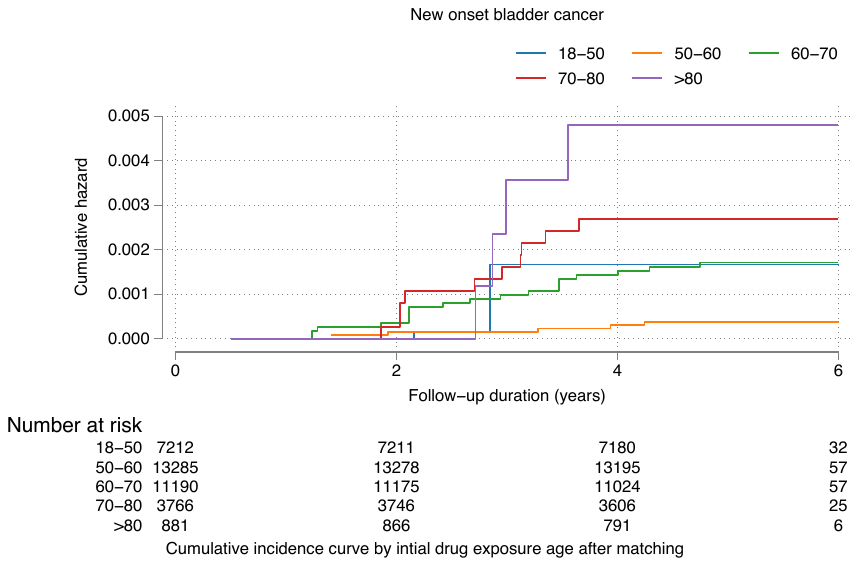

**Supplementary Figure 2. Cumulative incidence curves for primary and secondary outcomes stratified by initial drug exposure age in the matched cohort.**

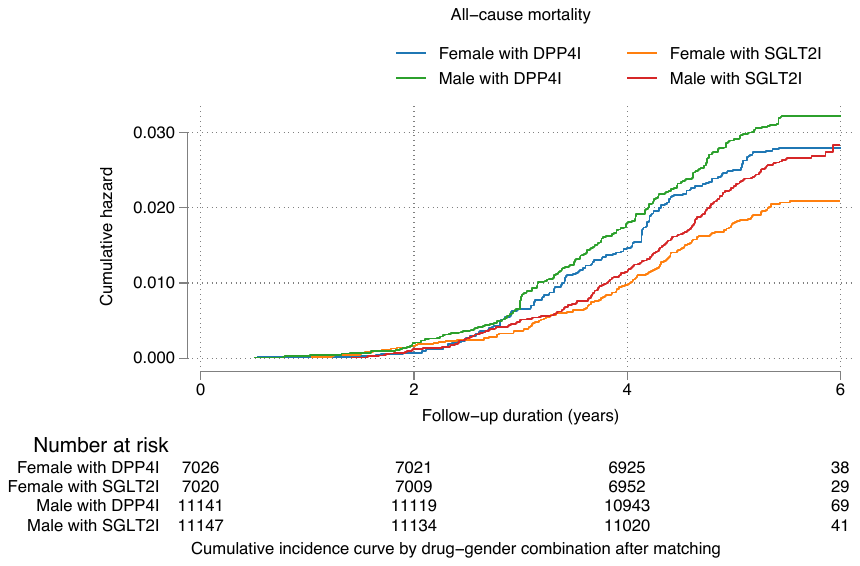

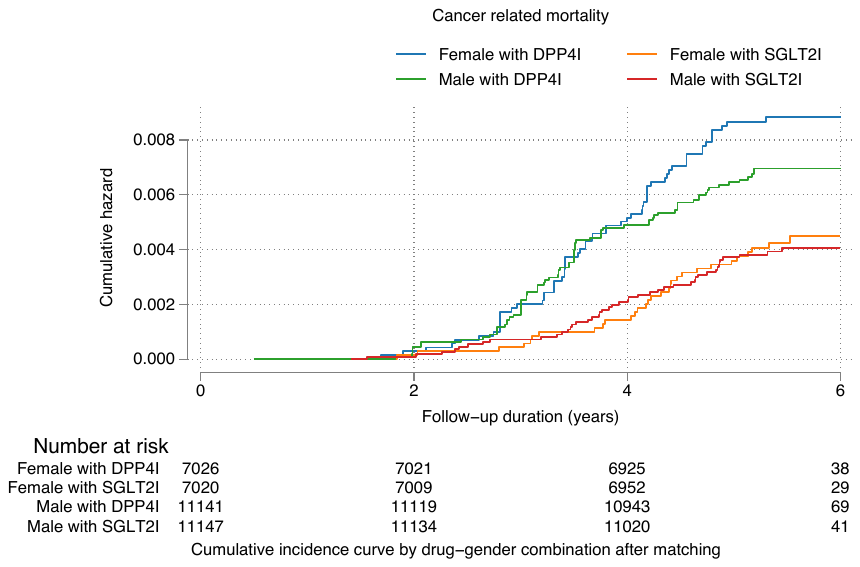

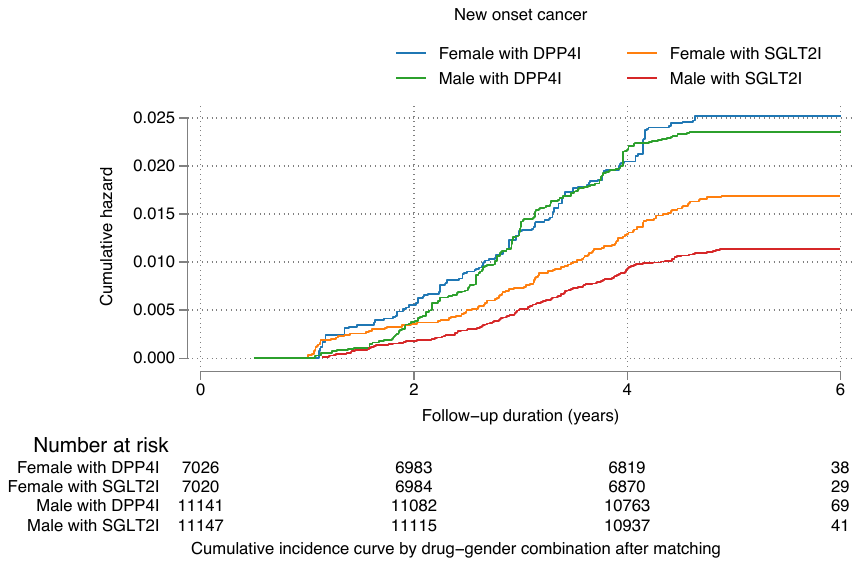

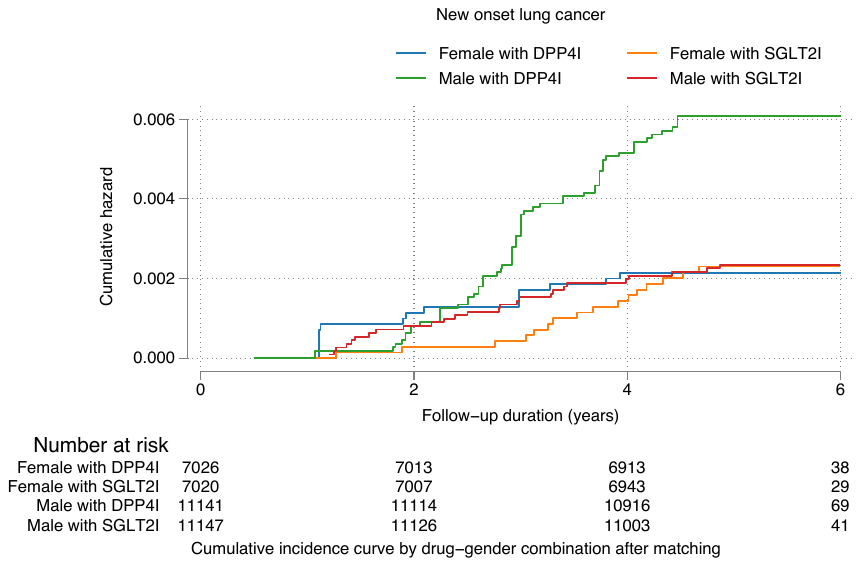

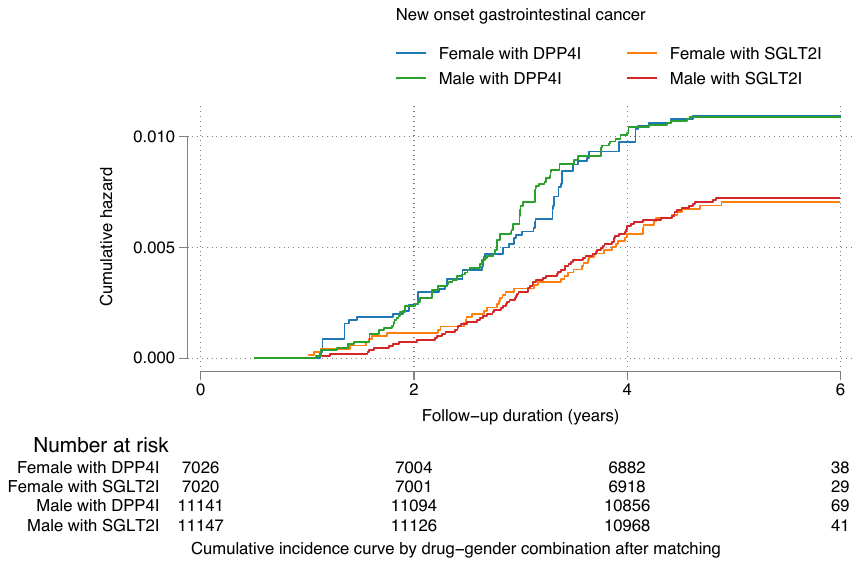

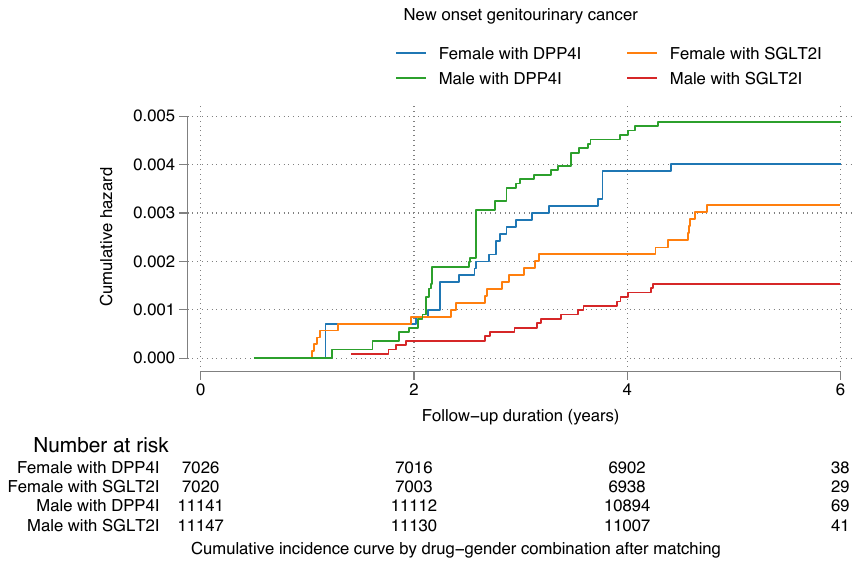

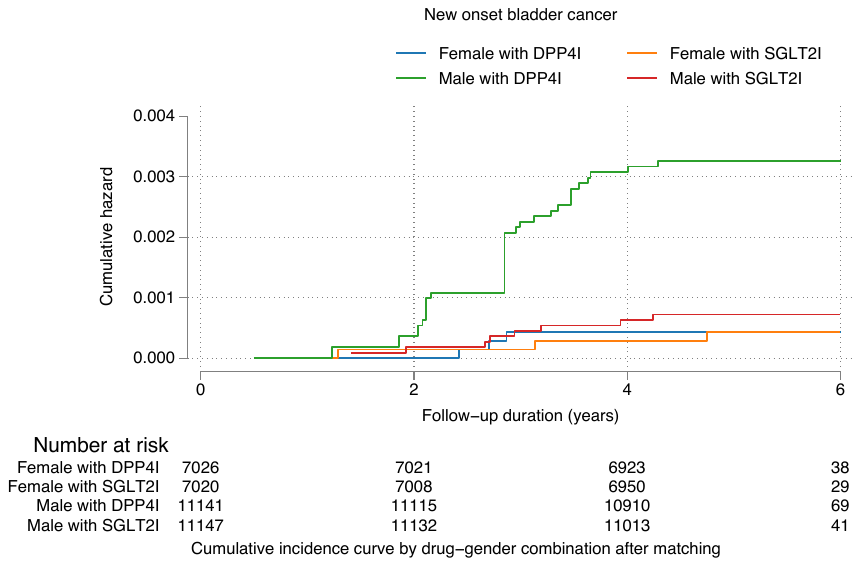

**Supplementary Figure 3A. Cumulative incidence curves for new onset cancer stratified by combinations of gender and drug exposure effects of SGLT2I and DPP4I in the matched cohort**

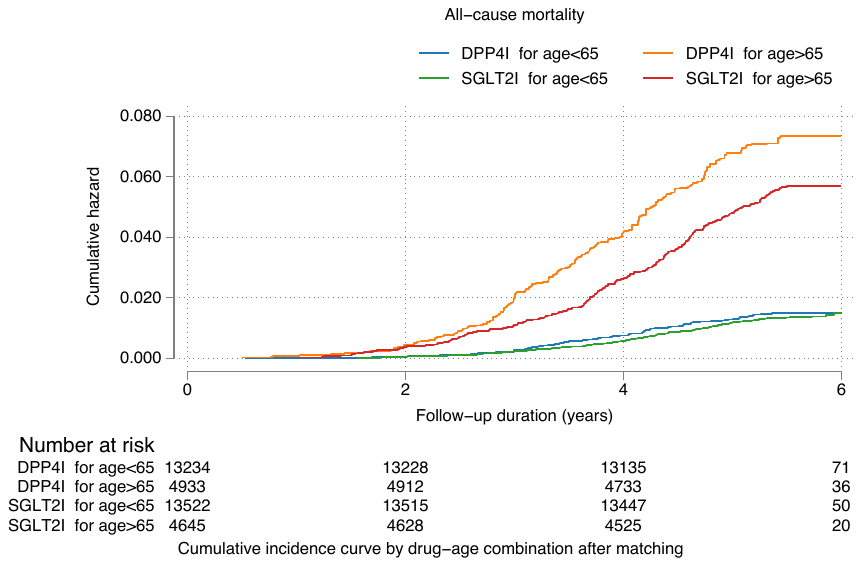

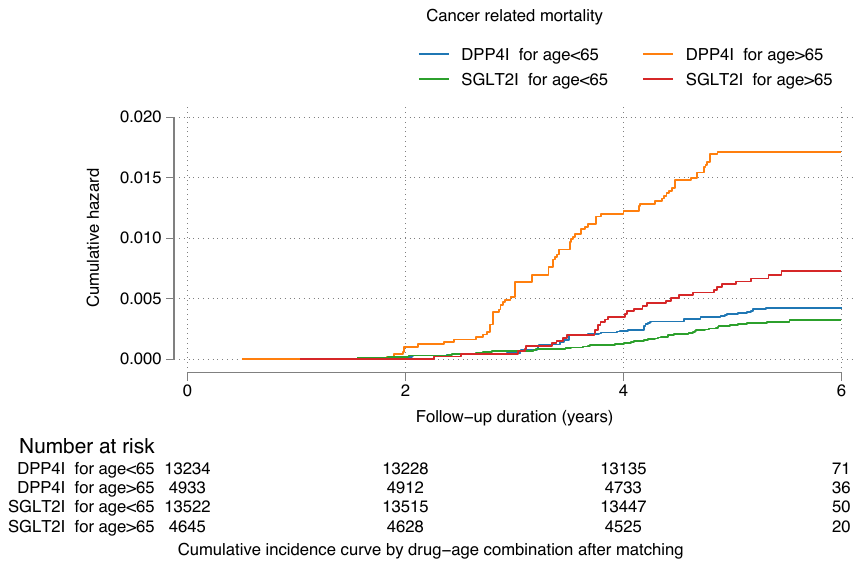

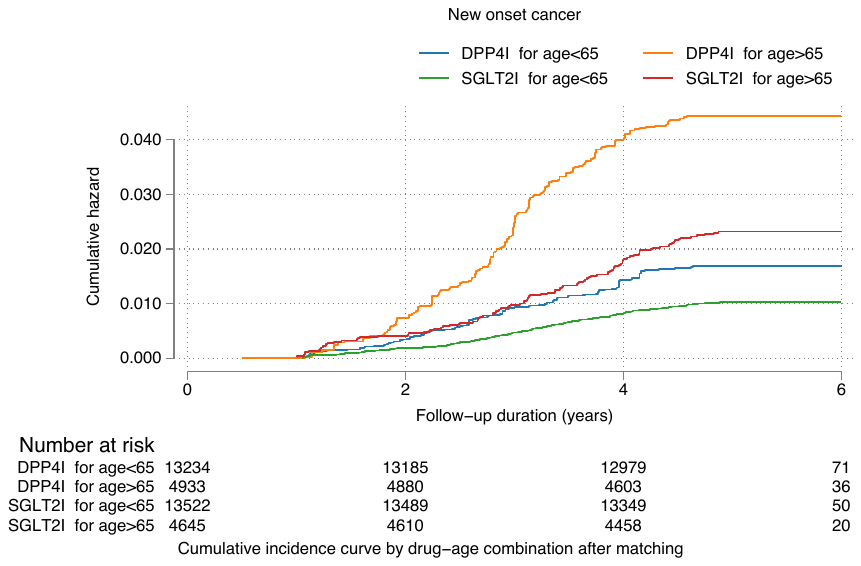

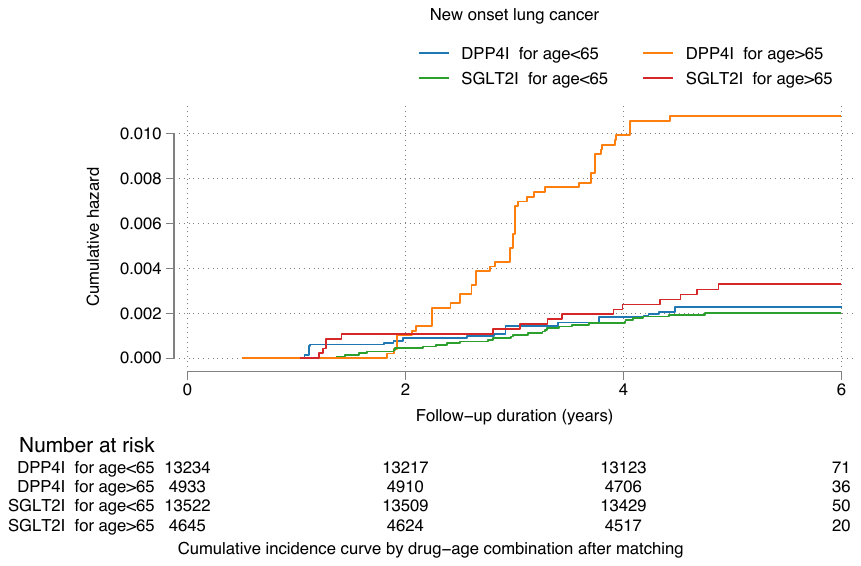

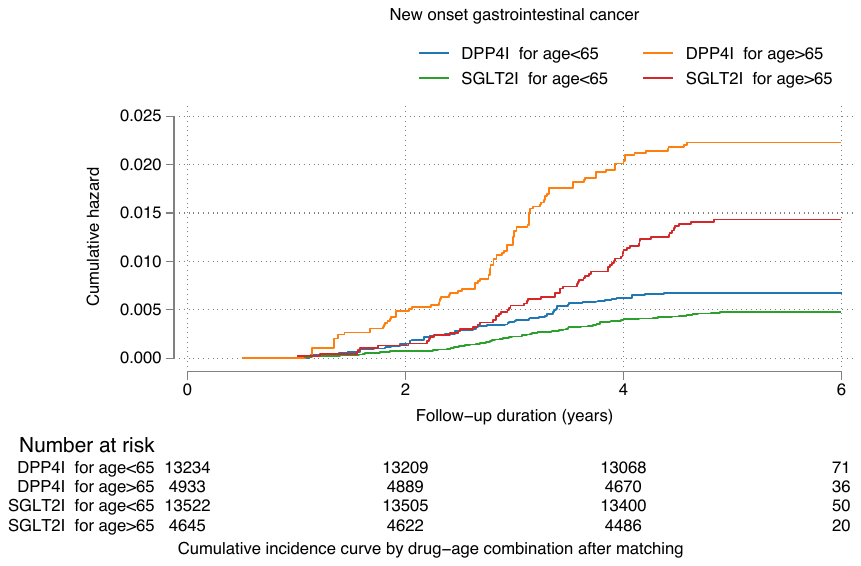

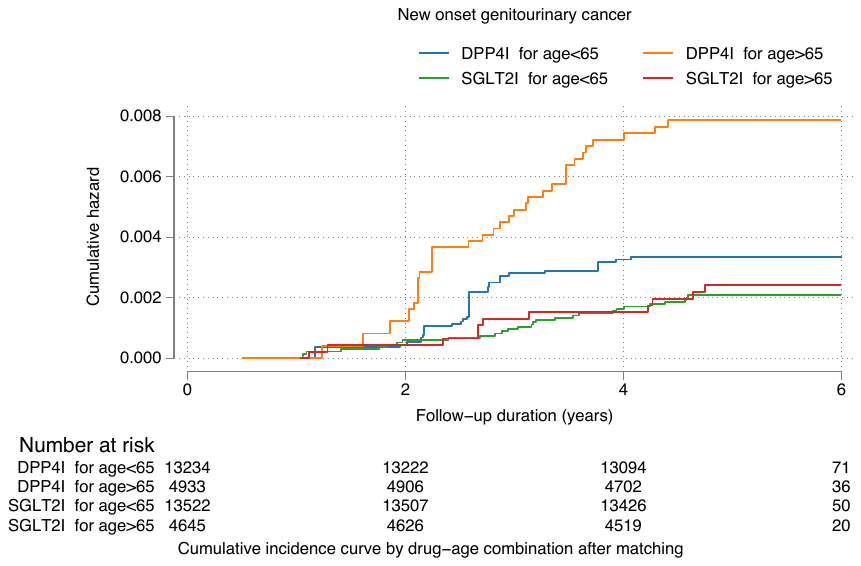

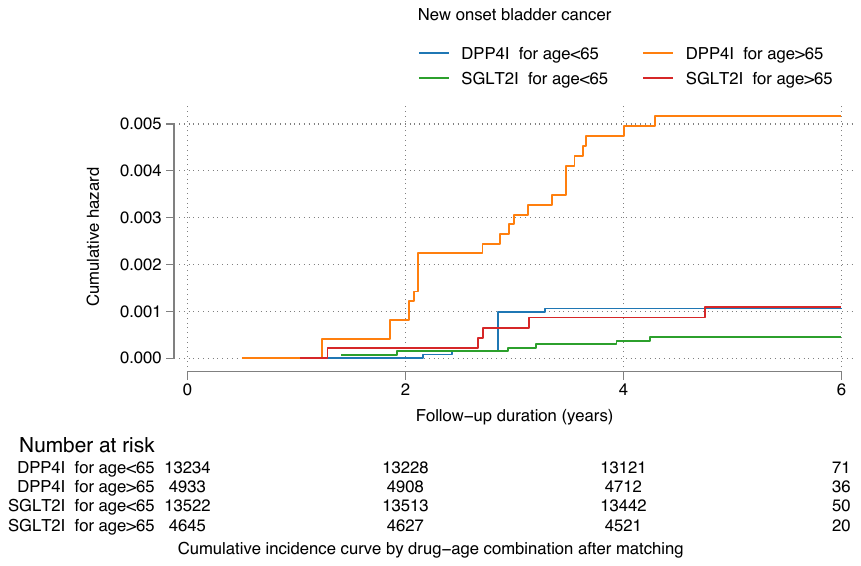

**Supplementary Figure 3B. Cumulative incidence curves for primary and secondary outcomes stratified by combinations of age and drug exposure effects of SGLT2I and DPP4I in the matched cohort**

**Supplementary Table 1. ICD9 codes for comorbidities and ICD10 codes for cancer-related mortality**

**ICD9 codes for comorbidities**

**Cancer**: 140 140.1 140.3 140.4 140.5 140.6 140.8 140.9 141 141.1 141.2 141.3 141.4 141.5 141.6 141.8 141.9 142 142.1 142.2 142.8 142.9 143 143.1 143.8 143.9 144 144.1 144.8 144.9 145 145.1 145.2 145.3 145.4 145.5 145.6 145.8 145.9 146 146.1 146.2 146.3 146.4 146.5 146.6 146.7 146.8 146.9 147 147.1 147.2 147.3 147.8 147.9 148 148.1 148.2 148.3 148.8 148.9 149 149.1 149.8 149.9 150 150.1 150.2 150.3 150.4 150.5 150.8 150.9 151 151.1 151.2 151.3 151.4 151.5 151.6 151.8 151.9 152 152.1 152.2 152.3 152.8 152.9 153 153.1 153.2 153.3 153.4 153.5 153.6 153.7 153.8 154 154.1 154.2 154.3 154.8 155 155.1 155.2 156 156.1 156.2 156.8 156.9 157 157.1 157.2 157.3 157.4 157.8 157.9 158 158.8 158.9 159 159.1 159.8 159.9 160 160.1 160.2 160.3 160.4 160.5 160.8 160.9 161 161.1 161.2 161.3 161.8 161.9 162 162.2 162.3 162.4 162.5 162.8 162.9 163 163.1 163.8 163.9 164 164.1 164.2 164.3 164.8 164.9 165 165.8 165.9 170 170.1 170.2 170.3 170.4 170.5 170.6 170.7 170.8 170.9 171 171.2 171.3 171.4 171.5 171.6 171.7 171.8 171.9 172 172.1 172.2 172.3 172.4 172.5 172.6 172.7 172.8 172.9 173 173.01 173.02 173.09 173.1 173.11 173.12 173.19 173.2 173.21 173.22 173.29 173.3 173.31 173.32 173.39 173.4 173.41 173.42 173.49 173.5 173.51 173.52 173.59 173.6 173.61 173.62 173.69 173.7 173.71 173.72 173.79 173.8 173.81 173.82 173.89 173.9 173.91 173.92 173.99 174 174.1 174.2 174.3 174.4 174.5 174.6 174.8 174.9 175 175.9 176 176.1 176.2 176.3 176.4 176.5 176.8 176.9 179 180 180.1 180.8 180.9 181 182 182.1 182.8 183 183.2 183.3 183.4 183.5 183.8 183.9 184 184.1 184.2 184.3 184.4 184.8 184.9 185 186 186.9 187 187.1 187.2 187.3 187.4 187.5 187.6 187.7 187.8 187.9 188 188.1 188.2 188.3 188.4 188.5 188.6 188.7 188.8 188.9 189 189.1 189.2 189.3 189.4 189.8 189.9 190 190.1 190.2 190.3 190.4 190.5 190.6 190.7 190.8 190.9 191 191.1 191.2 191.3 191.4 191.5 191.6 191.7 191.8 191.9 192 192.1 192.2 192.3 192.8 192.9 193 194 194.1 194.3 194.4 194.5 194.6 194.8 194.9 195 195.1 195.2 195.3 195.4 195.5 195.8 196 196.1 196.2 196.3 196.5 196.6 196.8 196.9 197 197.1 197.2 197.3 197.4 197.5 197.6 197.7 197.8 198 198.1 198.2 198.3 198.4 198.5 198.6 198.7 198.8 198.81 198.82 198.89 199 199.1 200 200.01 200.02 200.03 200.04 200.05 200.06 200.07 200.08 200.1 200.11 200.12 200.13 200.14 200.15 200.16 200.17 200.18 200.2 200.21 200.22 200.23 200.24 200.25 200.26 200.27 200.28 200.3 200.31 200.32 200.33 200.34 200.35 200.36 200.37 200.38 200.4 200.41 200.42 200.43 200.44 200.45 200.46 200.47 200.48 200.5 200.51 200.52 200.53 200.54 200.55 200.56 200.57 200.58 200.6 200.61 200.62 200.63 200.64 200.65 200.66 200.67 200.68 200.7 200.71 200.72 200.73 200.74 200.75 200.76 200.77 200.78 200.8 200.81 200.82 200.83 200.84 200.85 200.86 200.87 200.88 201 201.01 201.02 201.03 201.04 201.05 201.06 201.07 201.08 201.1 201.11 201.12 201.13 201.14 201.15 201.16 201.17 201.18 201.2 201.21 201.22 201.23 201.24 201.25 201.26 201.27 201.28 201.4 201.41 201.42 201.43 201.44 201.45 201.46 201.47 201.48 201.5 201.51 201.52 201.53 201.54 201.55 201.56 201.57 201.58 201.6 201.61 201.62 201.63 201.64 201.65 201.66 201.67 201.68 201.7 201.71 201.72 201.73 201.74 201.75 201.76 201.77 201.78 201.9 201.91 201.92 201.93 201.94 201.95 201.96 201.97 201.98 202 202.01 202.02 202.03 202.04 202.05 202.06 202.07 202.08 202.1 202.11 202.12 202.13 202.14 202.15 202.16 202.17 202.18 202.2 202.21 202.22 202.23 202.24 202.25 202.26 202.27 202.28 202.3 202.31 202.32 202.33 202.34 202.35 202.36 202.37 202.38 202.4 202.41 202.42 202.43 202.44 202.45 202.46 202.47 202.48 202.5 202.51 202.52 202.53 202.54 202.55 202.56 202.57 202.58 202.6 202.61 202.62 202.63 202.64 202.65 202.66 202.67 202.68 202.7 202.71 202.72 202.73 202.74 202.75 202.76 202.77 202.78 202.8 202.81 202.82 202.83 202.84 202.85 202.86 202.87 202.88 202.9 202.91 202.92 202.93 202.94 202.95 202.96 202.97 202.98 203 203.01 203.02 203.1 203.11 203.12 203.8 203.81 203.82 204 204.01 204.02 204.1 204.11 204.12 204.2 204.21 204.22 204.8 204.81 204.82 204.9 204.91 204.92 205 205.01 205.02 205.1 205.11 205.12 205.2 205.21 205.22 205.3 205.31 205.32 205.8 205.81 205.82 205.9 205.91 205.92 206 206.01 206.02 206.1 206.11 206.12 206.2 206.21 206.22 206.8 206.81 206.82 206.9 206.91 206.92 207 207.01 207.02 207.1 207.11 207.12 207.2 207.21 207.22 207.8 207.81 207.82 208 208.01 208.02 208.1 208.11 208.12 208.2 208.21 208.22 208.8 208.81 208.82 208.9 208.91 208.92

**Hypertension**: 401 401.1 401.9 402 402.01 402.1 402.11 402.9 402.91 403 403.01 403.1 403.11 403.9 403.91 404 404.01 404.02 404.03 404.1 404.11 404.12 404.13 404.9 404.91 404.92 404.93 405 405.01 405.09 405.1 405.11 405.19 405.9 405.91 405.99 437.2

**Hyperlipidemia**: 272.0 272.1 272.2 272.3 272.4

**Obesity**: 278 278.0 278.00 278.01 278.02 278.03

**Alcholism**: 291 303 535.3 571.0 571.1 571.2 572.3 980.0

**Chronic liver disease and cirrhosis**: 571.0 571.1 571.2 571.3 571.40 571.41 571.42 471.49 571.5 571.6 571.8, 571.9

**Viral hepatitis**: 070.0 070.1 070.20 070.21 070.22 070.23 070.30 070.31 070.32 070.33 070.41 070.42 070.43 070.44 070.49 070.51 070.52 070.53 070.54 070.59 070.6 070.70 070.71 070.9 573.1 573.2 V02.61

**History of acute liver injury:** 570 572.2

**Other liver disease:** 275.1 275.0 572.0 572.4 572.1 572.3 572.8 573.0 573.4 573.8 573.9

**Autoimmune diseases:** 136.1 359.71 359.79 443.1 446 555 556.8 556.7 556.8 556.9 556.0 556.1 556.2 556.3 556.4 556.5 556.6 695.4 710 714 720 725 726

**HIV infection:** 042

**Other carcinogen pathogens:** 041.86 079.4 075 795.15

**Previous irradation:** 92.29 V58.0

**COPD:** 491.0 491.1 491.2 491.8 491.9 492.0 492.8 494.0 494.1 493.0 493.1 493.2 493.8 493.9 496 506.4

**Gastrointestinal diseases:** 211.3 569.0 578.9 V12.72

**Heart failure:** 428 428 428.1 428.2 428.2 428.21 428.22 428.23 428.3 428.3 428.31 428.32 428.33 428.4 428.4 428.41 428.42 428.43 428.9 398.91 402.01 402.11 402.91 404.01 404.03 404.11 404.13 404.91 404.93

**Atrial fibrillation:** 427.31 429.4

**Stroke/transient ischemic attack:** 435 435.1 435.2 435.3 435.8 435.9 433.81 433.91 434 436 437 437.1 433.31 433.01 434.01 434.1 434.11 434.9 434.91 437.2 437.3 437.4 437.5 437.6 437.7 437.8 437.9 430 431 432 432.1 432.9

**Ischemic heart disease:** 410.01 410.02 410.1 410.11 410.12 410.2 410.21 410.22 410.3 410.31 410.32 410.4 410.41 410.42 410.5 410.51 410.52 410.6 410.61 410.62 410.7 410.71 410.72 410.8 410.81 410.82 410.9 410.91 410.92 411 411.1 411.8 411.81 411.89 413 413.1 413.9 414 414.01 414.02 414.03 414.04 414.05 414.06 414.07 414.1 414.11 414.12 414.19 414.2 414.3 414.4 414.8 414.9 410 412

**Peripheral vascular disease:** 250.7 443.9 443 443.1 443.2 443.21 443.22 443.23 443.24 443.29 443.8 443.81 443.82 443.89 441 443.9 785.4 V43.4

**Acute myocardial infarction:** 410 410.01 410.02 410.1 410.11 410.12 410.2 410.21 410.22 410.3 410.31 410.32 410.4 410.41 410.42 410.5 410.51 410.52 410.6 410.61 410.62 410.7 410.71 410.72 410.8 410.81 410.82 410.9 410.91 410.92

**Diabetic eye disease:** 362.0 362.01 250.50

**Renal diseases:** 582 582 582.1 582.2 582.4 582.8 582.81 582.89 582.9 583 583 583.1 583.2 583.4 583.6 583.7 585 585.1 585.2 585.3 585.4 585.5 585.6 585.9 586 588 588 588.1 588.8 588.81 588.89 588.9

**ICD 10 codes for cancer-related mortality**

C00 C01 C02 C03 C04 C05 C06 C07 C08 C09 C10 C11 C12 C13 C14 C15 C16 C17 C18 C19 C20 C21 C22 C23 C24 C25 C26 C27 C28 C29 C30 C31 C32 C33 C34 C35 C36 C37 C38 C39 C40 C41 C42 C43 C44 C45 C46 C47 C48 C49 C50 C51 C52 C53 C54 C55 C56 C57 C58 C59 C60 C61 C62 C63 C64 C65 C66 C67 C68 C69 C70 C71 C72 C73 C74 C75 C76 C77 C78 C79 C80 C81 C82 C83 C84 C85 C86 C87 C88 C89 C90 C91 C92 C93 C94 C95 C96 C97

**Supplementary Table 2. Calculations for SD variability measure**

| **Variability measure** | **Definition** |
| --- | --- |
| Standard deviation | $\sqrt{\frac{1}{Number of measurements}\sum_{i=1}^{Number of measurements} {({test}_{i}-individual mean)}^{2}}$ |

.

**Supplementary Table 3A. Baseline and clinical characteristics of patients with adverse cancer and mortality before propensity score matching.**

* for SMD$\geq$0.2; SD: standard deviation; SGLT2I: sodium glucose cotransporter-2 inhibitor; DPP4I: dipeptidyl peptidase-4 inhibitor.

| **Characteristics** | **All (N=60112) Mean(SD);N or Count(%)** | **All-cause mortality (N=3033) Mean(SD);N or Count(%)** | **Cancer related mortality (N=506) Mean(SD);N or Count(%)** | **New onset lung cancer (N=249) Mean(SD);N or Count(%)** | **New onset gastrointestinal cancer (N=817) Mean(SD);N or Count(%)** | **New onset breast cancer (N=201) Mean(SD);N or Count(%)** | **New onset genitourinary cancer (N=261) Mean(SD);N or Count(%)** | **New onset bladder cancer (N=97) Mean(SD);N or Count(%)** |
| --- | --- | --- | --- | --- | --- | --- | --- | --- |
| Demographics |  |  |  |  |  |  |  |  |
| Male gender | 33883(56.36%) | 1822(60.07%) | 307(60.67%) | 170(68.27%) | 530(64.87%) | 6(2.98%) | 139(53.25%) | 83(85.56%) |
| Female gender | 26229(43.63%) | 1211(39.92%) | 199(39.32%) | 79(31.72%) | 287(35.12%) | 195(97.01%) | 122(46.74%) | 14(14.43%) |
| Baseline age, years | 62.1(12.4);n=60112 | 73.0(11.9);n=3033 | 70.5(10.8);n=506 | 68.1(9.4);n=249 | 68.1(10.0);n=817 | 64.7(11.2);n=201 | 66.9(12.2);n=261 | 71.1(11.6);n=97 |
| 18-50 | 9057(15.06%) | 109(3.59%) | 15(2.96%) | 4(1.60%) | 29(3.54%) | 17(8.45%) | 21(8.04%) | 5(5.15%) |
| 50-60 | 17550(29.19%) | 349(11.50%) | 76(15.01%) | 48(19.27%) | 129(15.78%) | 52(25.87%) | 59(22.60%) | 12(12.37%) |
| 60-70 | 18029(29.99%) | 679(22.38%) | 138(27.27%) | 93(37.34%) | 320(39.16%) | 64(31.84%) | 72(27.58%) | 25(25.77%) |
| 70-80 | 10338(17.19%) | 904(29.80%) | 176(34.78%) | 76(30.52%) | 226(27.66%) | 48(23.88%) | 62(23.75%) | 31(31.95%) |
| >80 | 5145(8.55%) | 992(32.70%) | 101(19.96%) | 28(11.24%) | 113(13.83%) | 20(9.95%) | 47(18.00%) | 24(24.74%) |
| Past comorbidities |  |  |  |  |  |  |  |  |
| Charlson standard comorbidity index | 1.9(1.4);n=60112 | 3.2(1.6);n=3033 | 2.8(1.4);n=506 | 2.6(1.2);n=249 | 2.5(1.2);n=817 | 2.2(1.2);n=201 | 2.4(1.4);n=261 | 2.8(1.3);n=97 |
| Duration from earliest diabetes mellitus date to baseline date, day | 619.9(1341.9);n=60112 | 465.9(1219.0);n=3033 | 312.1(978.5);n=506 | 543.4(1316.1);n=249 | 503.6(1261.1);n=817 | 626.6(1344.9);n=201 | 512.7(1245.9);n=261 | 413.1(985.0);n=97 |
| Hypertension | 13376(22.25%) | 964(31.78%) | 120(23.71%) | 59(23.69%) | 203(24.84%) | 56(27.86%) | 67(25.67%) | 24(24.74%) |
| Hyperlipidaemia | 1620(2.69%) | 60(1.97%) | 10(1.97%) | 2(0.80%) | 10(1.22%) | 5(2.48%) | 6(2.29%) | 2(2.06%) |
| Hypotension | 350(0.58%) | 47(1.54%) | 2(0.39%) | 3(1.20%) | 5(0.61%) | 1(0.49%) | 1(0.38%) | 1(1.03%) |
| Overweight, obesity and hyperalimentation | 432(0.71%) | 5(0.16%) | 1(0.19%) | 0(0.00%) | 2(0.24%) | 3(1.49%) | 2(0.76%) | 1(1.03%) |
| Gout | 1510(2.51%) | 143(4.71%) | 14(2.76%) | 7(2.81%) | 25(3.05%) | 4(1.99%) | 11(4.21%) | 5(5.15%) |
| Heart failure | 1629(2.70%) | 253(8.34%) | 14(2.76%) | 7(2.81%) | 21(2.57%) | 5(2.48%) | 10(3.83%) | 3(3.09%) |
| Acute myocardial infarction | 1521(2.53%) | 134(4.41%) | 14(2.76%) | 4(1.60%) | 23(2.81%) | 3(1.49%) | 11(4.21%) | 6(6.18%) |
| Ischemic heart disease | 5680(9.44%) | 362(11.93%) | 44(8.69%) | 23(9.23%) | 80(9.79%) | 14(6.96%) | 29(11.11%) | 15(15.46%) |
| Peripheral vascular disease | 375(0.62%) | 68(2.24%) | 5(0.98%) | 3(1.20%) | 1(0.12%) | 1(0.49%) | 2(0.76%) | 2(2.06%) |
| Stroke/transient ischemic attack | 1777(2.95%) | 184(6.06%) | 18(3.55%) | 12(4.81%) | 23(2.81%) | 6(2.98%) | 7(2.68%) | 5(5.15%) |
| Atrial fibrillation | 1325(2.20%) | 167(5.50%) | 12(2.37%) | 4(1.60%) | 17(2.08%) | 6(2.98%) | 8(3.06%) | 4(4.12%) |
| Diabetic eye disease | 4045(6.72%) | 330(10.88%) | 32(6.32%) | 13(5.22%) | 46(5.63%) | 13(6.46%) | 19(7.27%) | 7(7.21%) |
| Alcohol dependence | 119(0.19%) | 29(0.95%) | 4(0.79%) | 0(0.00%) | 11(1.34%) | 0(0.00%) | 0(0.00%) | 0(0.00%) |
| Chronic liver disease and cirrhosis | 1195(1.98%) | 57(1.87%) | 6(1.18%) | 3(1.20%) | 22(2.69%) | 2(0.99%) | 5(1.91%) | 2(2.06%) |
| Viral hepatitis | 629(1.04%) | 32(1.05%) | 10(1.97%) | 2(0.80%) | 26(3.18%) | 0(0.00%) | 2(0.76%) | 1(1.03%) |
| History of acute liver injury | 159(0.26%) | 24(0.79%) | 2(0.39%) | 0(0.00%) | 13(1.59%) | 1(0.49%) | 0(0.00%) | 0(0.00%) |
| Other liver disease | 612(1.01%) | 51(1.68%) | 2(0.39%) | 2(0.80%) | 18(2.20%) | 3(1.49%) | 0(0.00%) | 0(0.00%) |
| Autoimmune disease tissue | 620(1.03%) | 22(0.72%) | 2(0.39%) | 2(0.80%) | 10(1.22%) | 2(0.99%) | 0(0.00%) | 0(0.00%) |
| Other carcinogens pathogen | 13(0.02%) | 1(0.03%) | 0(0.00%) | 1(0.40%) | 0(0.00%) | 0(0.00%) | 0(0.00%) | 0(0.00%) |
| Chronic obstructive pulmonary disease | 69(0.11%) | 8(0.26%) | 0(0.00%) | 1(0.40%) | 0(0.00%) | 0(0.00%) | 0(0.00%) | 0(0.00%) |
| Gastrointestinal disease | 1353(2.25%) | 95(3.13%) | 8(1.58%) | 5(2.00%) | 18(2.20%) | 7(3.48%) | 10(3.83%) | 6(6.18%) |
| Medications |  |  |  |  |  |  |  |  |
| SGLT2I v.s. DPP4I | 18167(30.22%) | 492(16.22%) | 115(22.72%) | 48(19.27%) | 151(18.48%) | 44(21.89%) | 57(21.83%) | 24(24.74%) |
| SGLT2I frequency | 7.4(10.1);n=18167 | 10.9(15.9);n=492 | 13.4(17.6);n=115 | 9.9(11.3);n=48 | 10.9(15.5);n=151 | 10.0(12.6);n=44 | 11.9(14.6);n=57 | 11.3(15.4);n=24 |
| DPP4I frequency | 5.5(7.2);n=41945 | 5.1(14.8);n=2541 | 5.8(8.8);n=391 | 6.5(8.8);n=201 | 6.4(9.6);n=666 | 6.2(8.1);n=157 | 4.9(6.3);n=204 | 4.3(6.5);n=73 |
| SGLT2I duration, days | 538.1(674.5);n=18167 | 444.8(572.2);n=492 | 573.6(655.8);n=115 | 573.1(689.1);n=48 | 540.0(657.0);n=151 | 588.2(629.0);n=44 | 548.8(674.7);n=57 | 514.2(663.4);n=24 |
| DPP4I duration, days | 528.9(301.0);n=41945 | 428.6(298.7);n=2541 | 419.6(309.8);n=391 | 469.6(313.5);n=201 | 453.9(300.1);n=666 | 534.4(304.8);n=157 | 486.3(294.0);n=204 | 510.3(292.4);n=73 |
| Metformin | 54407(90.50%) | 2227(73.42%) | 432(85.37%) | 222(89.15%) | 721(88.24%) | 180(89.55%) | 226(86.59%) | 82(84.53%) |
| Sulphonylurea | 46521(77.39%) | 2369(78.10%) | 386(76.28%) | 205(82.32%) | 634(77.60%) | 156(77.61%) | 204(78.16%) | 75(77.31%) |
| Insulin | 29349(48.82%) | 2489(82.06%) | 377(74.50%) | 159(63.85%) | 614(75.15%) | 103(51.24%) | 173(66.28%) | 65(67.01%) |
| Acarbose | 1566(2.60%) | 92(3.03%) | 15(2.96%) | 6(2.40%) | 23(2.81%) | 7(3.48%) | 10(3.83%) | 4(4.12%) |
| Thiozolidinedone | 12046(20.03%) | 287(9.46%) | 69(13.63%) | 38(15.26%) | 107(13.09%) | 30(14.92%) | 35(13.40%) | 14(14.43%) |
| Glucagon-like peptide-1 receptor agonists | 2044(3.40%) | 20(0.65%) | 4(0.79%) | 2(0.80%) | 7(0.85%) | 3(1.49%) | 5(1.91%) | 3(3.09%) |
| ACEI/ARB | 18249(30.35%) | 773(25.48%) | 128(25.29%) | 75(30.12%) | 227(27.78%) | 59(29.35%) | 86(32.95%) | 34(35.05%) |
| Antidepressants | 2742(4.56%) | 230(7.58%) | 47(9.28%) | 18(7.22%) | 45(5.50%) | 13(6.46%) | 15(5.74%) | 2(2.06%) |
| Antihypertensive drugs | 2311(3.84%) | 110(3.62%) | 18(3.55%) | 6(2.40%) | 26(3.18%) | 4(1.99%) | 9(3.44%) | 6(6.18%) |
| Antihepatitis | 809(1.34%) | 41(1.35%) | 13(2.56%) | 4(1.60%) | 58(7.09%) | 2(0.99%) | 3(1.14%) | 2(2.06%) |
| Anticoagulants | 29401(48.91%) | 1286(42.40%) | 239(47.23%) | 119(47.79%) | 354(43.32%) | 84(41.79%) | 120(45.97%) | 44(45.36%) |
| Antiplatelets | 10115(16.82%) | 626(20.63%) | 77(15.21%) | 49(19.67%) | 126(15.42%) | 22(10.94%) | 42(16.09%) | 18(18.55%) |
| Statins and fibrates | 34307(57.07%) | 1281(42.23%) | 262(51.77%) | 140(56.22%) | 432(52.87%) | 111(55.22%) | 142(54.40%) | 48(49.48%) |
| Nitrates | 4652(7.73%) | 322(10.61%) | 38(7.50%) | 21(8.43%) | 68(8.32%) | 10(4.97%) | 24(9.19%) | 9(9.27%) |
| Non-steroidal anti-inflammatory drugs | 9719(16.16%) | 595(19.61%) | 73(14.42%) | 48(19.27%) | 119(14.56%) | 18(8.95%) | 41(15.70%) | 17(17.52%) |
| Diuretics | 10179(16.93%) | 785(25.88%) | 114(22.52%) | 47(18.87%) | 171(20.93%) | 33(16.41%) | 65(24.90%) | 22(22.68%) |
| Beta-blockers | 8075(13.43%) | 538(17.73%) | 70(13.83%) | 36(14.45%) | 103(12.60%) | 25(12.43%) | 42(16.09%) | 13(13.40%) |
| Calcium channel blockers | 14154(23.54%) | 787(25.94%) | 123(24.30%) | 62(24.89%) | 197(24.11%) | 51(25.37%) | 77(29.50%) | 27(27.83%) |
| SUbclinical biomarkers |  |  |  |  |  |  |  |  |
| Abbreviated MDRD, mL/min/1.73m^2 | 81.4(27.9);n=49633 | 59.9(28.9);n=2450 | 72.9(24.6);n=389 | 78.3(24.7);n=195 | 75.1(25.1);n=671 | 78.7(27.4);n=171 | 71.8(27.1);n=211 | 70.3(24.0);n=82 |
| Most severe renal damage (<15 mL/min/1.73m^2) | 436.0(0.72%) | 129.0(4.25%) | 0.0(0.00%) | 1.0(0.40%) | 6.0(0.73%) | 3.0(1.49%) | 4.0(1.53%) | 0.0(0.00%) |
| Severe renal damage ([15, 30) mL/min/1.73m^2) | 1108.0(1.84%) | 235.0(7.74%) | 13.0(2.56%) | 5.0(2.00%) | 15.0(1.83%) | 2.0(0.99%) | 9.0(3.44%) | 3.0(3.09%) |
| Moderate to severe renal damage ([30, 45) mL/min/1.73m^2) | 3548.0(5.90%) | 454.0(14.96%) | 45.0(8.89%) | 14.0(5.62%) | 60.0(7.34%) | 15.0(7.46%) | 17.0(6.51%) | 6.0(6.18%) |
| Mild to moderate renal damage ([45, 60) mL/min/1.73m^2) | 5816.0(9.67%) | 458.0(15.10%) | 63.0(12.45%) | 25.0(10.04%) | 111.0(13.58%) | 24.0(11.94%) | 38.0(14.55%) | 22.0(22.68%) |
| Mild renal damage ([60, 90] mL/min/1.73m^2) | 19920.0(33.13%) | 801.0(26.40%) | 176.0(34.78%) | 84.0(33.73%) | 297.0(36.35%) | 68.0(33.83%) | 97.0(37.16%) | 35.0(36.08%) |
| Chronic kidney disease (>90 mL/min/1.73m^2) | 18805.0(31.28%) | 373.0(12.29%) | 92.0(18.18%) | 66.0(26.50%) | 182.0(22.27%) | 59.0(29.35%) | 46.0(17.62%) | 16.0(16.49%) |
| Neutrophil-to-lymphocyte ratio | 3.4(4.5);n=23978 | 5.2(7.2);n=1590 | 4.3(6.6);n=187 | 3.3(4.2);n=81 | 4.3(7.7);n=316 | 3.2(2.9);n=90 | 4.0(6.6);n=95 | 3.8(3.7);n=37 |
| Platelet-to-lymphocyte ratio | 142.0(148.2);n=23976 | 170.5(139.2);n=1589 | 148.6(98.0);n=187 | 134.3(61.9);n=81 | 144.0(196.8);n=316 | 140.7(75.7);n=90 | 161.1(122.9);n=95 | 161.6(94.3);n=37 |
| Neutrophil-to-high-density lipoprotein ratio | 0.3(0.2);n=21968 | 0.31(0.24);n=1311 | 0.28(0.19);n=160 | 0.26(0.15);n=76 | 0.31(0.56);n=277 | 0.2(0.1);n=75 | 0.27(0.14);n=87 | 0.26(0.13);n=34 |
| Low density lipoprotein ratio-to-high density lipoprotein ratio | 2.1(0.9);n=46116 | 2.14(1.01);n=2073 | 2.0(0.8);n=354 | 2.0(0.8);n=183 | 2.06(0.86);n=606 | 2.06(0.8);n=152 | 2.06(0.88);n=201 | 2.0(0.9);n=77 |
| Triglyceride-glucose index | 7.6(0.7);n=42025 | 7.5(0.8);n=1881 | 7.4(0.6);n=311 | 7.5(0.6);n=160 | 7.4(0.7);n=539 | 7.64(0.71);n=133 | 7.5(0.7);n=172 | 7.4(0.7);n=65 |
| Protein-to-creatinine ratio | 3.0(1.5);n=29872 | 2.2(1.4);n=1833 | 2.7(1.5);n=228 | 3.0(1.42);n=109 | 2.5(1.3);n=393 | 3.5(1.5);n=102 | 2.8(1.4);n=119 | 2.6(1.4);n=46 |
| Aspartate aminotransferase-to-alanine transaminase ratio | 1.1(3.2);n=8773 | 1.3(1.0);n=503 | 1.2(0.8);n=62 | 1.07(0.55);n=39 | 1.2(0.9);n=121 | 1.0(0.4);n=26 | 1.07(0.48);n=43 | 1.08(0.37);n=17 |
| Complete blood counts, renal and liver functions |  |  |  |  |  |  |  |  |
| Mean corpuscular volume, fL | 87.1(7.5);n=29998 | 88.2(7.7);n=1841 | 88.5(6.7);n=231 | 88.7(7.1);n=110 | 88.4(7.6);n=394 | 87.0(8.0);n=102 | 86.9(7.6);n=119 | 86.5(8.4);n=46 |
| Eosinophil, x10^9/L | 0.2(0.3);n=23959 | 0.22(0.24);n=1585 | 0.21(0.19);n=186 | 0.3(0.9);n=81 | 0.22(0.28);n=316 | 0.2(0.43);n=90 | 0.3(0.3);n=95 | 0.3(0.3);n=37 |
| Lymphocyte, x10^9/L | 2.0(0.9);n=23981 | 1.7(1.2);n=1590 | 1.9(1.0);n=187 | 2.01(0.71);n=81 | 1.9(0.9);n=316 | 2.04(0.74);n=90 | 1.9(0.8);n=95 | 1.8(0.8);n=37 |
| Neutrophil, x10^9/L | 5.3(2.8);n=23981 | 6.0(3.5);n=1590 | 5.5(2.9);n=187 | 5.2(2.4);n=81 | 5.2(2.7);n=316 | 5.26(2.58);n=90 | 5.25(2.22);n=95 | 5.0(2.1);n=37 |
| White cell count, x10^9/L | 8.0(3.0);n=30008 | 8.4(3.4);n=1841 | 8.1(2.6);n=231 | 8.2(2.5);n=110 | 7.8(2.8);n=395 | 7.9(2.7);n=102 | 8.3(2.4);n=119 | 8.2(2.5);n=46 |
| Platelet, x10^9/L | 241.3(71.0);n=30006 | 228.0(79.1);n=1840 | 229.2(80.7);n=231 | 244.2(65.1);n=110 | 214.5(78.4);n=395 | 250.6(69.4);n=102 | 244.7(65.3);n=119 | 234.7(71.1);n=46 |
| Red cell count, x10^12/L | 4.6(0.7);n=29998 | 4.2(0.8);n=1841 | 4.4(0.6);n=231 | 4.5(0.6);n=110 | 4.4(0.6);n=394 | 4.3(0.7);n=102 | 4.4(0.8);n=119 | 4.5(0.8);n=46 |
| Potassium, mmol/L | 4.3(0.5);n=49471 | 4.4(0.6);n=2445 | 4.34(0.46);n=388 | 4.5(0.5);n=195 | 4.35(0.5);n=669 | 4.3(0.5);n=169 | 4.4(0.5);n=211 | 4.4(0.5);n=82 |
| Albumin, g/L | 41.9(3.8);n=37720 | 38.7(4.8);n=2011 | 40.8(3.9);n=276 | 42.2(3.8);n=141 | 41.1(4.0);n=499 | 42.0(3.2);n=125 | 41.1(4.0);n=152 | 40.7(4.1);n=57 |
| Sodium, mmol/L | 139.3(2.9);n=49494 | 138.8(3.9);n=2447 | 139.4(3.2);n=388 | 139.5(2.8);n=195 | 139.31(2.95);n=669 | 139.7(3.1);n=169 | 139.4(3.0);n=211 | 139.4(3.0);n=82 |
| Urea, mmol/L | 6.4(3.3);n=49483 | 9.0(6.0);n=2445 | 6.8(3.0);n=387 | 6.5(2.9);n=193 | 6.7(3.2);n=669 | 6.8(4.8);n=171 | 7.2(4.2);n=211 | 7.2(2.5);n=82 |
| Protein, g/L | 73.9(5.4);n=35490 | 72.5(6.9);n=1896 | 73.91(5.66);n=263 | 74.4(6.2);n=131 | 74.0(5.2);n=471 | 74.8(5.8);n=119 | 73.2(6.4);n=142 | 73.2(6.3);n=56 |
| Creatinine, umol/L | 92.5(71.5);n=49633 | 144.7(156.8);n=2450 | 96.0(36.9);n=389 | 92.49(42.54);n=195 | 98.5(68.1);n=671 | 87.1(96.8);n=171 | 107.6(107.3);n=211 | 102.7(32.7);n=82 |
| Alkaline phosphatase, U/L | 76.2(30.1);n=37836 | 87.8(44.0);n=2017 | 81.8(46.9);n=278 | 75.9(23.4);n=144 | 78.8(29.3);n=501 | 84.5(51.3);n=125 | 74.3(22.0);n=151 | 73.9(23.6);n=57 |
| Aspartate transaminase, U/L | 28.2(53.4);n=15006 | 29.2(57.3);n=824 | 34.8(76.0);n=95 | 24.0(12.5);n=58 | 40.1(79.7);n=214 | 25.1(13.7);n=42 | 21.8(14.2);n=66 | 20.8(10.8);n=28 |
| Alanine transaminase, U/L | 29.4(33.6);n=32037 | 24.9(37.6);n=1692 | 27.9(32.5);n=241 | 26.1(17.1);n=123 | 30.8(32.9);n=403 | 26.8(19.7);n=108 | 21.9(13.1);n=122 | 20.2(9.5);n=44 |
| Bilirubin, umol/L | 11.3(6.7);n=37645 | 11.1(8.4);n=2008 | 11.8(10.3);n=276 | 9.8(4.3);n=142 | 12.6(9.6);n=498 | 9.7(4.0);n=125 | 10.5(6.1);n=151 | 11.0(8.0);n=57 |
| Lipid profiles |  |  |  |  |  |  |  |  |
| Triglyceride, mmol/L | 1.7(1.6);n=46943 | 1.6(1.2);n=2104 | 1.4(0.8);n=355 | 1.5(0.9);n=185 | 1.5(1.0);n=610 | 1.8(1.0);n=153 | 1.6(1.2);n=201 | 1.4(0.8);n=77 |
| SD of triglyceride | 0.5(1.0);n=24256 | 0.4(0.6);n=1044 | 0.3(0.3);n=153 | 0.4(0.6);n=83 | 0.3(0.4);n=300 | 0.4(0.5);n=72 | 0.4(0.9);n=92 | 0.3(0.3);n=32 |
| Low-density lipoprotein, mmol/L | 2.4(0.8);n=46121 | 2.38(0.9);n=2075 | 2.3(0.8);n=355 | 2.2(0.7);n=184 | 2.3(0.7);n=606 | 2.44(0.86);n=152 | 2.3(0.8);n=201 | 2.1(0.7);n=77 |
| SD of low-density lipoprotein | 0.4(0.3);n=23541 | 0.4(0.39);n=1020 | 0.36(0.37);n=152 | 0.3(0.3);n=82 | 0.3(0.3);n=299 | 0.39(0.38);n=69 | 0.36(0.37);n=93 | 0.2(0.2);n=32 |
| High-density lipoprotein, mmol/L | 1.2(0.3);n=46872 | 1.2(0.36);n=2102 | 1.2(0.34);n=355 | 1.21(0.32);n=185 | 1.19(0.33);n=610 | 1.24(0.3);n=153 | 1.2(0.38);n=201 | 1.18(0.45);n=77 |
| SD of high-density lipoprotein | 0.1(0.1);n=23320 | 0.12(0.1);n=1015 | 0.11(0.09);n=146 | 0.12(0.11);n=80 | 0.1(0.08);n=283 | 0.1(0.07);n=68 | 0.11(0.09);n=91 | 0.11(0.08);n=31 |
| Total cholesterol, mmol/L | 4.3(1.0);n=46986 | 4.3(1.12);n=2107 | 4.1(0.9);n=355 | 4.1(0.9);n=185 | 4.2(0.9);n=610 | 4.5(1.1);n=153 | 4.2(1.0);n=201 | 3.9(0.8);n=77 |
| SD of total cholesterol | 0.4(0.4);n=24282 | 0.5(0.5);n=1042 | 0.41(0.42);n=153 | 0.42(0.38);n=84 | 0.39(0.34);n=299 | 0.5(0.4);n=73 | 0.5(0.4);n=92 | 0.3(0.2);n=32 |
| Hemoglobin A1C, % | 8.1(1.5);n=48920 | 8.0(1.9);n=2316 | 7.9(1.4);n=375 | 7.9(1.4);n=195 | 7.9(1.3);n=644 | 8.06(1.54);n=165 | 7.8(1.5);n=204 | 7.8(1.6);n=80 |
| SD of hemoglobin A1C | 0.6(0.7);n=34629 | 0.7(1.0);n=1517 | 0.59(0.61);n=237 | 0.5(0.5);n=126 | 0.5(0.5);n=422 | 0.58(0.58);n=116 | 0.59(0.65);n=133 | 0.61(0.72);n=53 |
| Fasting glucose, mmol/L | 9.9(3.3);n=34323 | 10.8(5.5);n=1504 | 9.7(3.2);n=246 | 9.2(2.2);n=123 | 9.7(3.2);n=423 | 10.0(3.0);n=117 | 9.8(2.8);n=123 | 9.6(2.4);n=45 |
| SD of fasting glucose | 1.9(2.1);n=27210 | 2.7(2.7);n=1435 | 2.2(2.0);n=209 | 1.8(1.7);n=101 | 1.93(1.93);n=363 | 2.2(2.5);n=89 | 2.1(2.1);n=100 | 2.4(2.3);n=36 |
| SGLT2I v.s. DPP4I.1 | 18167(30.22%) | 492(16.22%) | 115(22.72%) | 48(19.27%) | 151(18.48%) | 44(21.89%) | 57(21.83%) | 24(24.74%) |
| Dapagliflozin | 10556(17.56%) | 294(9.69%) | 72(14.22%) | 32(12.85%) | 86(10.52%) | 21(10.44%) | 37(14.17%) | 16(16.49%) |
| Empagliflozin | 3780(6.28%) | 85(2.80%) | 21(4.15%) | 8(3.21%) | 30(3.67%) | 8(3.98%) | 12(4.59%) | 4(4.12%) |
| Canagliflozin | 4523(7.52%) | 117(3.85%) | 24(4.74%) | 8(3.21%) | 37(4.52%) | 14(6.96%) | 10(3.83%) | 4(4.12%) |
| Ertugliflozin | 2527(4.20%) | 73(2.40%) | 12(2.37%) | 5(2.00%) | 18(2.20%) | 7(3.48%) | 3(1.14%) | 1(1.03%) |

**Supplementary Table 3B. Baseline and clinical characteristics of patients with adverse cancer and mortality after propensity score matching (1:1).**

* for SMD$\geq$0.2; SD: standard deviation; SGLT2I: sodium glucose cotransporter-2 inhibitor; DPP4I: dipeptidyl peptidase-4 inhibitor.

| **Characteristics** | **All (N=36334) Mean(SD);N or Count(%)** | **All-cause mortality (N=970) Mean(SD);N or Count(%)** | **Cancer related mortality (N=211) Mean(SD);N or Count(%)** | **New onset cancer (N=674) Mean(SD);N or Count(%)** | **New onset lung cancer (N=124) Mean(SD);N or Count(%)** | **New onset gastrointestinal cancer (N=325) Mean(SD);N or Count(%)** | **New onset breast cancer (N=101) Mean(SD);N or Count(%)** | **New onset genitourinary cancer (N=121) Mean(SD);N or Count(%)** | **New onset bladder cancer (N=50) Mean(SD);N or Count(%)** |
| --- | --- | --- | --- | --- | --- | --- | --- | --- | --- |
| Demographics |  |  |  |  |  |  |  |  |  |
| Male gender | 22288(61.34%) | 636(65.56%) | 120(56.87%) | 383(56.82%) | 93(74.99%) | 200(61.53%) | 18(17.82%) | 71(58.67%) | 44(87.99%) |
| Female gender | 14046(38.65%) | 334(34.43%) | 91(43.12%) | 291(43.17%) | 31(24.99%) | 125(38.46%) | 83(82.17%) | 50(41.32%) | 6(11.99%) |
| Baseline age, years | 58.2(11.0);n=36334 | 68.3(11.0);n=970 | 66.9(9.5);n=211 | 63.9(10.2);n=674 | 65.6(8.4);n=124 | 65.4(9.4);n=325 | 61.0(8.5);n=101 | 60.7(13.1);n=121 | 60.6(16.6);n=50 |
| 18-50 | 7212(19.84%) | 46(4.74%) | 8(3.79%) | 47(6.97%) | 2(1.61%) | 22(6.76%) | 5(4.95%) | 16(13.22%) | 12(23.99%) |
| 50-60 | 13285(36.56%) | 165(17.01%) | 43(20.37%) | 176(26.11%) | 36(29.03%) | 54(16.61%) | 50(49.50%) | 37(30.57%) | 5(9.99%) |
| 60-70 | 11194(30.80%) | 335(34.53%) | 82(38.86%) | 278(41.24%) | 52(41.93%) | 152(46.76%) | 30(29.70%) | 43(35.53%) | 19(37.99%) |
| 70-80 | 3766(10.36%) | 276(28.45%) | 57(27.01%) | 136(20.17%) | 26(20.96%) | 76(23.38%) | 15(14.85%) | 18(14.87%) | 10(19.99%) |
| >80 | 881(2.42%) | 148(15.25%) | 21(9.95%) | 37(5.48%) | 8(6.45%) | 21(6.46%) | 1(0.99%) | 7(5.78%) | 4(7.99%) |
| Past comorbidities |  |  |  |  |  |  |  |  |  |
| Charlson standard comorbidity index | 1.5(1.2);n=36334 | 2.7(1.5);n=970 | 2.4(1.3);n=211 | 2.1(1.2);n=674 | 2.3(1.2);n=124 | 2.3(1.2);n=325 | 1.7(0.9);n=101 | 1.8(1.2);n=121 | 1.8(1.3);n=50 |
| Duration from earliest diabetes mellitus date to baseline date, day | 590.9(1323.1);n=36334 | 403.8(1109.1);n=970 | 263.2(901.1);n=211 | 386.4(1033.8);n=674 | 368.8(1075.5);n=124 | 389.1(1013.9);n=325 | 421.3(1182.6);n=101 | 353.4(899.2);n=121 | 231.7(672.1);n=50 |
| Hypertension | 8325(22.91%) | 305(31.44%) | 52(24.64%) | 164(24.33%) | 24(19.35%) | 87(26.76%) | 24(23.76%) | 29(23.96%) | 9(17.99%) |
| Hyperlipidaemia | 1265(3.48%) | 16(1.64%) | 8(3.79%) | 17(2.52%) | 0(0.00%) | 10(3.07%) | 3(2.97%) | 3(2.47%) | 1(1.99%) |
| Hypotension | 152(0.41%) | 12(1.23%) | 1(0.47%) | 5(0.74%) | 1(0.80%) | 1(0.30%) | 0(0.00%) | 2(1.65%) | 2(3.99%) |
| Overweight, obesity and hyperalimentation | 576(1.58%) | 5(0.51%) | 1(0.47%) | 8(1.18%) | 0(0.00%) | 4(1.23%) | 1(0.99%) | 2(1.65%) | 1(1.99%) |
| Gout | 782(2.15%) | 37(3.81%) | 7(3.31%) | 17(2.52%) | 3(2.41%) | 10(3.07%) | 1(0.99%) | 3(2.47%) | 1(1.99%) |
| Heart failure | 874(2.40%) | 86(8.86%) | 4(1.89%) | 16(2.37%) | 2(1.61%) | 8(2.46%) | 2(1.98%) | 3(2.47%) | 1(1.99%) |
| Acute myocardial infarction | 1210(3.33%) | 70(7.21%) | 7(3.31%) | 18(2.67%) | 2(1.61%) | 13(3.99%) | 0(0.00%) | 4(3.30%) | 2(3.99%) |
| Ischemic heart disease | 4466(12.29%) | 167(17.21%) | 26(12.32%) | 77(11.42%) | 17(13.70%) | 46(14.15%) | 4(3.96%) | 11(9.09%) | 5(9.99%) |
| Peripheral vascular disease | 194(0.53%) | 29(2.98%) | 1(0.47%) | 2(0.29%) | 0(0.00%) | 1(0.30%) | 0(0.00%) | 0(0.00%) | 0(0.00%) |
| Stroke/transient ischemic attack | 940(2.58%) | 66(6.80%) | 4(1.89%) | 19(2.81%) | 4(3.22%) | 12(3.69%) | 1(0.99%) | 1(0.82%) | 0(0.00%) |
| Atrial fibrillation | 766(2.10%) | 59(6.08%) | 6(2.84%) | 19(2.81%) | 1(0.80%) | 10(3.07%) | 5(4.95%) | 2(1.65%) | 2(3.99%) |
| Diabetic eye disease | 2459(6.76%) | 88(9.07%) | 19(9.00%) | 42(6.23%) | 4(3.22%) | 27(8.30%) | 2(1.98%) | 8(6.61%) | 4(7.99%) |
| Alcohol dependence | 44(0.12%) | 8(0.82%) | 3(1.42%) | 4(0.59%) | 0(0.00%) | 4(1.23%) | 0(0.00%) | 0(0.00%) | 0(0.00%) |
| Chronic liver disease and cirrhosis | 995(2.73%) | 25(2.57%) | 4(1.89%) | 22(3.26%) | 1(0.80%) | 17(5.23%) | 1(0.99%) | 3(2.47%) | 1(1.99%) |
| Viral hepatitis | 414(1.13%) | 6(0.61%) | 0(0.00%) | 11(1.63%) | 1(0.80%) | 10(3.07%) | 0(0.00%) | 0(0.00%) | 0(0.00%) |
| History of acute liver injury | 72(0.19%) | 5(0.51%) | 0(0.00%) | 5(0.74%) | 0(0.00%) | 5(1.53%) | 0(0.00%) | 0(0.00%) | 0(0.00%) |
| Other liver disease | 324(0.89%) | 14(1.44%) | 2(0.94%) | 15(2.22%) | 1(0.80%) | 12(3.69%) | 1(0.99%) | 0(0.00%) | 0(0.00%) |
| Autoimmune disease tissue | 386(1.06%) | 12(1.23%) | 2(0.94%) | 6(0.89%) | 1(0.80%) | 4(1.23%) | 1(0.99%) | 0(0.00%) | 0(0.00%) |
| Other carcinogens pathogen | 12(0.03%) | 0(0.00%) | 0(0.00%) | 5(0.74%) | 5(4.03%) | 0(0.00%) | 0(0.00%) | 0(0.00%) | 0(0.00%) |
| Chronic obstructive pulmonary disease | 24(0.06%) | 5(0.51%) | 0(0.00%) | 4(0.59%) | 4(3.22%) | 0(0.00%) | 0(0.00%) | 0(0.00%) | 0(0.00%) |
| Gastrointestinal disease | 690(1.89%) | 33(3.40%) | 4(1.89%) | 22(3.26%) | 7(5.64%) | 8(2.46%) | 2(1.98%) | 5(4.13%) | 4(7.99%) |
| Medications |  |  |  |  |  |  |  |  |  |
| SGLT2I v.s. DPP4I | 18167(50.00%) | 431(44.43%) | 74(35.07%) | 242(35.90%) | 42(33.87%) | 129(39.69%) | 32(31.68%) | 39(32.23%) | 11(21.99%) |
| SGLT2I frequency | 7.3(10.0);n=18167 | 10.5(15.0);n=431 | 12.5(14.7);n=74 | 10.6(13.6);n=242 | 9.7(11.7);n=42 | 10.9(15.2);n=129 | 9.9(11.2);n=32 | 11.9(13.6);n=39 | 10.6(9.9);n=11 |
| DPP4I frequency | 6.3(8.0);n=18167 | 7.0(13.7);n=539 | 7.2(13.9);n=137 | 7.5(10.9);n=432 | 5.0(7.0);n=82 | 7.8(11.5);n=196 | 11.3(12.8);n=69 | 5.5(9.2);n=82 | 5.8(11.8);n=39 |
| SGLT2I duration, days | 538.5(673.6);n=18167 | 433.6(562.3);n=431 | 534.0(650.9);n=74 | 580.3(666.1);n=242 | 548.5(702.2);n=42 | 554.7(666.5);n=129 | 617.4(602.0);n=32 | 632.0(705.1);n=39 | 725.8(712.3);n=11 |
| DPP4I duration, days | 575.3(360.3);n=18167 | 448.7(374.4);n=539 | 489.9(464.7);n=137 | 498.8(394.6);n=432 | 516.7(356.1);n=82 | 481.2(393.9);n=196 | 638.0(397.6);n=69 | 412.3(393.8);n=82 | 302.5(419.2);n=39 |
| Metformin | 33945(93.42%) | 834(85.97%) | 197(93.36%) | 612(90.80%) | 113(91.12%) | 295(90.76%) | 95(94.05%) | 106(87.60%) | 44(87.99%) |
| Sulphonylurea | 26376(72.59%) | 743(76.59%) | 168(79.62%) | 511(75.81%) | 104(83.87%) | 249(76.61%) | 80(79.20%) | 74(61.15%) | 29(57.99%) |
| Insulin | 19074(52.49%) | 811(83.60%) | 172(81.51%) | 465(68.99%) | 72(58.06%) | 252(77.53%) | 47(46.53%) | 93(76.85%) | 39(77.99%) |
| Acarbose | 1429(3.93%) | 53(5.46%) | 8(3.79%) | 29(4.30%) | 3(2.41%) | 12(3.69%) | 8(7.92%) | 6(4.95%) | 2(3.99%) |
| Thiozolidinedone | 9569(26.33%) | 151(15.56%) | 49(23.22%) | 124(18.39%) | 22(17.74%) | 63(19.38%) | 19(18.81%) | 18(14.87%) | 5(9.99%) |
| Glucagon-like peptide-1 receptor agonists | 2797(7.69%) | 19(1.95%) | 4(1.89%) | 17(2.52%) | 2(1.61%) | 7(2.15%) | 3(2.97%) | 3(2.47%) | 1(1.99%) |
| ACEI/ARB | 22352(61.51%) | 651(67.11%) | 116(54.97%) | 423(62.75%) | 72(58.06%) | 214(65.84%) | 52(51.48%) | 79(65.28%) | 31(61.99%) |
| Antidepressants | 2971(8.17%) | 149(15.36%) | 43(20.37%) | 83(12.31%) | 10(8.06%) | 43(13.23%) | 20(19.80%) | 10(8.26%) | 1(1.99%) |
| Antihypertensive drugs | 3051(8.39%) | 120(12.37%) | 17(8.05%) | 41(6.08%) | 6(4.83%) | 23(7.07%) | 2(1.98%) | 8(6.61%) | 5(9.99%) |
| Antihepatitis | 664(1.82%) | 11(1.13%) | 3(1.42%) | 28(4.15%) | 1(0.80%) | 26(7.99%) | 0(0.00%) | 0(0.00%) | 0(0.00%) |
| Anticoagulants | 36332(99.99%) | 968(99.79%) | 210(99.52%) | 674(99.99%) | 124(99.99%) | 325(99.99%) | 101(99.99%) | 121(99.99%) | 50(99.99%) |
| Antiplatelets | 11686(32.16%) | 492(50.72%) | 57(27.01%) | 207(30.71%) | 48(38.70%) | 113(34.76%) | 12(11.88%) | 34(28.09%) | 13(25.99%) |
| Statins and fibrates | 27749(76.37%) | 763(78.65%) | 156(73.93%) | 519(77.00%) | 94(75.80%) | 269(82.76%) | 60(59.40%) | 95(78.51%) | 34(67.99%) |
| Nitrates | 5278(14.52%) | 230(23.71%) | 25(11.84%) | 93(13.79%) | 16(12.90%) | 57(17.53%) | 5(4.95%) | 18(14.87%) | 6(11.99%) |
| Non-steroidal anti-inflammatory drugs | 11369(31.29%) | 478(49.27%) | 54(25.59%) | 199(29.52%) | 48(38.70%) | 108(33.23%) | 9(8.91%) | 34(28.09%) | 13(25.99%) |
| Diuretics | 11095(30.53%) | 530(54.63%) | 71(33.64%) | 227(33.67%) | 43(34.67%) | 111(34.15%) | 23(22.77%) | 49(40.49%) | 20(39.99%) |
| Beta-blockers | 9261(25.48%) | 390(40.20%) | 48(22.74%) | 156(23.14%) | 29(23.38%) | 78(23.99%) | 19(18.81%) | 30(24.79%) | 11(21.99%) |
| Calcium channel blockers | 16462(45.30%) | 582(60.00%) | 114(54.02%) | 374(55.48%) | 75(60.48%) | 188(57.84%) | 44(43.56%) | 65(53.71%) | 23(45.99%) |
| SUbclinical biomarkers |  |  |  |  |  |  |  |  |  |
| Abbreviated MDRD, mL/min/1.73m^2 | 88.6(23.5);n=29687 | 73.5(25.4);n=788 | 81.9(22.0);n=168 | 82.9(23.1);n=537 | 84.9(25.1);n=84 | 81.3(21.6);n=276 | 90.9(22.1);n=83 | 76.1(24.4);n=89 | 74.5(17.7);n=33 |
| Most severe renal damage (<15 mL/min/1.73m^2) | 40.0(0.11%) | 6.0(0.61%) | 0.0(0.00%) | 3.0(0.44%) | 1.0(0.80%) | 0.0(0.00%) | 0.0(0.00%) | 2.0(1.65%) | 0.0(0.00%) |
| Severe renal damage ([15, 30) mL/min/1.73m^2) | 111.0(0.30%) | 23.0(2.37%) | 1.0(0.47%) | 4.0(0.59%) | 1.0(0.80%) | 2.0(0.61%) | 0.0(0.00%) | 1.0(0.82%) | 0.0(0.00%) |
| Moderate to severe renal damage ([30, 45) mL/min/1.73m^2) | 657.0(1.80%) | 72.0(7.42%) | 5.0(2.36%) | 15.0(2.22%) | 3.0(2.41%) | 9.0(2.76%) | 3.0(2.97%) | 2.0(1.65%) | 0.0(0.00%) |
| Mild to moderate renal damage ([45, 60) mL/min/1.73m^2) | 2222.0(6.11%) | 132.0(13.60%) | 19.0(9.00%) | 59.0(8.75%) | 9.0(7.25%) | 28.0(8.61%) | 5.0(4.95%) | 15.0(12.39%) | 8.0(15.99%) |
| Mild renal damage ([60, 90] mL/min/1.73m^2) | 12997.0(35.77%) | 358.0(36.90%) | 90.0(42.65%) | 260.0(38.57%) | 29.0(23.38%) | 151.0(46.46%) | 33.0(32.67%) | 47.0(38.84%) | 17.0(33.99%) |
| Chronic kidney disease (>90 mL/min/1.73m^2) | 13660.0(37.59%) | 197.0(20.30%) | 53.0(25.11%) | 196.0(29.08%) | 41.0(33.06%) | 86.0(26.46%) | 42.0(41.58%) | 22.0(18.18%) | 8.0(15.99%) |
| Neutrophil-to-lymphocyte ratio | 3.1(3.8);n=15970 | 4.5(4.8);n=489 | 3.4(2.8);n=69 | 3.09(2.77);n=221 | 2.7(1.1);n=37 | 3.4(3.1);n=109 | 2.4(1.7);n=36 | 3.3(3.8);n=34 | 2.6(2.4);n=13 |
| Platelet-to-lymphocyte ratio | 133.6(137.8);n=15969 | 156.6(115.1);n=488 | 142.6(87.0);n=69 | 134.0(67.1);n=221 | 126.9(39.2);n=37 | 127.0(69.3);n=109 | 135.5(51.7);n=36 | 152.4(85.3);n=34 | 117.4(59.7);n=13 |
| Neutrophil-to-high-density lipoprotein ratio | 0.3(0.2);n=15080 | 0.3(0.19);n=424 | 0.27(0.14);n=65 | 0.25(0.14);n=210 | 0.27(0.12);n=34 | 0.26(0.14);n=105 | 0.2(0.1);n=33 | 0.26(0.14);n=33 | 0.2(0.1);n=12 |
| Low density lipoprotein ratio-to-high density lipoprotein ratio | 2.1(0.8);n=27933 | 2.2(0.9);n=683 | 2.05(0.7);n=156 | 2.0(0.8);n=511 | 2.06(0.75);n=80 | 2.0(0.8);n=263 | 2.09(0.78);n=78 | 2.0(0.9);n=85 | 2.0(0.8);n=32 |
| Triglyceride-glucose index | 7.6(0.7);n=25514 | 7.62(0.77);n=599 | 7.5(0.6);n=135 | 7.5(0.7);n=431 | 7.5(0.6);n=66 | 7.5(0.7);n=224 | 7.7(0.6);n=70 | 7.5(0.7);n=68 | 7.5(0.8);n=22 |
| Protein-to-creatinine ratio | 3.3(1.5);n=19746 | 2.6(1.4);n=563 | 3.1(1.7);n=93 | 3.1(1.4);n=287 | 3.4(1.6);n=43 | 2.8(1.3);n=144 | 4.1(1.1);n=46 | 2.9(1.3);n=48 | 2.9(1.3);n=16 |
| Aspartate aminotransferase-to-alanine transaminase ratio | 0.9(0.8);n=6377 | 1.0(0.4);n=191 | 0.94(0.31);n=25 | 1.0(0.8);n=108 | 0.88(0.2);n=18 | 1.0(1.0);n=53 | 1.1(0.4);n=10 | 1.1(0.5);n=28 | 1.3(0.2);n=8 |
| Complete blood counts, renal and liver functions |  |  |  |  |  |  |  |  |  |
| Mean corpuscular volume, fL | 86.7(7.2);n=19809 | 87.2(7.6);n=564 | 87.8(6.6);n=93 | 87.5(7.3);n=286 | 88.4(8.7);n=43 | 87.2(7.5);n=143 | 88.6(6.5);n=46 | 86.9(5.6);n=48 | 87.3(6.6);n=16 |
| Eosinophil, x10^9/L | 0.2(0.2);n=15957 | 0.2(0.18);n=483 | 0.2(0.17);n=69 | 0.3(0.6);n=221 | 0.4(1.3);n=37 | 0.19(0.14);n=109 | 0.17(0.1);n=36 | 0.4(0.5);n=34 | 0.5(0.6);n=13 |
| Lymphocyte, x10^9/L | 2.1(0.8);n=15972 | 1.8(1.0);n=489 | 2.0(0.8);n=69 | 2.0(0.8);n=221 | 2.14(0.74);n=37 | 1.9(0.8);n=109 | 2.09(0.5);n=36 | 2.08(0.95);n=34 | 2.3(0.7);n=13 |
| Neutrophil, x10^9/L | 5.2(2.5);n=15972 | 6.0(3.3);n=489 | 5.3(2.2);n=69 | 5.0(2.0);n=221 | 5.21(1.63);n=37 | 5.0(2.1);n=109 | 4.6(1.9);n=36 | 5.0(2.3);n=34 | 4.9(1.4);n=13 |
| White cell count, x10^9/L | 8.0(2.9);n=19826 | 8.4(3.0);n=564 | 8.2(2.3);n=93 | 7.8(2.4);n=287 | 8.5(2.6);n=43 | 7.7(2.3);n=144 | 7.3(2.2);n=46 | 8.4(2.6);n=48 | 8.6(2.1);n=16 |
| Platelet, x10^9/L | 242.1(67.8);n=19824 | 229.8(73.3);n=563 | 240.7(75.1);n=93 | 237.6(75.3);n=287 | 260.5(70.3);n=43 | 217.1(78.6);n=144 | 261.1(60.0);n=46 | 250.6(71.5);n=48 | 242.8(97.0);n=16 |
| Red cell count, x10^12/L | 4.7(0.6);n=19809 | 4.5(0.7);n=564 | 4.6(0.6);n=93 | 4.5(0.6);n=286 | 4.5(0.6);n=43 | 4.5(0.6);n=143 | 4.3(0.6);n=46 | 4.6(0.7);n=48 | 4.8(0.6);n=16 |
| Potassium, mmol/L | 4.3(0.4);n=29604 | 4.32(0.5);n=784 | 4.29(0.46);n=165 | 4.3(0.45);n=532 | 4.4(0.4);n=84 | 4.32(0.47);n=275 | 4.2(0.3);n=79 | 4.28(0.49);n=89 | 4.4(0.6);n=33 |
| Albumin, g/L | 42.5(3.3);n=24595 | 40.1(4.4);n=656 | 41.3(3.3);n=123 | 41.7(3.5);n=386 | 42.0(3.0);n=65 | 41.3(3.7);n=195 | 42.8(2.9);n=56 | 42.0(3.5);n=65 | 42.3(3.5);n=23 |
| Sodium, mmol/L | 139.3(2.7);n=29612 | 138.8(3.9);n=784 | 139.4(3.3);n=165 | 139.3(2.89);n=532 | 138.8(2.6);n=84 | 139.4(2.9);n=275 | 139.6(3.3);n=79 | 139.2(2.8);n=89 | 140.1(2.1);n=33 |
| Urea, mmol/L | 5.8(2.1);n=29607 | 7.1(3.5);n=787 | 6.2(2.9);n=167 | 6.2(2.9);n=536 | 6.0(2.9);n=84 | 6.3(2.2);n=275 | 5.7(3.4);n=83 | 6.9(4.0);n=89 | 7.0(2.1);n=33 |
| Protein, g/L | 74.4(5.0);n=23401 | 73.8(6.5);n=630 | 74.5(5.1);n=123 | 74.9(5.3);n=379 | 74.3(6.5);n=64 | 75.1(4.8);n=192 | 76.1(4.0);n=55 | 74.0(6.0);n=63 | 74.2(7.1);n=23 |
| Creatinine, umol/L | 80.4(33.0);n=29687 | 99.0(50.9);n=788 | 81.9(25.8);n=168 | 85.2(61.4);n=537 | 88.6(50.6);n=84 | 84.4(25.6);n=276 | 65.9(18.0);n=83 | 105.0(132.0);n=89 | 93.0(19.0);n=33 |
| Alkaline phosphatase, U/L | 74.8(27.2);n=24597 | 82.6(32.8);n=655 | 74.9(25.4);n=123 | 79.4(26.8);n=386 | 80.1(31.1);n=65 | 82.4(28.1);n=195 | 79.5(23.8);n=56 | 69.0(17.1);n=65 | 63.3(13.0);n=23 |
| Aspartate transaminase, U/L | 28.8(30.5);n=10477 | 26.1(27.0);n=257 | 24.4(9.1);n=36 | 28.9(18.8);n=173 | 31.4(18.1);n=28 | 31.7(21.2);n=81 | 23.3(11.5);n=24 | 25.0(16.7);n=39 | 22.0(11.4);n=16 |
| Alanine transaminase, U/L | 32.3(27.5);n=20438 | 27.7(39.3);n=584 | 29.0(15.5);n=106 | 29.9(21.0);n=316 | 32.4(21.2);n=50 | 31.7(22.9);n=164 | 26.7(18.0);n=45 | 24.7(15.6);n=54 | 20.7(10.4);n=15 |
| Bilirubin, umol/L | 11.4(5.7);n=24545 | 11.1(6.7);n=655 | 10.9(4.3);n=123 | 11.3(5.7);n=385 | 9.1(4.9);n=65 | 12.6(5.8);n=194 | 10.5(3.7);n=56 | 10.5(6.8);n=65 | 11.9(9.4);n=23 |
| Lipid profiles |  |  |  |  |  |  |  |  |  |
| Triglyceride, mmol/L | 1.8(1.8);n=28423 | 1.7(1.3);n=702 | 1.5(0.7);n=156 | 1.6(1.1);n=512 | 1.6(0.8);n=81 | 1.6(1.0);n=263 | 1.76(0.96);n=78 | 1.7(1.6);n=85 | 1.6(0.9);n=32 |
| SD of triglyceride | 0.5(1.1);n=17175 | 0.4(0.6);n=387 | 0.2(0.2);n=77 | 0.3(0.4);n=266 | 0.4(0.7);n=40 | 0.3(0.2);n=146 | 0.3(0.3);n=37 | 0.4(0.4);n=43 | 0.3(0.3);n=18 |
| Low-density lipoprotein, mmol/L | 2.4(0.8);n=27937 | 2.36(0.83);n=685 | 2.3(0.7);n=157 | 2.2(0.7);n=512 | 2.3(0.7);n=81 | 2.2(0.7);n=263 | 2.43(0.77);n=78 | 2.2(0.7);n=85 | 2.1(0.7);n=32 |
| SD of low-density lipoprotein | 0.3(0.3);n=16636 | 0.4(0.3);n=377 | 0.31(0.29);n=77 | 0.32(0.28);n=266 | 0.29(0.18);n=41 | 0.32(0.28);n=147 | 0.3(0.3);n=35 | 0.33(0.25);n=43 | 0.27(0.15);n=18 |
| High-density lipoprotein, mmol/L | 1.2(0.3);n=28370 | 1.1(0.3);n=699 | 1.16(0.29);n=156 | 1.18(0.31);n=512 | 1.17(0.31);n=81 | 1.18(0.32);n=263 | 1.22(0.25);n=78 | 1.17(0.31);n=85 | 1.1(0.4);n=32 |
| SD of high-density lipoprotein | 0.1(0.1);n=16522 | 0.1(0.1);n=373 | 0.07(0.05);n=76 | 0.09(0.07);n=257 | 0.09(0.05);n=42 | 0.09(0.08);n=138 | 0.1(0.05);n=35 | 0.07(0.04);n=41 | 0.07(0.05);n=17 |
| Total cholesterol, mmol/L | 4.3(1.0);n=28445 | 4.25(1.04);n=702 | 4.1(0.7);n=156 | 4.2(0.9);n=512 | 4.2(0.8);n=81 | 4.1(0.9);n=263 | 4.5(0.8);n=78 | 4.1(1.0);n=85 | 4.0(0.8);n=32 |
| SD of total cholesterol | 0.4(0.4);n=17175 | 0.5(0.4);n=387 | 0.3(0.3);n=77 | 0.38(0.32);n=269 | 0.37(0.24);n=42 | 0.38(0.34);n=146 | 0.3(0.3);n=38 | 0.4(0.3);n=43 | 0.3(0.2);n=18 |
| Hemoglobin A1C, % | 8.3(1.6);n=29345 | 8.4(2.1);n=760 | 8.2(1.4);n=163 | 8.1(1.4);n=530 | 8.2(1.6);n=87 | 8.2(1.4);n=274 | 8.1(1.3);n=79 | 7.8(1.4);n=86 | 7.6(1.2);n=33 |
| SD of hemoglobin A1C | 0.6(0.7);n=22814 | 0.7(1.1);n=542 | 0.57(0.5);n=110 | 0.5(0.5);n=373 | 0.5(0.5);n=63 | 0.5(0.5);n=190 | 0.61(0.44);n=57 | 0.5(0.4);n=61 | 0.5(0.5);n=25 |
| Fasting glucose, mmol/L | 10.1(3.3);n=22141 | 11.4(5.6);n=500 | 10.07(3.77);n=104 | 10.0(3.2);n=342 | 9.7(2.3);n=57 | 10.12(3.71);n=177 | 10.0(2.0);n=55 | 9.7(2.8);n=50 | 9.4(2.5);n=18 |
| SD of fasting glucose | 1.9(1.9);n=18676 | 2.6(2.5);n=482 | 2.0(2.1);n=96 | 1.91(1.85);n=304 | 2.2(1.7);n=45 | 2.0(2.0);n=167 | 1.86(2.12);n=50 | 1.5(1.2);n=39 | 1.7(1.1);n=11 |
| SGLT2I v.s. DPP4I.1 | 18167(50.00%) | 431(44.43%) | 74(35.07%) | 242(35.90%) | 42(33.87%) | 129(39.69%) | 32(31.68%) | 39(32.23%) | 11(21.99%) |
| Dapagliflozin | 10580(29.11%) | 244(25.15%) | 42(19.90%) | 135(20.02%) | 26(20.96%) | 65(19.99%) | 15(14.85%) | 27(22.31%) | 7(13.99%) |
| Empagliflozin | 3768(10.37%) | 76(7.83%) | 12(5.68%) | 53(7.86%) | 8(6.45%) | 29(8.92%) | 7(6.93%) | 9(7.43%) | 3(5.99%) |
| Canagliflozin | 4515(12.42%) | 115(11.85%) | 22(10.42%) | 62(9.19%) | 8(6.45%) | 37(11.38%) | 13(12.87%) | 5(4.13%) | 1(1.99%) |
| Ertugliflozin | 2523(6.94%) | 73(7.52%) | 12(5.68%) | 28(4.15%) | 5(4.03%) | 18(5.53%) | 3(2.97%) | 3(2.47%) | 1(1.99%) |

**Supplementary Table 3C. Baseline and clinical characteristics of patients with/without new onset cancer risk before and after propensity score matching (1:1).**

* for SMD$\geq$0.2; SD: standard deviation; SGLT2I: sodium glucose cotransporter-2 inhibitor; DPP4I: dipeptidyl peptidase-4 inhibitor; SD: standard deviation; SGLT2I: sodium glucose cotransporter-2 inhibitor; DPP4I: dipeptidyl peptidase-4 inhibitor; MDRD: modification of diet in renal disease; # indicated the difference in patients with/without new onset cancer.

|  | **Before matching** |  |  |  | **After matching** |  |  |  |
| --- | --- | --- | --- | --- | --- | --- | --- | --- |
| **Characteristics** | **All (N=60112) Mean(SD);N or Count(%)** | **New onset cancer (N=1533) Mean(SD);N or Count(%)** | **No New onset cancer (N=58579) Mean(SD);N or Count(%)** | **SMD** | **All (N=36334) Mean(SD);N or Count(%)** | **New onset cancer (N=674) Mean(SD);N or Count(%)** | **No New onset cancer (N=35660) Mean(SD);N or Count(%)** | **SMD** |
| Demographics |  |  |  |  |  |  |  |  |
| Male gender | 33883(56.36%) | 851(55.51%) | 33032(56.38%) | 0.02 | 22288(61.34%) | 383(56.82%) | 21905(61.42%) | 0.09 |
| Female gender | 26229(43.63%) | 682(44.48%) | 25547(43.61%) | 0.02 | 14046(38.65%) | 291(43.17%) | 13755(38.57%) | 0.09 |
| Baseline age, years | 62.1(12.4);n=60112 | 67.4(10.5);n=1533 | 61.9(12.4);n=58579 | 0.48* | 58.2(11.0);n=36334 | 63.9(10.2);n=674 | 58.1(11.0);n=35660 | 0.54* |
| 18-50 | 9057(15.06%) | 73(4.76%) | 8984(15.33%) | 0.36* | 7212(19.84%) | 47(6.97%) | 7165(20.09%) | 0.39* |
| 50-60 | 17550(29.19%) | 287(18.72%) | 17263(29.46%) | 0.25* | 13285(36.56%) | 176(26.11%) | 13109(36.76%) | 0.23* |
| 60-70 | 18029(29.99%) | 553(36.07%) | 17476(29.83%) | 0.13 | 11194(30.80%) | 278(41.24%) | 10916(30.61%) | 0.22* |
| 70-80 | 10338(17.19%) | 413(26.94%) | 9925(16.94%) | 0.24* | 3766(10.36%) | 136(20.17%) | 3630(10.17%) | 0.28* |
| >80 | 5145(8.55%) | 207(13.50%) | 4938(8.42%) | 0.16 | 881(2.42%) | 37(5.48%) | 844(2.36%) | 0.16 |
| Past comorbidities |  |  |  |  |  |  |  |  |
| Charlson standard comorbidity index | 1.9(1.4);n=60112 | 2.5(1.3);n=1533 | 1.9(1.4);n=58579 | 0.41* | 1.5(1.2);n=36334 | 2.1(1.2);n=674 | 1.5(1.2);n=35660 | 0.49* |
| Duration from earliest diabetes mellitus date to baseline date, day | 619.9(1341.9);n=60112 | 526.2(1276.3);n=1533 | 622.4(1343.5);n=58579 | 0.07 | 590.9(1323.1);n=36334 | 386.4(1033.8);n=674 | 594.8(1327.6);n=35660 | 0.18 |
| Hypertension | 13376(22.25%) | 384(25.04%) | 12992(22.17%) | 0.07 | 8325(22.91%) | 164(24.33%) | 8161(22.88%) | 0.03 |
| Hyperlipidaemia | 1620(2.69%) | 25(1.63%) | 1595(2.72%) | 0.07 | 1265(3.48%) | 17(2.52%) | 1248(3.49%) | 0.06 |
| Hypotension | 350(0.58%) | 11(0.71%) | 339(0.57%) | 0.02 | 152(0.41%) | 5(0.74%) | 147(0.41%) | 0.04 |
| Overweight, obesity and hyperalimentation | 432(0.71%) | 8(0.52%) | 424(0.72%) | 0.03 | 576(1.58%) | 8(1.18%) | 568(1.59%) | 0.03 |
| Gout | 1510(2.51%) | 46(3.00%) | 1464(2.49%) | 0.03 | 782(2.15%) | 17(2.52%) | 765(2.14%) | 0.02 |
| Heart failure | 1629(2.70%) | 42(2.73%) | 1587(2.70%) | <0.01 | 874(2.40%) | 16(2.37%) | 858(2.40%) | <0.01 |
| Acute myocardial infarction | 1521(2.53%) | 39(2.54%) | 1482(2.52%) | <0.01 | 1210(3.33%) | 18(2.67%) | 1192(3.34%) | 0.04 |
| Ischemic heart disease | 5680(9.44%) | 142(9.26%) | 5538(9.45%) | 0.01 | 4466(12.29%) | 77(11.42%) | 4389(12.30%) | 0.03 |
| Peripheral vascular disease | 375(0.62%) | 8(0.52%) | 367(0.62%) | 0.01 | 194(0.53%) | 2(0.29%) | 192(0.53%) | 0.04 |
| Stroke/transient ischemic attack | 1777(2.95%) | 48(3.13%) | 1729(2.95%) | 0.01 | 940(2.58%) | 19(2.81%) | 921(2.58%) | 0.01 |
| Atrial fibrillation | 1325(2.20%) | 34(2.21%) | 1291(2.20%) | <0.01 | 766(2.10%) | 19(2.81%) | 747(2.09%) | 0.05 |
| Diabetic eye disease | 4045(6.72%) | 97(6.32%) | 3948(6.73%) | 0.02 | 2459(6.76%) | 42(6.23%) | 2417(6.77%) | 0.02 |
| Alcohol dependence | 119(0.19%) | 11(0.71%) | 108(0.18%) | 0.08 | 44(0.12%) | 4(0.59%) | 40(0.11%) | 0.08 |
| Chronic liver disease and cirrhosis | 1195(1.98%) | 31(2.02%) | 1164(1.98%) | <0.01 | 995(2.73%) | 22(3.26%) | 973(2.72%) | 0.03 |
| Viral hepatitis | 629(1.04%) | 30(1.95%) | 599(1.02%) | 0.08 | 414(1.13%) | 11(1.63%) | 403(1.13%) | 0.04 |
| History of acute liver injury | 159(0.26%) | 14(0.91%) | 145(0.24%) | 0.09 | 72(0.19%) | 5(0.74%) | 67(0.18%) | 0.08 |
| Other liver disease | 612(1.01%) | 24(1.56%) | 588(1.00%) | 0.05 | 324(0.89%) | 15(2.22%) | 309(0.86%) | 0.11 |
| Autoimmune disease tissue | 620(1.03%) | 14(0.91%) | 606(1.03%) | 0.01 | 386(1.06%) | 6(0.89%) | 380(1.06%) | 0.02 |
| Other carcinogens pathogen | 13(0.02%) | 1(0.06%) | 12(0.02%) | 0.02 | 12(0.03%) | 5(0.74%) | 7(0.01%) | 0.12 |
| Chronic obstructive pulmonary disease | 69(0.11%) | 1(0.06%) | 68(0.11%) | 0.02 | 24(0.06%) | 4(0.59%) | 20(0.05%) | 0.09 |
| Gastrointestinal disease | 1353(2.25%) | 40(2.60%) | 1313(2.24%) | 0.02 | 690(1.89%) | 22(3.26%) | 668(1.87%) | 0.09 |
| Medications |  |  |  |  |  |  |  |  |
| SGLT2I v.s. DPP4I | 18167(30.22%) | 303(19.76%) | 17864(30.49%) | 0.25* | 18167(50.00%) | 242(35.90%) | 17925(50.26%) | 0.29* |
| SGLT2I frequency | 7.4(10.1);n=18167 | 10.8(14.2);n=303 | 7.3(10.0);n=17864 | 0.29* | 7.3(10.0);n=18167 | 10.6(13.6);n=242 | 7.3(9.9);n=17925 | 0.28* |
| DPP4I frequency | 5.5(7.2);n=41945 | 6.2(8.9);n=1230 | 5.5(7.2);n=40715 | 0.08 | 6.3(8.0);n=18167 | 7.5(10.9);n=432 | 6.2(7.9);n=17735 | 0.13 |
| SGLT2I duration, days | 538.1(674.5);n=18167 | 559.4(657.2);n=303 | 537.7(674.8);n=17864 | 0.03 | 538.5(673.6);n=18167 | 580.3(666.1);n=242 | 538.0(673.7);n=17925 | 0.06 |
| DPP4I duration, days | 528.9(301.0);n=41945 | 470.6(302.8);n=1230 | 530.7(300.8);n=40715 | 0.2 | 575.3(360.3);n=18167 | 498.8(394.6);n=432 | 577.1(359.3);n=17735 | 0.21* |
| Metformin | 54407(90.50%) | 1353(88.25%) | 53054(90.56%) | 0.08 | 33945(93.42%) | 612(90.80%) | 33333(93.47%) | 0.1 |
| Sulphonylurea | 46521(77.39%) | 1204(78.53%) | 45317(77.36%) | 0.03 | 26376(72.59%) | 511(75.81%) | 25865(72.53%) | 0.08 |
| Insulin | 29349(48.82%) | 1051(68.55%) | 28298(48.30%) | 0.42* | 19074(52.49%) | 465(68.99%) | 18609(52.18%) | 0.35* |
| Acarbose | 1566(2.60%) | 46(3.00%) | 1520(2.59%) | 0.02 | 1429(3.93%) | 29(4.30%) | 1400(3.92%) | 0.02 |
| Thiozolidinedone | 12046(20.03%) | 214(13.95%) | 11832(20.19%) | 0.17 | 9569(26.33%) | 124(18.39%) | 9445(26.48%) | 0.19 |
| Glucagon-like peptide-1 receptor agonists | 2044(3.40%) | 19(1.23%) | 2025(3.45%) | 0.15 | 2797(7.69%) | 17(2.52%) | 2780(7.79%) | 0.24* |
| ACEI/ARB | 18249(30.35%) | 449(29.28%) | 17800(30.38%) | 0.02 | 22352(61.51%) | 423(62.75%) | 21929(61.49%) | 0.03 |
| Antidepressants | 2742(4.56%) | 90(5.87%) | 2652(4.52%) | 0.06 | 2971(8.17%) | 83(12.31%) | 2888(8.09%) | 0.14 |
| Antihypertensive drugs | 2311(3.84%) | 47(3.06%) | 2264(3.86%) | 0.04 | 3051(8.39%) | 41(6.08%) | 3010(8.44%) | 0.09 |
| Antihepatitis | 809(1.34%) | 67(4.37%) | 742(1.26%) | 0.19 | 664(1.82%) | 28(4.15%) | 636(1.78%) | 0.14 |
| Anticoagulants | 29401(48.91%) | 677(44.16%) | 28724(49.03%) | 0.1 | 36332(99.99%) | 674(99.99%) | 35658(99.99%) | 0.01 |
| Antiplatelets | 10115(16.82%) | 239(15.59%) | 9876(16.85%) | 0.03 | 11686(32.16%) | 207(30.71%) | 11479(32.19%) | 0.03 |
| Statins and fibrates | 34307(57.07%) | 826(53.88%) | 33481(57.15%) | 0.07 | 27749(76.37%) | 519(77.00%) | 27230(76.36%) | 0.02 |
| Nitrates | 4652(7.73%) | 120(7.82%) | 4532(7.73%) | <0.01 | 5278(14.52%) | 93(13.79%) | 5185(14.54%) | 0.02 |
| Non-steroidal anti-inflammatory drugs | 9719(16.16%) | 226(14.74%) | 9493(16.20%) | 0.04 | 11369(31.29%) | 199(29.52%) | 11170(31.32%) | 0.04 |
| Diuretics | 10179(16.93%) | 314(20.48%) | 9865(16.84%) | 0.09 | 11095(30.53%) | 227(33.67%) | 10868(30.47%) | 0.07 |
| Beta-blockers | 8075(13.43%) | 204(13.30%) | 7871(13.43%) | <0.01 | 9261(25.48%) | 156(23.14%) | 9105(25.53%) | 0.06 |
| Calcium channel blockers | 14154(23.54%) | 386(25.17%) | 13768(23.50%) | 0.04 | 16462(45.30%) | 374(55.48%) | 16088(45.11%) | 0.21* |
| SUbclinical biomarkers |  |  |  |  |  |  |  |  |
| Abbreviated MDRD, mL/min/1.73m^2 | 81.4(27.9);n=49633 | 75.7(25.9);n=1254 | 81.5(28.0);n=48379 | 0.22* | 88.6(23.5);n=29687 | 82.9(23.1);n=537 | 88.7(23.5);n=29150 | 0.25* |
| Most severe renal damage (<15 mL/min/1.73m^2) | 436.0(0.72%) | 14.0(0.91%) | 422.0(0.72%) | 0.02 | 40.0(0.11%) | 3.0(0.44%) | 37.0(0.10%) | 0.07 |
| Severe renal damage ([15, 30) mL/min/1.73m^2) | 1108.0(1.84%) | 32.0(2.08%) | 1076.0(1.83%) | 0.02 | 111.0(0.30%) | 4.0(0.59%) | 107.0(0.30%) | 0.04 |
| Moderate to severe renal damage ([30, 45) mL/min/1.73m^2) | 3548.0(5.90%) | 106.0(6.91%) | 3442.0(5.87%) | 0.04 | 657.0(1.80%) | 15.0(2.22%) | 642.0(1.80%) | 0.03 |
| Mild to moderate renal damage ([45, 60) mL/min/1.73m^2) | 5816.0(9.67%) | 201.0(13.11%) | 5615.0(9.58%) | 0.11 | 2222.0(6.11%) | 59.0(8.75%) | 2163.0(6.06%) | 0.1 |
| Mild renal damage ([60, 90] mL/min/1.73m^2) | 19920.0(33.13%) | 542.0(35.35%) | 19378.0(33.08%) | 0.05 | 12997.0(35.77%) | 260.0(38.57%) | 12737.0(35.71%) | 0.06 |
| Chronic kidney disease (>90 mL/min/1.73m^2) | 18805.0(31.28%) | 359.0(23.41%) | 18446.0(31.48%) | 0.18 | 13660.0(37.59%) | 196.0(29.08%) | 13464.0(37.75%) | 0.18 |
| Neutrophil-to-lymphocyte ratio | 3.4(4.5);n=23978 | 3.9(6.5);n=586 | 3.4(4.5);n=23392 | 0.08 | 3.1(3.8);n=15970 | 3.09(2.77);n=221 | 3.08(3.83);n=15749 | <0.01 |
| Platelet-to-lymphocyte ratio | 142.0(148.2);n=23976 | 145.0(157.6);n=586 | 141.9(148.0);n=23390 | 0.02 | 133.6(137.8);n=15969 | 134.0(67.1);n=221 | 133.6(138.5);n=15748 | <0.01 |
| Neutrophil-to-high-density lipoprotein ratio | 0.3(0.2);n=21968 | 0.28(0.42);n=518 | 0.27(0.18);n=21450 | 0.05 | 0.3(0.2);n=15080 | 0.25(0.14);n=210 | 0.27(0.16);n=14870 | 0.08 |
| Low density lipoprotein ratio-to-high density lipoprotein ratio | 2.1(0.9);n=46116 | 2.0(0.8);n=1147 | 2.1(0.9);n=44969 | 0.08 | 2.1(0.8);n=27933 | 2.0(0.8);n=511 | 2.1(0.8);n=27422 | 0.14 |
| Triglyceride-glucose index | 7.6(0.7);n=42025 | 7.5(0.7);n=1007 | 7.6(0.7);n=41018 | 0.14 | 7.6(0.7);n=25514 | 7.5(0.7);n=431 | 7.6(0.7);n=25083 | 0.16 |
| Protein-to-creatinine ratio | 3.0(1.5);n=29872 | 2.8(1.4);n=732 | 3.0(1.5);n=29140 | 0.17 | 3.3(1.5);n=19746 | 3.1(1.4);n=287 | 3.3(1.5);n=19459 | 0.1 |
| Aspartate aminotransferase-to-alanine transaminase ratio | 1.1(3.2);n=8773 | 1.11(0.74);n=226 | 1.07(3.2);n=8547 | 0.02 | 0.9(0.8);n=6377 | 1.0(0.8);n=108 | 0.9(0.8);n=6269 | 0.14 |
| Complete blood counts, renal and liver functions |  |  |  |  |  |  |  |  |
| Mean corpuscular volume, fL | 87.1(7.5);n=29998 | 87.9(7.7);n=734 | 87.1(7.5);n=29264 | 0.11 | 86.7(7.2);n=19809 | 87.5(7.3);n=286 | 86.7(7.2);n=19523 | 0.1 |
| Eosinophil, x10^9/L | 0.2(0.3);n=23959 | 0.24(0.45);n=586 | 0.22(0.24);n=23373 | 0.06 | 0.2(0.2);n=15957 | 0.3(0.6);n=221 | 0.2(0.2);n=15736 | 0.07 |
| Lymphocyte, x10^9/L | 2.0(0.9);n=23981 | 1.9(0.8);n=586 | 2.0(0.9);n=23395 | 0.12 | 2.1(0.8);n=15972 | 2.0(0.8);n=221 | 2.1(0.8);n=15751 | 0.17 |
| Neutrophil, x10^9/L | 5.3(2.8);n=23981 | 5.2(2.5);n=586 | 5.3(2.8);n=23395 | 0.04 | 5.2(2.5);n=15972 | 5.0(2.0);n=221 | 5.2(2.5);n=15751 | 0.11 |
| White cell count, x10^9/L | 8.0(3.0);n=30008 | 7.9(2.7);n=735 | 8.0(3.0);n=29273 | 0.03 | 8.0(2.9);n=19826 | 7.8(2.4);n=287 | 8.0(2.9);n=19539 | 0.07 |
| Platelet, x10^9/L | 241.3(71.0);n=30006 | 229.3(74.8);n=735 | 241.6(70.9);n=29271 | 0.17 | 242.1(67.8);n=19824 | 237.6(75.3);n=287 | 242.2(67.7);n=19537 | 0.06 |
| Red cell count, x10^12/L | 4.6(0.7);n=29998 | 4.4(0.7);n=734 | 4.6(0.7);n=29264 | 0.21* | 4.7(0.6);n=19809 | 4.5(0.6);n=286 | 4.7(0.6);n=19523 | 0.37* |
| Potassium, mmol/L | 4.3(0.5);n=49471 | 4.4(0.5);n=1250 | 4.3(0.5);n=48221 | 0.02 | 4.3(0.4);n=29604 | 4.3(0.45);n=532 | 4.29(0.44);n=29072 | 0.02 |
| Albumin, g/L | 41.9(3.8);n=37720 | 41.4(3.8);n=924 | 41.9(3.8);n=36796 | 0.14 | 42.5(3.3);n=24595 | 41.7(3.5);n=386 | 42.5(3.3);n=24209 | 0.23* |
| Sodium, mmol/L | 139.3(2.9);n=49494 | 139.4(3.0);n=1250 | 139.3(2.9);n=48244 | 0.04 | 139.3(2.7);n=29612 | 139.3(2.89);n=532 | 139.26(2.74);n=29080 | 0.02 |
| Urea, mmol/L | 6.4(3.3);n=49483 | 6.8(3.6);n=1250 | 6.4(3.3);n=48233 | 0.1 | 5.8(2.1);n=29607 | 6.2(2.9);n=536 | 5.8(2.1);n=29071 | 0.17 |
| Protein, g/L | 73.9(5.4);n=35490 | 74.0(5.6);n=870 | 73.9(5.4);n=34620 | 0.01 | 74.4(5.0);n=23401 | 74.9(5.3);n=379 | 74.4(5.0);n=23022 | 0.09 |
| Creatinine, umol/L | 92.5(71.5);n=49633 | 97.6(77.8);n=1254 | 92.3(71.3);n=48379 | 0.07 | 80.4(33.0);n=29687 | 85.2(61.4);n=537 | 80.3(32.3);n=29150 | 0.1 |
| Alkaline phosphatase, U/L | 76.2(30.1);n=37836 | 78.5(31.4);n=928 | 76.2(30.1);n=36908 | 0.08 | 74.8(27.2);n=24597 | 79.4(26.8);n=386 | 74.7(27.2);n=24211 | 0.17 |
| Aspartate transaminase, U/L | 28.2(53.4);n=15006 | 32.8(61.1);n=378 | 28.1(53.2);n=14628 | 0.08 | 28.8(30.5);n=10477 | 28.9(18.8);n=173 | 28.8(30.7);n=10304 | <0.01 |
| Alanine transaminase, U/L | 29.4(33.6);n=32037 | 28.1(26.8);n=762 | 29.5(33.7);n=31275 | 0.05 | 32.3(27.5);n=20438 | 29.9(21.0);n=316 | 32.3(27.6);n=20122 | 0.1 |
| Bilirubin, umol/L | 11.3(6.7);n=37645 | 11.4(7.9);n=923 | 11.2(6.6);n=36722 | 0.03 | 11.4(5.7);n=24545 | 11.3(5.7);n=385 | 11.4(5.7);n=24160 | 0.02 |
| Lipid profiles |  |  |  |  |  |  |  |  |
| Triglyceride, mmol/L | 1.7(1.6);n=46943 | 1.6(1.0);n=1154 | 1.7(1.6);n=45789 | 0.13 | 1.8(1.8);n=28423 | 1.6(1.1);n=512 | 1.8(1.8);n=27911 | 0.12 |
| SD of triglyceride | 0.5(1.0);n=24256 | 0.3(0.5);n=545 | 0.5(1.0);n=23711 | 0.18 | 0.5(1.1);n=17175 | 0.3(0.4);n=266 | 0.5(1.1);n=16909 | 0.27* |
| Low-density lipoprotein, mmol/L | 2.4(0.8);n=46121 | 2.3(0.8);n=1148 | 2.4(0.8);n=44973 | 0.11 | 2.4(0.8);n=27937 | 2.2(0.7);n=512 | 2.4(0.8);n=27425 | 0.17 |
| SD of low-density lipoprotein | 0.4(0.3);n=23541 | 0.3(0.3);n=542 | 0.4(0.3);n=22999 | 0.06 | 0.3(0.3);n=16636 | 0.32(0.28);n=266 | 0.35(0.32);n=16370 | 0.08 |
| High-density lipoprotein, mmol/L | 1.2(0.3);n=46872 | 1.2(0.33);n=1154 | 1.19(0.33);n=45718 | 0.01 | 1.2(0.3);n=28370 | 1.18(0.31);n=512 | 1.17(0.31);n=27858 | 0.03 |
| SD of high-density lipoprotein | 0.1(0.1);n=23320 | 0.1(0.09);n=522 | 0.1(0.08);n=22798 | 0.06 | 0.1(0.1);n=16522 | 0.09(0.07);n=257 | 0.09(0.08);n=16265 | 0.09 |
| Total cholesterol, mmol/L | 4.3(1.0);n=46986 | 4.2(0.9);n=1154 | 4.3(1.0);n=45832 | 0.14 | 4.3(1.0);n=28445 | 4.2(0.9);n=512 | 4.3(1.0);n=27933 | 0.16 |
| SD of total cholesterol | 0.4(0.4);n=24282 | 0.42(0.38);n=546 | 0.44(0.43);n=23736 | 0.05 | 0.4(0.4);n=17175 | 0.38(0.32);n=269 | 0.43(0.43);n=16906 | 0.12 |
| Hemoglobin A1C, % | 8.1(1.5);n=48920 | 7.9(1.4);n=1213 | 8.1(1.5);n=47707 | 0.13 | 8.3(1.6);n=29345 | 8.1(1.4);n=530 | 8.3(1.6);n=28815 | 0.13 |
| SD of hemoglobin A1C | 0.6(0.7);n=34629 | 0.5(0.5);n=802 | 0.6(0.7);n=33827 | 0.06 | 0.6(0.7);n=22814 | 0.5(0.5);n=373 | 0.6(0.7);n=22441 | 0.05 |
| Fasting glucose, mmol/L | 9.9(3.3);n=34323 | 9.7(3.0);n=795 | 9.9(3.3);n=33528 | 0.06 | 10.1(3.3);n=22141 | 10.0(3.2);n=342 | 10.1(3.3);n=21799 | 0.02 |
| SD of fasting glucose | 1.9(2.1);n=27210 | 2.0(2.0);n=655 | 1.9(2.1);n=26555 | 0.01 | 1.9(1.9);n=18676 | 1.91(1.85);n=304 | 1.88(1.92);n=18372 | 0.02 |

**Supplementary Table 4A. Univariate Cox regression models to predict primary and secondary cancer outcomes before propensity score matching.**

* for p≤ 0.05, ** for p ≤ 0.01, *** for p ≤ 0.001; HR: hazard ratio; CI: confidence interval; SD: standard deviation; SGLT2I: sodium glucose cotransporter-2 inhibitor; DPP4I: dipeptidyl peptidase-4 inhibitor; MDRD: modification of diet in renal disease.

| **Characteristics** | **All.cause.mortality HR [95%** | **Cancer.related.mortality HR [95% CI];P value** | **New.onset.cancer HR [95% CI];P value** | **New.onset.lung.cancer HR [95% CI];P value** | **New.onset.gastrointestinal.cancer HR [95% CI];P value** | **New.onset.breast.cancer HR [95% CI];P value** | **New.onset.genitourinary.cancer HR [95% CI];P value** | **New.onset.bladder.cancer HR [95% CI];P value** |
| --- | --- | --- | --- | --- | --- | --- | --- | --- |
| Demographics |  |  |  |  |  |  |  |  |
| Male gender | 1.17[1.09-1.26];<0.0001*** | 1.20[1.00-1.43];0.0474* | 0.97[0.87-1.07];0.5116 | 1.67[1.28-2.18];0.0002*** | 1.44[1.24-1.66];<0.0001*** | 0.02[0.01-0.05];<0.0001*** | 0.88[0.69-1.13];0.3175 | 4.60[2.61-8.11];<0.0001*** |
| Female gender | 1.0[Reference] | 1.0[Reference] | 1.0[Reference] | 1.0[Reference] | 1.0[Reference] | 1.0[Reference] | 1.0[Reference] | 1.0[Reference] |
| Baseline age, years | 1.08[1.08-1.09];<0.0001*** | 1.06[1.06-1.07];<0.0001*** | 1.04[1.03-1.04];<0.0001*** | 1.04[1.03-1.05];<0.0001*** | 1.04[1.04-1.05];<0.0001*** | 1.02[1.01-1.03];0.0011** | 1.03[1.02-1.05];<0.0001*** | 1.07[1.05-1.09];<0.0001*** |
| 18-50 | 0.20[0.17-0.25];<0.0001*** | 0.17[0.10-0.28];<0.0001*** | 0.28[0.22-0.35];<0.0001*** | 0.09[0.03-0.24];<0.0001*** | 0.20[0.14-0.29];<0.0001*** | 0.51[0.31-0.84];0.0082** | 0.48[0.31-0.76];0.0015** | 0.30[0.12-0.74];0.0089** |
| 50-60 | 1.0[Reference] | 1.0[Reference] | 1.0[Reference] | 1.0[Reference] | 1.0[Reference] | 1.0[Reference] | 1.0[Reference] | 1.0[Reference] |
| 60-70 | 0.66[0.61-0.72];<0.0001*** | 0.86[0.71-1.05];0.1428 | 1.31[1.18-1.45];<0.0001*** | 1.38[1.07-1.78];0.0143* | 1.49[1.30-1.72];<0.0001*** | 1.08[0.80-1.45];0.6104 | 0.88[0.67-1.16];0.3584 | 0.80[0.51-1.26];0.3416 |
| 70-80 | 2.10[1.94-2.27];<0.0001*** | 2.63[2.19-3.16];<0.0001*** | 1.82[1.62-2.03];<0.0001*** | 2.16[1.65-2.82];<0.0001*** | 1.88[1.61-2.19];<0.0001*** | 1.54[1.11-2.13];0.0092** | 1.53[1.15-2.03];0.0036** | 2.31[1.50-3.53];0.0001*** |
| >80 | 5.72[5.30-6.17];<0.0001*** | 2.93[2.35-3.64];<0.0001*** | 1.79[1.54-2.07];<0.0001*** | 1.45[0.98-2.15];0.0643 | 1.83[1.50-2.24];<0.0001*** | 1.26[0.79-2.00];0.3254 | 2.51[1.83-3.44];<0.0001*** | 3.77[2.38-5.98];<0.0001*** |
| Past comorbidities |  |  |  |  |  |  |  |  |
| Charlson standard comorbidity index | 1.62[1.60-1.65];<0.0001*** | 1.45[1.39-1.52];<0.0001*** | 1.30[1.26-1.34];<0.0001*** | 1.34[1.25-1.44];<0.0001*** | 1.33[1.27-1.38];<0.0001*** | 1.16[1.05-1.27];0.0022** | 1.26[1.16-1.36];<0.0001*** | 1.44[1.29-1.60];<0.0001*** |
| Duration from earliest diabetes mellitus date to baseline date, day | 1.000[1.000-1.000];<0.0001*** | 1.000[1.000-1.000];<0.0001*** | 1.000[1.000-1.000];0.0051** | 1.000[1.000-1.000];0.3557 | 1.000[1.000-1.000];0.0119* | 1.000[1.000-1.000];0.9619 | 1.000[1.000-1.000];0.1904 | 1.000[1.000-1.000];0.1311 |
| Hypertension | 1.64[1.52-1.77];<0.0001*** | 1.10[0.89-1.35];0.3715 | 1.18[1.05-1.32];0.0054** | 1.09[0.82-1.47];0.5446 | 1.17[0.99-1.37];0.0589 | 1.36[1.00-1.85];0.0498* | 1.22[0.92-1.61];0.1656 | 1.16[0.73-1.84];0.5294 |
| Hyperlipidaemia | 0.72[0.56-0.93];0.0127* | 0.72[0.39-1.35];0.3090 | 0.59[0.40-0.88];0.0097** | 0.29[0.07-1.17];0.0821 | 0.44[0.24-0.83];0.0107* | 0.92[0.38-2.23];0.8479 | 0.85[0.38-1.90];0.6857 | 0.76[0.19-3.07];0.6966 |
| Hypotension | 2.82[2.11-3.76];<0.0001*** | 0.71[0.18-2.86];0.6318 | 1.28[0.71-2.32];0.4080 | 2.17[0.70-6.78];0.1820 | 1.09[0.45-2.63];0.8459 | 0.89[0.12-6.34];0.9060 | 0.68[0.10-4.86];0.7030 | 1.86[0.26-13.32];0.5381 |
| Overweight, obesity and hyperalimentation | 0.22[0.09-0.53];0.0008*** | 0.27[0.04-1.90];0.1879 | 0.72[0.36-1.43];0.3442 | 0.00[0.00-Inf];0.9863 | 0.33[0.08-1.34];0.1212 | 2.07[0.66-6.48];0.2101 | 1.05[0.26-4.24];0.9414 | 1.42[0.20-10.20];0.7262 |
| Gout | 1.96[1.65-2.31];<0.0001*** | 1.13[0.66-1.91];0.6635 | 1.22[0.91-1.63];0.1878 | 1.14[0.54-2.41];0.7357 | 1.24[0.83-1.85];0.2866 | 0.80[0.30-2.15];0.6565 | 1.73[0.95-3.17];0.0746 | 2.14[0.87-5.27];0.0972 |
| Heart failure | 3.46[3.04-3.94];<0.0001*** | 1.08[0.64-1.84];0.7664 | 1.06[0.78-1.44];0.7144 | 1.09[0.51-2.30];0.8278 | 0.99[0.64-1.53];0.9674 | 0.96[0.39-2.33];0.9239 | 1.50[0.79-2.81];0.2120 | 1.20[0.38-3.79];0.7543 |
| Acute myocardial infarction | 1.80[1.52-2.15];<0.0001*** | 1.11[0.65-1.89];0.6943 | 1.02[0.74-1.40];0.9069 | 0.64[0.24-1.71];0.3699 | 1.13[0.75-1.71];0.5574 | 0.59[0.19-1.85];0.3646 | 1.72[0.94-3.14];0.0795 | 2.57[1.13-5.88];0.0248* |
| Ischemic heart disease | 1.30[1.17-1.45];<0.0001*** | 0.92[0.67-1.25];0.5764 | 0.98[0.83-1.17];0.8323 | 0.98[0.64-1.50];0.9244 | 1.04[0.83-1.32];0.7116 | 0.72[0.42-1.24];0.2352 | 1.20[0.82-1.77];0.3481 | 1.76[1.02-3.05];0.0437* |
| Peripheral vascular disease | 3.87[3.04-4.92];<0.0001*** | 1.68[0.70-4.05];0.2485 | 0.86[0.43-1.73];0.6801 | 2.01[0.65-6.29];0.2282 | 0.20[0.03-1.43];0.1096 | 0.83[0.12-5.90];0.8497 | 1.28[0.32-5.13];0.7320 | 3.49[0.86-14.16];0.0803 |
| Stroke/transient ischemic attack | 2.18[1.88-2.53];<0.0001*** | 1.25[0.78-1.99];0.3591 | 1.08[0.81-1.44];0.5829 | 1.70[0.95-3.04];0.0725 | 0.97[0.64-1.47];0.8890 | 1.03[0.46-2.33];0.9381 | 0.92[0.44-1.96];0.8369 | 1.83[0.74-4.49];0.1895 |
| Atrial fibrillation | 2.69[2.30-3.15];<0.0001*** | 1.12[0.63-1.99];0.6908 | 1.04[0.74-1.46];0.8337 | 0.75[0.28-2.01];0.5637 | 0.97[0.60-1.57];0.9040 | 1.41[0.63-3.18];0.4073 | 1.45[0.72-2.92];0.3038 | 1.97[0.73-5.37];0.1829 |
| Diabetic eye disease | 1.71[1.53-1.92];<0.0001*** | 0.95[0.66-1.35];0.7625 | 0.94[0.77-1.16];0.5810 | 0.77[0.44-1.35];0.3589 | 0.83[0.62-1.12];0.2264 | 0.97[0.55-1.70];0.9067 | 1.10[0.69-1.75];0.6963 | 1.09[0.50-2.35];0.8301 |
| Alcohol dependence | 5.56[3.86-8.01];<0.0001*** | 4.58[1.71-12.24];0.0024** | 4.09[2.26-7.40];<0.0001*** | 0.00[0.00-Inf];0.9896 | 7.75[4.27-14.05];<0.0001*** | 0.00[0.00-Inf];0.9906 | 0.00[0.00-Inf];0.9893 | 0.00[0.00-Inf];0.9935 |
| Chronic liver disease and cirrhosis | 0.94[0.72-1.22];0.6335 | 0.59[0.26-1.32];0.1967 | 1.01[0.71-1.45];0.9383 | 0.60[0.19-1.87];0.3778 | 1.36[0.89-2.08];0.1532 | 0.49[0.12-1.99];0.3213 | 0.96[0.40-2.33];0.9273 | 1.04[0.26-4.20];0.9616 |
| Viral hepatitis | 1.00[0.71-1.42];0.9921 | 1.90[1.02-3.55];0.0448* | 1.90[1.32-2.73];0.0005*** | 0.76[0.19-3.08];0.7057 | 3.14[2.12-4.64];<0.0001*** | 0.00[0.00-Inf];0.9904 | 0.73[0.18-2.93];0.6548 | 0.98[0.14-7.05];0.9867 |
| History of acute liver injury | 3.17[2.12-4.74];<0.0001*** | 1.58[0.39-6.34];0.5173 | 3.75[2.22-6.35];<0.0001*** | 0.00[0.00-Inf];0.9876 | 6.57[3.80-11.36];<0.0001*** | 1.98[0.28-14.14];0.4952 | 0.00[0.00-Inf];0.9873 | 0.00[0.00-Inf];0.9923 |
| Other liver disease | 1.68[1.27-2.21];0.0002*** | 0.39[0.10-1.56];0.1837 | 1.57[1.05-2.35];0.0288* | 0.79[0.20-3.20];0.7465 | 2.23[1.39-3.55];0.0008*** | 1.49[0.47-4.64];0.4964 | 0.00[0.00-Inf];0.9893 | 0.00[0.00-Inf];0.9935 |
| Autoimmune disease tissue | 0.69[0.45-1.05];0.0853 | 0.38[0.09-1.51];0.1676 | 0.88[0.52-1.48];0.6253 | 0.77[0.19-3.10];0.7138 | 1.18[0.63-2.20];0.6007 | 0.96[0.24-3.86];0.9517 | 0.00[0.00-Inf];0.9891 | 0.00[0.00-Inf];0.9934 |
| Other carcinogens pathogen | 1.55[0.22-11.03];0.6596 | 0.00[0.00-Inf];0.9883 | 3.31[0.47-23.53];0.2312 | 20.72[2.91-147.71];0.0025** | 0.00[0.00-Inf];0.9850 | 0.00[0.00-Inf];0.9926 | 0.00[0.00-Inf];0.9916 | 0.00[0.00-Inf];0.9949 |
| Chronic obstructive pulmonary disease | 2.38[1.19-4.75];0.0145* | 0.00[0.00-Inf];0.9883 | 0.58[0.08-4.09];0.5812 | 3.59[0.50-25.58];0.2022 | 0.00[0.00-Inf];0.9850 | 0.00[0.00-Inf];0.9926 | 0.00[0.00-Inf];0.9916 | 0.00[0.00-Inf];0.9949 |
| Gastrointestinal disease | 1.42[1.15-1.74];0.0008*** | 0.70[0.35-1.42];0.3262 | 1.18[0.86-1.61];0.3055 | 0.90[0.37-2.18];0.8134 | 0.99[0.62-1.58];0.9609 | 1.58[0.74-3.36];0.2334 | 1.75[0.93-3.29];0.0833 | 2.89[1.27-6.61];0.0117* |
| Medications |  |  |  |  |  |  |  |  |
| SGLT2I v.s. DPP4I | 0.44[0.40-0.48];<0.0001*** | 0.66[0.54-0.82];0.0001*** | 0.56[0.49-0.63];<0.0001*** | 0.54[0.40-0.74];0.0001*** | 0.51[0.43-0.61];<0.0001*** | 0.64[0.46-0.89];0.0082** | 0.64[0.47-0.85];0.0025** | 0.75[0.47-1.18];0.2152 |
| SGLT2I frequency | 1.02[1.01-1.02];<0.0001*** | 1.02[1.01-1.03];<0.0001*** | 1.02[1.01-1.02];<0.0001*** | 1.01[1.00-1.03];0.0556 | 1.02[1.01-1.02];<0.0001*** | 1.01[1.00-1.03];0.0576 | 1.02[1.01-1.03];0.0002*** | 1.02[1.00-1.03];0.0309* |
| DPP4I frequency | 0.99[0.98-1.00];0.0018** | 1.01[0.99-1.02];0.3473 | 1.01[1.00-1.01];0.0004*** | 1.01[1.00-1.02];0.0265* | 1.01[1.00-1.01];0.0004*** | 1.01[1.00-1.02];0.1634 | 0.98[0.96-1.01];0.1728 | 0.96[0.92-1.01];0.1208 |
| SGLT2I duration, days | 1.000[1.000-1.000];0.0015** | 1.000[1.000-1.000];0.6099 | 1.000[1.000-1.000];0.6043 | 1.000[1.000-1.000];0.7280 | 1.000[1.000-1.000];0.9896 | 1.000[1.000-1.001];0.6297 | 1.000[1.000-1.000];0.9152 | 1.000[0.999-1.001];0.8569 |
| DPP4I duration, days | 0.999[0.999-0.999];<0.0001*** | 0.999[0.998-0.999];<0.0001*** | 0.999[0.999-0.999];<0.0001*** | 0.999[0.999-1.000];0.0035** | 0.999[0.999-0.999];<0.0001*** | 1.000[1.000-1.001];0.8994 | 0.999[0.999-1.000];0.0324* | 1.000[0.999-1.001];0.5443 |
| Metformin | 0.27[0.25-0.30];<0.0001*** | 0.58[0.45-0.74];<0.0001*** | 0.75[0.64-0.88];0.0003*** | 0.83[0.55-1.23];0.3476 | 0.75[0.61-0.93];0.0094** | 0.86[0.55-1.35];0.5188 | 0.65[0.45-0.93];0.0173* | 0.55[0.32-0.95];0.0322* |
| Sulphonylurea | 1.04[0.96-1.14];0.3163 | 0.94[0.77-1.16];0.5637 | 1.07[0.95-1.21];0.2754 | 1.36[0.98-1.89];0.0625 | 1.01[0.86-1.19];0.8839 | 1.01[0.73-1.41];0.9381 | 1.05[0.78-1.40];0.7605 | 1.00[0.62-1.60];0.9888 |
| Insulin | 4.96[4.52-5.44];<0.0001*** | 3.17[2.59-3.87];<0.0001*** | 2.36[2.12-2.62];<0.0001*** | 1.90[1.47-2.46];<0.0001*** | 3.26[2.78-3.82];<0.0001*** | 1.13[0.86-1.49];0.3929 | 2.11[1.63-2.73];<0.0001*** | 2.18[1.43-3.33];0.0003*** |
| Acarbose | 1.17[0.95-1.44];0.1350 | 1.14[0.68-1.91];0.6056 | 1.16[0.87-1.56];0.3208 | 0.93[0.41-2.08];0.8521 | 1.08[0.72-1.64];0.7016 | 1.35[0.64-2.88];0.4322 | 1.49[0.79-2.81];0.2124 | 1.61[0.59-4.39];0.3496 |
| Thiozolidinedone | 0.41[0.36-0.46];<0.0001*** | 0.62[0.48-0.80];0.0002*** | 0.64[0.55-0.73];<0.0001*** | 0.71[0.50-1.00];0.0498* | 0.59[0.48-0.72];<0.0001*** | 0.69[0.47-1.02];0.0606 | 0.61[0.43-0.87];0.0063** | 0.66[0.38-1.17];0.1543 |
| Glucagon-like peptide-1 receptor agonists | 0.18[0.12-0.28];<0.0001*** | 0.22[0.08-0.59];0.0026** | 0.35[0.22-0.55];<0.0001*** | 0.23[0.06-0.91];0.0362* | 0.24[0.11-0.51];0.0002*** | 0.42[0.14-1.32];0.1394 | 0.55[0.23-1.32];0.1801 | 0.89[0.28-2.81];0.8449 |
| ACEI/ARB | 0.78[0.71-0.84];<0.0001*** | 0.77[0.63-0.94];0.0101* | 0.94[0.85-1.05];0.3057 | 0.98[0.75-1.29];0.8977 | 0.88[0.75-1.02];0.0924 | 0.95[0.70-1.28];0.7240 | 1.12[0.87-1.45];0.3855 | 1.23[0.81-1.87];0.3302 |
| Antidepressants | 1.74[1.52-1.99];<0.0001*** | 2.17[1.61-2.93];<0.0001*** | 1.32[1.06-1.63];0.0114* | 1.65[1.02-2.66];0.0418* | 1.23[0.91-1.66];0.1780 | 1.46[0.83-2.56];0.1881 | 1.29[0.76-2.17];0.3438 | 0.44[0.11-1.80];0.2561 |
| Antihypertensive drugs | 0.93[0.77-1.13];0.4760 | 0.92[0.57-1.47];0.7132 | 0.79[0.59-1.05];0.1045 | 0.61[0.27-1.38];0.2383 | 0.82[0.55-1.21];0.3127 | 0.50[0.19-1.36];0.1761 | 0.89[0.46-1.73];0.7289 | 1.64[0.72-3.75];0.2395 |
| Antihepatitis | 1.00[0.73-1.35];0.9748 | 1.92[1.11-3.33];0.0203* | 3.45[2.70-4.41];<0.0001*** | 1.19[0.44-3.20];0.7261 | 5.77[4.41-7.53];<0.0001*** | 0.73[0.18-2.96];0.6642 | 0.85[0.27-2.65];0.7769 | 1.54[0.38-6.24];0.5471 |
| Anticoagulants | 0.76[0.71-0.82];<0.0001*** | 0.93[0.78-1.10];0.3869 | 0.82[0.74-0.91];0.0001*** | 0.95[0.74-1.22];0.6842 | 0.79[0.69-0.91];0.0010** | 0.74[0.56-0.99];0.0391* | 0.88[0.69-1.13];0.3179 | 0.86[0.58-1.28];0.4637 |
| Antiplatelets | 1.29[1.18-1.41];<0.0001*** | 0.89[0.70-1.13];0.3487 | 0.92[0.80-1.05];0.2121 | 1.22[0.89-1.66];0.2195 | 0.90[0.75-1.09];0.2983 | 0.61[0.39-0.95];0.0283* | 0.95[0.68-1.32];0.7664 | 1.13[0.68-1.89];0.6388 |
| Statins and fibrates | 0.54[0.50-0.58];<0.0001*** | 0.79[0.67-0.95];0.0096** | 0.87[0.79-0.96];0.0059** | 0.95[0.74-1.23];0.7181 | 0.83[0.73-0.96];0.0097** | 0.92[0.69-1.21];0.5420 | 0.89[0.70-1.13];0.3390 | 0.73[0.49-1.08];0.1181 |
| Nitrates | 1.43[1.27-1.60];<0.0001*** | 0.98[0.70-1.36];0.8863 | 1.02[0.85-1.23];0.8403 | 1.11[0.71-1.73];0.6592 | 1.09[0.85-1.40];0.4964 | 0.63[0.33-1.19];0.1513 | 1.22[0.80-1.85];0.3623 | 1.23[0.62-2.44];0.5566 |
| Non-steroidal anti-inflammatory drugs | 1.27[1.16-1.39];<0.0001*** | 0.88[0.68-1.12];0.2999 | 0.90[0.78-1.04];0.1393 | 1.24[0.91-1.70];0.1755 | 0.89[0.73-1.08];0.2244 | 0.51[0.32-0.83];0.0066** | 0.97[0.69-1.35];0.8550 | 1.11[0.66-1.87];0.7069 |
| Diuretics | 1.73[1.60-1.88];<0.0001*** | 1.45[1.17-1.78];0.0005*** | 1.28[1.13-1.45];0.0001*** | 1.15[0.84-1.59];0.3756 | 1.31[1.11-1.55];0.0015** | 0.97[0.67-1.41];0.8890 | 1.65[1.24-2.18];0.0005*** | 1.46[0.91-2.34];0.1214 |
| Beta-blockers | 1.40[1.27-1.53];<0.0001*** | 1.04[0.81-1.34];0.7575 | 0.99[0.86-1.15];0.9409 | 1.09[0.77-1.56];0.6148 | 0.93[0.76-1.15];0.5163 | 0.92[0.61-1.40];0.6967 | 1.24[0.89-1.73];0.1977 | 1.00[0.56-1.80];0.9928 |
| Calcium channel blockers | 1.14[1.05-1.23];0.0022** | 1.04[0.85-1.28];0.6971 | 1.09[0.97-1.23];0.1273 | 1.08[0.81-1.43];0.6177 | 1.03[0.88-1.21];0.7065 | 1.10[0.80-1.52];0.5440 | 1.36[1.04-1.77];0.0238* | 1.25[0.80-1.95];0.3217 |
| SUbclinical biomarkers |  |  |  |  |  |  |  |  |
| Abbreviated MDRD, mL/min/1.73m^2 | 0.971[0.969-0.972];<0.0001*** | 0.99[0.98-0.99];<0.0001*** | 0.992[0.990-0.994];<0.0001*** | 1.00[0.99-1.00];0.0767 | 0.991[0.988-0.994];<0.0001*** | 1.00[0.99-1.00];0.1497 | 0.99[0.98-0.99];<0.0001*** | 0.98[0.98-0.99];0.0002*** |
| Most severe renal damage (<15 mL/min/1.73m^2) | 7.06[5.92-8.43];<0.0001*** | 0.00[0.00-Inf];0.9885 | 1.39[0.82-2.36];0.2187 | 0.64[0.09-4.53];0.6512 | 1.11[0.50-2.47];0.8041 | 2.21[0.71-6.92];0.1736 | 2.37[0.88-6.37];0.0874 | 0.00[0.00-Inf];0.9946 |
| Severe renal damage ([15, 30) mL/min/1.73m^2) | 5.16[4.51-5.90];<0.0001*** | 1.68[0.97-2.93];0.0648 | 1.24[0.87-1.76];0.2299 | 1.25[0.51-3.04];0.6231 | 1.08[0.65-1.80];0.7655 | 0.56[0.14-2.26];0.4144 | 2.10[1.08-4.10];0.0292* | 1.79[0.57-5.68];0.3212 |
| Moderate to severe renal damage ([30, 45) mL/min/1.73m^2) | 3.09[2.79-3.42];<0.0001*** | 1.78[1.30-2.43];0.0003*** | 1.24[1.01-1.51];0.0353* | 1.04[0.60-1.79];0.8935 | 1.32[1.01-1.72];0.0424* | 1.29[0.76-2.19];0.3456 | 1.17[0.71-1.93];0.5283 | 1.06[0.46-2.43];0.8960 |
| Mild to moderate renal damage ([45, 60) mL/min/1.73m^2) | 1.76[1.59-1.95];<0.0001*** | 1.48[1.13-1.94];0.0045** | 1.46[1.26-1.70];<0.0001*** | 1.12[0.74-1.71];0.5950 | 1.51[1.23-1.85];0.0001*** | 1.24[0.81-1.92];0.3206 | 1.67[1.18-2.38];0.0040** | 2.80[1.72-4.56];<0.0001*** |
| Mild renal damage ([60, 90] mL/min/1.73m^2) | 0.72[0.66-0.78];<0.0001*** | 1.22[1.00-1.49];0.0491* | 1.13[1.01-1.26];0.0323* | 1.12[0.84-1.49];0.4280 | 1.18[1.01-1.37];0.0347* | 0.98[0.72-1.33];0.8892 | 1.26[0.96-1.65];0.0916 | 1.10[0.71-1.71];0.6578 |
| Chronic kidney disease (>90 mL/min/1.73m^2) | 0.29[0.26-0.32];<0.0001*** | 0.50[0.39-0.63];<0.0001*** | 0.64[0.57-0.73];<0.0001*** | 0.82[0.61-1.11];0.2010 | 0.60[0.51-0.71];<0.0001*** | 0.85[0.62-1.16];0.3069 | 0.45[0.32-0.62];<0.0001*** | 0.39[0.23-0.67];0.0007*** |
| Neutrophil-to-lymphocyte ratio | 1.04[1.03-1.05];<0.0001*** | 1.03[1.01-1.05];0.0038** | 1.02[1.01-1.03];0.0054** | 0.99[0.94-1.05];0.8196 | 1.03[1.01-1.04];0.0003*** | 0.99[0.93-1.05];0.6652 | 1.02[0.99-1.05];0.1839 | 1.02[0.96-1.07];0.5595 |
| Platelet-to-lymphocyte ratio | 1.000[1.000-1.001];<0.0001*** | 1.000[1.000-1.001];0.4833 | 1.000[1.000-1.001];0.5212 | 0.999[0.997-1.002];0.6541 | 1.000[1.000-1.001];0.7252 | 1.000[0.999-1.001];0.9753 | 1.000[1.000-1.001];0.2130 | 1.000[0.999-1.001];0.4263 |
| Neutrophil-to-high-density lipoprotein ratio | 1.55[1.39-1.72];<0.0001*** | 1.31[0.79-2.17];0.2943 | 1.35[1.01-1.80];0.0421* | 0.88[0.23-3.26];0.8431 | 1.59[1.24-2.04];0.0003*** | 0.41[0.08-2.20];0.2984 | 1.04[0.37-2.97];0.9384 | 0.84[0.11-6.30];0.8695 |
| Low density lipoprotein ratio-to-high density lipoprotein ratio | 1.04[0.99-1.09];0.0985 | 0.90[0.79-1.02];0.0991 | 0.91[0.85-0.98];0.0127* | 0.79[0.65-0.95];0.0142* | 0.93[0.85-1.03];0.1633 | 0.94[0.77-1.14];0.5208 | 0.94[0.79-1.11];0.4467 | 0.88[0.66-1.17];0.3716 |
| Triglyceride-glucose index | 0.90[0.84-0.96];0.0009*** | 0.72[0.61-0.84];0.0001*** | 0.82[0.75-0.90];<0.0001*** | 0.79[0.63-0.99];0.0368* | 0.78[0.69-0.88];0.0001*** | 1.14[0.91-1.44];0.2631 | 0.80[0.64-0.99];0.0394* | 0.66[0.46-0.94];0.0230* |
| Protein-to-creatinine ratio | 0.59[0.56-0.61];<0.0001*** | 0.82[0.74-0.90];0.0001*** | 0.88[0.83-0.92];<0.0001*** | 0.98[0.87-1.11];0.7794 | 0.74[0.69-0.81];<0.0001*** | 1.18[1.07-1.31];0.0013** | 0.88[0.77-1.01];0.0656 | 0.77[0.61-0.97];0.0258* |
| Aspartate aminotransferase-to-alanine transaminase ratio | 1.01[1.00-1.02];0.1556 | 1.01[0.97-1.04];0.7076 | 1.00[0.98-1.03];0.8114 | 1.00[0.91-1.10];0.9910 | 1.01[0.98-1.04];0.7228 | 0.99[0.73-1.33];0.9320 | 1.00[0.92-1.09];0.9898 | 1.00[0.88-1.14];0.9804 |
| Complete blood counts, renal and liver functions |  |  |  |  |  |  |  |  |
| Mean corpuscular volume, fL | 1.02[1.02-1.03];<0.0001*** | 1.03[1.01-1.05];0.0024** | 1.02[1.01-1.03];0.0015** | 1.04[1.01-1.07];0.0181* | 1.03[1.01-1.04];0.0005*** | 1.00[0.97-1.03];0.9351 | 1.00[0.97-1.02];0.8457 | 0.99[0.95-1.03];0.5941 |
| Eosinophil, x10^9/L | 1.07[0.91-1.26];0.4123 | 0.75[0.35-1.59];0.4514 | 1.19[1.01-1.40];0.0332* | 1.33[1.09-1.62];0.0043** | 0.97[0.61-1.55];0.8916 | 0.67[0.22-2.06];0.4814 | 1.30[1.05-1.61];0.0158* | 1.31[0.95-1.82];0.1004 |
| Lymphocyte, x10^9/L | 0.55[0.52-0.60];<0.0001*** | 0.74[0.61-0.90];0.0028** | 0.83[0.75-0.93];0.0009*** | 0.95[0.73-1.25];0.7251 | 0.75[0.65-0.88];0.0002*** | 1.00[0.79-1.27];0.9908 | 0.86[0.66-1.12];0.2704 | 0.66[0.42-1.04];0.0752 |
| Neutrophil, x10^9/L | 1.06[1.05-1.07];<0.0001*** | 1.03[0.98-1.07];0.2112 | 0.99[0.96-1.02];0.4787 | 0.99[0.91-1.08];0.8496 | 0.99[0.95-1.04];0.7622 | 1.00[0.92-1.08];0.9250 | 1.00[0.92-1.07];0.9025 | 0.96[0.83-1.10];0.5426 |
| White cell count, x10^9/L | 1.02[1.01-1.03];<0.0001*** | 1.01[0.97-1.05];0.7029 | 0.99[0.96-1.02];0.4086 | 1.01[0.97-1.06];0.5657 | 0.97[0.93-1.01];0.1420 | 0.99[0.92-1.06];0.8030 | 1.02[0.99-1.05];0.2411 | 1.01[0.95-1.09];0.7211 |
| Platelet, x10^9/L | 0.997[0.996-0.998];<0.0001*** | 0.997[0.995-0.999];0.0069** | 0.997[0.996-0.998];<0.0001*** | 1.001[0.998-1.003];0.6943 | 0.99[0.99-1.00];<0.0001*** | 1.002[0.999-1.004];0.1941 | 1.001[0.998-1.003];0.6288 | 1.00[0.99-1.00];0.5063 |
| Red cell count, x10^12/L | 0.40[0.38-0.43];<0.0001*** | 0.62[0.51-0.76];<0.0001*** | 0.71[0.64-0.79];<0.0001*** | 0.81[0.61-1.07];0.1340 | 0.71[0.62-0.83];<0.0001*** | 0.59[0.44-0.79];0.0004*** | 0.71[0.54-0.93];0.0127* | 0.93[0.60-1.44];0.7349 |
| Potassium, mmol/L | 1.19[1.09-1.29];<0.0001*** | 0.98[0.80-1.21];0.8709 | 1.05[0.94-1.18];0.3905 | 1.58[1.19-2.10];0.0015** | 1.00[0.85-1.18];0.9819 | 0.83[0.60-1.14];0.2516 | 1.05[0.79-1.39];0.7454 | 1.54[0.99-2.38];0.0539 |
| Albumin, g/L | 0.86[0.85-0.86];<0.0001*** | 0.93[0.90-0.95];<0.0001*** | 0.96[0.95-0.98];<0.0001*** | 1.02[0.97-1.07];0.4058 | 0.95[0.93-0.97];<0.0001*** | 1.00[0.96-1.05];0.9592 | 0.95[0.91-0.98];0.0045** | 0.93[0.88-0.98];0.0133* |
| Sodium, mmol/L | 0.95[0.94-0.96];<0.0001*** | 1.01[0.98-1.05];0.4674 | 1.01[0.99-1.03];0.1525 | 1.03[0.98-1.08];0.2990 | 1.00[0.98-1.03];0.8688 | 1.05[1.00-1.11];0.0738 | 1.01[0.96-1.06];0.6355 | 1.02[0.94-1.10];0.7005 |
| Urea, mmol/L | 1.09[1.09-1.10];<0.0001*** | 1.04[1.01-1.06];0.0032** | 1.03[1.02-1.04];<0.0001*** | 1.01[0.97-1.05];0.6714 | 1.02[1.00-1.04];0.0159* | 1.03[0.99-1.07];0.1218 | 1.05[1.03-1.08];0.0001*** | 1.05[1.01-1.10];0.0253* |
| Protein, g/L | 0.95[0.94-0.96];<0.0001*** | 1.00[0.97-1.02];0.8077 | 1.00[0.99-1.01];0.9893 | 1.01[0.98-1.05];0.4079 | 1.00[0.98-1.02];0.9509 | 1.03[0.99-1.06];0.1062 | 0.97[0.95-1.00];0.0928 | 0.97[0.93-1.02];0.2485 |
| Creatinine, umol/L | 1.003[1.003-1.003];<0.0001*** | 1.001[1.000-1.002];0.1597 | 1.001[1.000-1.001];0.0020** | 1.000[0.998-1.002];0.8419 | 1.001[1.000-1.002];0.0092** | 0.999[0.995-1.002];0.3982 | 1.002[1.001-1.003];0.0012** | 1.001[1.000-1.003];0.1545 |
| Alkaline phosphatase, U/L | 1.01[1.00-1.01];<0.0001*** | 1.00[1.00-1.01];0.0005*** | 1.002[1.001-1.004];0.0077** | 1.00[0.99-1.01];0.9992 | 1.003[1.000-1.005];0.0253* | 1.00[1.00-1.01];0.0007*** | 1.00[0.99-1.00];0.5005 | 1.00[0.99-1.01];0.6006 |
| Aspartate transaminase, U/L | 1.000[0.999-1.001];0.5629 | 1.001[0.999-1.002];0.2718 | 1.001[1.000-1.001];0.1079 | 0.99[0.98-1.01];0.3282 | 1.001[1.000-1.002];0.0062** | 1.00[0.98-1.01];0.6128 | 0.98[0.96-1.00];0.0517 | 0.97[0.94-1.01];0.1174 |
| Alanine transaminase, U/L | 0.989[0.986-0.992];<0.0001*** | 1.00[0.99-1.00];0.3802 | 1.00[0.99-1.00];0.1821 | 0.99[0.98-1.00];0.1596 | 1.001[0.999-1.003];0.4317 | 1.00[0.99-1.00];0.3132 | 0.97[0.96-0.99];0.0003*** | 0.96[0.94-0.99];0.0057** |
| Bilirubin, umol/L | 0.99[0.99-1.00];0.1957 | 1.01[1.00-1.02];0.1809 | 1.00[1.00-1.01];0.3569 | 0.94[0.91-0.98];0.0023** | 1.010[1.006-1.014];<0.0001*** | 0.94[0.90-0.98];0.0028** | 0.97[0.94-1.01];0.1098 | 0.99[0.95-1.04];0.7998 |
| Lipid profiles |  |  |  |  |  |  |  |  |
| Triglyceride, mmol/L | 0.94[0.91-0.98];0.0015** | 0.75[0.66-0.85];<0.0001*** | 0.89[0.85-0.95];0.0001*** | 0.85[0.73-0.99];0.0337* | 0.86[0.79-0.93];0.0003*** | 1.02[0.94-1.10];0.6599 | 0.92[0.81-1.04];0.1798 | 0.75[0.57-0.99];0.0424* |
| SD of triglyceride | 0.92[0.84-1.00];0.0524 | 0.39[0.22-0.67];0.0008*** | 0.69[0.57-0.84];0.0002*** | 0.75[0.47-1.18];0.2073 | 0.53[0.38-0.74];0.0002*** | 0.86[0.58-1.26];0.4390 | 0.92[0.70-1.22];0.5833 | 0.51[0.18-1.44];0.2033 |
| Low-density lipoprotein, mmol/L | 0.99[0.94-1.05];0.7230 | 0.84[0.73-0.97];0.0140* | 0.87[0.80-0.94];0.0003*** | 0.77[0.63-0.93];0.0087** | 0.85[0.77-0.95];0.0030** | 1.09[0.90-1.32];0.3861 | 0.86[0.71-1.03];0.1012 | 0.62[0.45-0.86];0.0037** |
| SD of low-density lipoprotein | 1.36[1.15-1.59];0.0002*** | 0.94[0.58-1.52];0.8110 | 0.83[0.64-1.08];0.1650 | 0.55[0.25-1.19];0.1285 | 0.72[0.50-1.04];0.0836 | 1.19[0.62-2.28];0.5978 | 1.00[0.55-1.81];0.9919 | 0.16[0.03-0.84];0.0307* |
| High-density lipoprotein, mmol/L | 1.01[0.89-1.16];0.8242 | 1.04[0.76-1.43];0.7994 | 1.05[0.88-1.25];0.6076 | 1.16[0.76-1.79];0.4929 | 0.94[0.74-1.20];0.6289 | 1.44[0.92-2.25];0.1122 | 1.06[0.70-1.61];0.7856 | 0.88[0.43-1.77];0.7164 |
| SD of high-density lipoprotein | 10.65[6.14-18.49];<0.0001*** | 4.70[0.89-24.89];0.0691 | 2.09[0.78-5.59];0.1420 | 10.47[1.45-75.48];0.0198* | 0.92[0.21-3.98];0.9161 | 2.48[0.17-35.52];0.5037 | 4.08[0.48-35.09];0.1999 | 3.42[0.08-148.65];0.5230 |
| Total cholesterol, mmol/L | 0.96[0.92-1.01];0.1038 | 0.77[0.69-0.87];<0.0001*** | 0.87[0.81-0.92];<0.0001*** | 0.79[0.67-0.93];0.0046** | 0.82[0.75-0.89];<0.0001*** | 1.16[1.02-1.32];0.0291* | 0.86[0.74-1.00];0.0563 | 0.60[0.45-0.79];0.0003*** |
| SD of total cholesterol | 1.28[1.16-1.41];<0.0001*** | 0.85[0.56-1.28];0.4306 | 0.89[0.72-1.10];0.2866 | 0.91[0.53-1.56];0.7300 | 0.70[0.50-0.96];0.0281* | 1.13[0.70-1.81];0.6227 | 1.13[0.75-1.72];0.5572 | 0.33[0.09-1.16];0.0839 |
| Hemoglobin A1C, % | 0.98[0.95-1.01];0.1398 | 0.91[0.84-0.98];0.0112* | 0.91[0.88-0.95];<0.0001*** | 0.90[0.81-1.00];0.0402* | 0.91[0.86-0.97];0.0015** | 0.99[0.90-1.10];0.8759 | 0.89[0.80-0.99];0.0276* | 0.88[0.75-1.04];0.1320 |
| SD of hemoglobin A1C | 1.15[1.12-1.19];<0.0001*** | 1.06[0.92-1.22];0.4422 | 0.91[0.80-1.03];0.1306 | 0.69[0.47-1.03];0.0719 | 0.83[0.69-1.00];0.0551 | 1.04[0.83-1.30];0.7257 | 1.06[0.87-1.28];0.5589 | 1.08[0.82-1.42];0.5970 |
| Fasting glucose, mmol/L | 1.05[1.04-1.06];<0.0001*** | 0.98[0.94-1.02];0.2979 | 0.98[0.96-1.00];0.1137 | 0.90[0.83-0.98];0.0130* | 0.98[0.95-1.01];0.2576 | 1.01[0.96-1.06];0.7726 | 0.99[0.94-1.05];0.7856 | 0.96[0.86-1.07];0.4892 |
| SD of fasting glucose | 1.10[1.09-1.12];<0.0001*** | 1.04[0.99-1.10];0.1124 | 1.01[0.97-1.04];0.6297 | 0.97[0.87-1.07];0.5126 | 1.00[0.95-1.05];0.9929 | 1.05[0.97-1.13];0.2290 | 1.04[0.96-1.12];0.3451 | 1.07[0.97-1.19];0.1897 |

**Supplementary Table 4B. Univariate Cox regression models to predict primary and secondary cancer outcomes after propensity score matching.**

* for p≤ 0.05, ** for p ≤ 0.01, *** for p ≤ 0.001; HR: hazard ratio; CI: confidence interval; SD: standard deviation; SGLT2I: sodium glucose cotransporter-2 inhibitor; DPP4I: dipeptidyl peptidase-4 inhibitor; MDRD: modification of diet in renal disease.

| **Characteristics** | **All.cause.mortality HR [95% CI];P value** | **Cancer.related.mortality HR [95% CI];P value** | **New.onset.cancer HR [95% CI];P value** | **New.onset.lung.cancer HR [95% CI];P value** | **New.onset.gastrointestinal.cancer HR [95% CI];P value** | **New.onset.breast.cancer HR [95% CI];P value** | **New.onset.genitourinary.cancer HR [95% CI];P value** | **New.onset.bladder.cancer HR [95% CI];P value** |
| --- | --- | --- | --- | --- | --- | --- | --- | --- |
| Demographics |  |  |  |  |  |  |  |  |
| Male gender | 1.20[1.05-1.37];0.0061** | 0.83[0.63-1.09];0.1885 | 0.83[0.71-0.97];0.0158* | 1.89[1.26-2.84];0.0021** | 1.01[0.81-1.26];0.9332 | 0.14[0.08-0.23];<0.0001*** | 0.90[0.62-1.29];0.5505 | 4.63[1.97-10.86];0.0004*** |
| Female gender | 1.0[Reference] | 1.0[Reference] | 1.0[Reference] | 1.0[Reference] | 1.0[Reference] | 1.0[Reference] | 1.0[Reference] | 1.0[Reference] |
| Baseline age, years | 1.09[1.09-1.10];<0.0001*** | 1.08[1.07-1.09];<0.0001*** | 1.05[1.04-1.06];<0.0001*** | 1.07[1.05-1.08];<0.0001*** | 1.07[1.05-1.08];<0.0001*** | 1.02[1.01-1.04];0.0095** | 1.02[1.00-1.04];0.0121* | 1.02[1.00-1.05];0.1105 |
| 18-50 | 0.20[0.15-0.27];<0.0001*** | 0.16[0.08-0.32];<0.0001*** | 0.30[0.22-0.40];<0.0001*** | 0.07[0.02-0.27];0.0001*** | 0.29[0.19-0.45];<0.0001*** | 0.21[0.08-0.51];0.0006*** | 0.61[0.36-1.03];0.0669 | 1.27[0.66-2.43];0.4724 |
| 50-60 | 1.0[Reference] | 1.0[Reference] | 1.0[Reference] | 1.0[Reference] | 1.0[Reference] | 1.0[Reference] | 1.0[Reference] | 1.0[Reference] |
| 60-70 | 1.19[1.04-1.35];0.0117* | 1.43[1.08-1.88];0.0116* | 1.58[1.36-1.85];<0.0001*** | 1.62[1.14-2.32];0.0079** | 1.98[1.59-2.46];<0.0001*** | 0.95[0.62-1.45];0.8084 | 1.24[0.85-1.80];0.2603 | 1.38[0.78-2.44];0.2734 |
| 70-80 | 3.53[3.07-4.06];<0.0001*** | 3.28[2.42-4.45];<0.0001*** | 2.24[1.85-2.70];<0.0001*** | 2.33[1.51-3.60];0.0001*** | 2.69[2.08-3.48];<0.0001*** | 1.54[0.89-2.67];0.1216 | 1.53[0.93-2.53];0.0949 | 2.19[1.10-4.38];0.0264* |
| >80 | 7.93[6.66-9.45];<0.0001*** | 4.84[3.09-7.60];<0.0001*** | 2.47[1.78-3.44];<0.0001*** | 2.92[1.43-5.99];0.0033** | 2.93[1.89-4.57];<0.0001*** | 0.43[0.06-3.08];0.4005 | 2.58[1.20-5.54];0.0149* | 3.65[1.32-10.15];0.0129* |
| Past comorbidities |  |  |  |  |  |  |  |  |
| Charlson standard comorbidity index | 1.75[1.70-1.80];<0.0001*** | 1.59[1.48-1.72];<0.0001*** | 1.41[1.34-1.49];<0.0001*** | 1.53[1.38-1.70];<0.0001*** | 1.51[1.41-1.61];<0.0001*** | 1.11[0.95-1.31];0.1901 | 1.22[1.06-1.40];0.0053** | 1.23[1.00-1.52];0.0537 |
| Duration from earliest diabetes mellitus date to baseline date, day | 1.000[1.000-1.000];<0.0001*** | 1.000[0.999-1.000];0.0007*** | 1.000[1.000-1.000];0.0001*** | 1.000[1.000-1.000];0.0654 | 1.000[1.000-1.000];0.0064** | 1.000[1.000-1.000];0.1981 | 1.000[1.000-1.000];0.0529 | 1.000[0.999-1.000];0.0767 |
| Hypertension | 1.55[1.35-1.77];<0.0001*** | 1.10[0.81-1.51];0.5381 | 1.08[0.91-1.29];0.3644 | 0.81[0.52-1.26];0.3530 | 1.23[0.96-1.58];0.0948 | 1.05[0.67-1.67];0.8246 | 1.06[0.70-1.61];0.7754 | 0.74[0.36-1.52];0.4135 |
| Hyperlipidaemia | 0.46[0.28-0.76];0.0021** | 1.08[0.53-2.20];0.8234 | 0.71[0.44-1.15];0.1653 | 0.00[0.00-Inf];0.9911 | 0.88[0.47-1.64];0.6791 | 0.84[0.27-2.66];0.7711 | 0.70[0.22-2.21];0.5444 | 0.56[0.08-4.08];0.5703 |
| Hypotension | 3.10[1.75-5.47];0.0001*** | 1.18[0.17-8.43];0.8671 | 1.86[0.77-4.47];0.1683 | 2.01[0.28-14.37];0.4876 | 0.76[0.11-5.42];0.7850 | 0.00[0.00-Inf];0.9935 | 4.19[1.04-16.95];0.0445* | 10.42[2.53-42.86];0.0012** |
| Overweight, obesity and hyperalimentation | 0.32[0.13-0.76];0.0105* | 0.29[0.04-2.09];0.2202 | 0.74[0.37-1.49];0.4011 | 0.00[0.00-Inf];0.9907 | 0.77[0.29-2.07];0.6047 | 0.62[0.09-4.43];0.6316 | 1.04[0.26-4.21];0.9565 | 1.26[0.17-9.15];0.8165 |
| Gout | 1.81[1.31-2.52];0.0004*** | 1.57[0.74-3.33];0.2417 | 1.18[0.73-1.91];0.4947 | 1.13[0.36-3.56];0.8333 | 1.45[0.77-2.72];0.2479 | 0.46[0.06-3.28];0.4362 | 1.16[0.37-3.65];0.7998 | 0.93[0.13-6.74];0.9428 |
| Heart failure | 4.05[3.25-5.05];<0.0001*** | 0.80[0.30-2.16];0.6665 | 1.00[0.61-1.65];0.9905 | 0.67[0.17-2.73];0.5806 | 1.04[0.51-2.10];0.9154 | 0.84[0.21-3.40];0.8041 | 1.04[0.33-3.29];0.9403 | 0.84[0.12-6.07];0.8607 |
| Acute myocardial infarction | 2.27[1.78-2.90];<0.0001*** | 1.00[0.47-2.13];0.9930 | 0.80[0.50-1.28];0.3512 | 0.48[0.12-1.93];0.3002 | 1.22[0.70-2.12];0.4853 | 0.00[0.00-Inf];0.9922 | 1.00[0.37-2.70];0.9931 | 1.21[0.29-4.99];0.7890 |
| Ischemic heart disease | 1.48[1.26-1.75];<0.0001*** | 1.00[0.67-1.51];0.9854 | 0.92[0.73-1.17];0.4992 | 1.14[0.68-1.90];0.6244 | 1.18[0.86-1.61];0.2987 | 0.29[0.11-0.80];0.0166* | 0.71[0.38-1.33];0.2869 | 0.79[0.32-2.00];0.6244 |
| Peripheral vascular disease | 6.06[4.19-8.77];<0.0001*** | 0.93[0.13-6.66];0.9450 | 0.57[0.14-2.28];0.4263 | 0.00[0.00-Inf];0.9918 | 0.59[0.08-4.22];0.6009 | 0.00[0.00-Inf];0.9927 | 0.00[0.00-Inf];0.9919 | 0.00[0.00-Inf];0.9948 |
| Stroke/transient ischemic attack | 2.81[2.19-3.61];<0.0001*** | 0.74[0.28-2.00];0.5566 | 1.11[0.70-1.75];0.6645 | 1.27[0.47-3.44];0.6360 | 1.46[0.82-2.61];0.1946 | 0.38[0.05-2.74];0.3394 | 0.32[0.04-2.27];0.2528 | 0.00[0.00-Inf];0.9952 |
| Atrial fibrillation | 3.09[2.37-4.02];<0.0001*** | 1.40[0.62-3.15];0.4187 | 1.38[0.87-2.17];0.1686 | 0.38[0.05-2.75];0.3411 | 1.50[0.80-2.82];0.2043 | 2.49[1.01-6.12];0.0467* | 0.79[0.20-3.21];0.7452 | 1.97[0.48-8.10];0.3476 |
| Diabetic eye disease | 1.37[1.10-1.71];0.0044** | 1.36[0.85-2.19];0.1970 | 0.92[0.67-1.25];0.5810 | 0.46[0.17-1.24];0.1260 | 1.25[0.84-1.85];0.2679 | 0.28[0.07-1.13];0.0734 | 0.98[0.48-2.00];0.9487 | 1.20[0.43-3.33];0.7263 |
| Alcohol dependence | 7.55[3.76-15.13];<0.0001*** | 12.95[4.14-40.49];<0.0001*** | 5.17[1.93-13.80];0.0011** | 0.00[0.00-Inf];0.9941 | 10.78[4.02-28.91];<0.0001*** | 0.00[0.00-Inf];0.9947 | 0.00[0.00-Inf];0.9942 | 0.00[0.00-Inf];0.9962 |
| Chronic liver disease and cirrhosis | 0.94[0.63-1.40];0.7525 | 0.69[0.25-1.84];0.4542 | 1.20[0.78-1.83];0.4080 | 0.29[0.04-2.06];0.2155 | 1.96[1.20-3.19];0.0069** | 0.36[0.05-2.55];0.3032 | 0.90[0.29-2.84];0.8592 | 0.72[0.10-5.24];0.7487 |
| Viral hepatitis | 0.54[0.24-1.19];0.1267 | 0.00[0.00-Inf];0.9897 | 1.44[0.79-2.61];0.2300 | 0.70[0.10-5.03];0.7254 | 2.77[1.47-5.19];0.0015** | 0.00[0.00-Inf];0.9929 | 0.00[0.00-Inf];0.9922 | 0.00[0.00-Inf];0.9950 |
| History of acute liver injury | 2.67[1.11-6.44];0.0283* | 0.00[0.00-Inf];0.9900 | 3.88[1.61-9.34];0.0025** | 0.00[0.00-Inf];0.9923 | 8.11[3.35-19.63];<0.0001*** | 0.00[0.00-Inf];0.9931 | 0.00[0.00-Inf];0.9924 | 0.00[0.00-Inf];0.9951 |
| Other liver disease | 1.62[0.96-2.75];0.0719 | 1.06[0.26-4.28];0.9316 | 2.56[1.53-4.27];0.0003*** | 0.90[0.13-6.47];0.9201 | 4.31[2.42-7.68];<0.0001*** | 1.12[0.16-8.00];0.9128 | 0.00[0.00-Inf];0.9931 | 0.00[0.00-Inf];0.9956 |
| Autoimmune disease tissue | 1.16[0.66-2.05];0.6039 | 0.89[0.22-3.58];0.8675 | 0.83[0.37-1.86];0.6572 | 0.75[0.11-5.39];0.7781 | 1.16[0.43-3.11];0.7699 | 0.93[0.13-6.65];0.9404 | 0.00[0.00-Inf];0.9925 | 0.00[0.00-Inf];0.9952 |
| Other carcinogens pathogen | 0.00[0.00-Inf];0.9794 | 0.00[0.00-Inf];0.9904 | 37.59[15.59-90.60];<0.0001*** | 210.39[85.97-514.90];<0.0001*** | 0.00[0.00-Inf];0.9882 | 0.00[0.00-Inf];0.9934 | 0.00[0.00-Inf];0.9928 | 0.00[0.00-Inf];0.9954 |
| Chronic obstructive pulmonary disease | 8.84[3.67-21.28];<0.0001*** | 0.00[0.00-Inf];0.9916 | 9.69[3.63-25.90];<0.0001*** | 54.27[20.04-146.97];<0.0001*** | 0.00[0.00-Inf];0.9893 | 0.00[0.00-Inf];0.9941 | 0.00[0.00-Inf];0.9934 | 0.00[0.00-Inf];0.9958 |
| Gastrointestinal disease | 1.83[1.29-2.59];0.0006*** | 1.01[0.37-2.71];0.9891 | 1.77[1.16-2.71];0.0083** | 3.14[1.46-6.72];0.0033** | 1.31[0.65-2.65];0.4453 | 1.05[0.26-4.27];0.9415 | 2.24[0.92-5.49];0.0769 | 4.53[1.63-12.57];0.0038** |
| Medications |  |  |  |  |  |  |  |  |
| SGLT2I v.s. DPP4I | 0.80[0.70-0.90];0.0004*** | 0.54[0.41-0.71];<0.0001*** | 0.56[0.48-0.65];<0.0001*** | 0.51[0.35-0.74];0.0004*** | 0.65[0.52-0.82];0.0002*** | 0.46[0.30-0.70];0.0003*** | 0.47[0.32-0.69];0.0001*** | 0.28[0.14-0.55];0.0002*** |
| SGLT2I frequency | 1.02[1.01-1.02];<0.0001*** | 1.02[1.01-1.03];<0.0001*** | 1.02[1.01-1.02];<0.0001*** | 1.01[1.00-1.03];0.0949 | 1.02[1.01-1.02];<0.0001*** | 1.01[1.00-1.03];0.1015 | 1.02[1.01-1.03];0.0016** | 1.02[0.99-1.04];0.2204 |
| DPP4I frequency | 1.01[1.00-1.02];0.0299* | 1.01[1.00-1.03];0.1476 | 1.01[1.01-1.02];0.0013** | 0.97[0.94-1.01];0.1294 | 1.02[1.00-1.03];0.0065** | 1.03[1.02-1.05];<0.0001*** | 0.99[0.95-1.02];0.3935 | 0.99[0.95-1.04];0.7322 |
| SGLT2I duration, days | 1.000[1.000-1.000];0.0009*** | 1.000[1.000-1.000];0.9170 | 1.000[1.000-1.000];0.3467 | 1.000[1.000-1.000];0.9313 | 1.000[1.000-1.000];0.7998 | 1.000[1.000-1.001];0.5141 | 1.000[1.000-1.001];0.3931 | 1.000[1.000-1.001];0.3645 |
| DPP4I duration, days | 0.999[0.999-0.999];<0.0001*** | 0.999[0.999-1.000];0.0046** | 0.999[0.999-1.000];<0.0001*** | 1.000[0.999-1.000];0.1331 | 0.999[0.999-1.000];0.0002*** | 1.000[1.000-1.001];0.1612 | 0.999[0.998-0.999];<0.0001*** | 0.997[0.996-0.998];<0.0001*** |
| Metformin | 0.42[0.35-0.51];<0.0001*** | 0.97[0.56-1.67];0.9134 | 0.68[0.53-0.89];0.0041** | 0.71[0.38-1.33];0.2879 | 0.68[0.47-0.99];0.0458* | 1.10[0.48-2.50];0.8247 | 0.49[0.29-0.84];0.0100* | 0.51[0.22-1.20];0.1226 |
| Sulphonylurea | 1.24[1.07-1.44];0.0047** | 1.48[1.06-2.07];0.0222* | 1.19[0.99-1.41];0.0578 | 1.97[1.22-3.17];0.0056** | 1.24[0.96-1.60];0.1024 | 1.44[0.89-2.33];0.1364 | 0.59[0.41-0.86];0.0053** | 0.52[0.30-0.91];0.0230* |
| Insulin | 4.69[3.96-5.56];<0.0001*** | 4.05[2.86-5.74];<0.0001*** | 2.04[1.73-2.40];<0.0001*** | 1.26[0.88-1.81];0.1982 | 3.16[2.44-4.10];<0.0001*** | 0.80[0.54-1.18];0.2572 | 3.03[1.99-4.63];<0.0001*** | 3.23[1.66-6.31];0.0006*** |
| Acarbose | 1.42[1.07-1.87];0.0139* | 0.97[0.48-1.96];0.9225 | 1.10[0.76-1.60];0.6113 | 0.61[0.19-1.91];0.3937 | 0.94[0.53-1.67];0.8285 | 2.11[1.02-4.34];0.0431* | 1.28[0.56-2.91];0.5560 | 1.02[0.25-4.20];0.9767 |
| Thiozolidinedone | 0.51[0.43-0.61];<0.0001*** | 0.84[0.61-1.15];0.2768 | 0.62[0.51-0.76];<0.0001*** | 0.60[0.38-0.95];0.0297* | 0.67[0.51-0.88];0.0039** | 0.64[0.39-1.06];0.0828 | 0.49[0.29-0.80];0.0048** | 0.31[0.12-0.78];0.0128* |
| Glucagon-like peptide-1 receptor agonists | 0.24[0.15-0.37];<0.0001*** | 0.23[0.08-0.61];0.0035** | 0.31[0.19-0.50];<0.0001*** | 0.20[0.05-0.79];0.0219* | 0.26[0.12-0.55];0.0004*** | 0.36[0.12-1.15];0.0844 | 0.30[0.10-0.95];0.0410* | 0.24[0.03-1.76];0.1617 |
| ACEI/ARB | 1.27[1.11-1.46];0.0004*** | 0.76[0.58-1.00];0.0512 | 1.06[0.90-1.23];0.4964 | 0.87[0.61-1.24];0.4330 | 1.21[0.96-1.52];0.1060 | 0.66[0.45-0.98];0.0401* | 1.18[0.81-1.71];0.3907 | 1.02[0.58-1.81];0.9410 |
| Antidepressants | 2.06[1.73-2.45];<0.0001*** | 2.90[2.08-4.06];<0.0001*** | 1.58[1.26-1.99];0.0001*** | 0.99[0.52-1.89];0.9757 | 1.72[1.25-2.37];0.0009*** | 2.79[1.71-4.56];<0.0001*** | 1.02[0.53-1.94];0.9640 | 0.23[0.03-1.66];0.1456 |
| Antihypertensive drugs | 1.55[1.28-1.87];<0.0001*** | 0.96[0.58-1.58];0.8727 | 0.71[0.52-0.97];0.0316* | 0.56[0.24-1.26];0.1608 | 0.83[0.54-1.27];0.3973 | 0.22[0.05-0.90];0.0345* | 0.77[0.38-1.58];0.4831 | 1.21[0.48-3.06];0.6800 |
| Antihepatitis | 0.61[0.34-1.10];0.1008 | 0.77[0.25-2.40];0.6481 | 2.35[1.61-3.43];<0.0001*** | 0.43[0.06-3.11];0.4070 | 4.72[3.16-7.04];<0.0001*** | 0.00[0.00-Inf];0.9910 | 0.00[0.00-Inf];0.9902 | 0.00[0.00-Inf];0.9937 |
| Anticoagulants | 0.01[0.00-0.03];<0.0001*** | 0.00[0.00-0.03];<0.0001*** | 8103.75[0.00-Inf];0.9879 | 8103.76[0.00-Inf];0.9947 | 8103.74[0.00-Inf];0.9915 | 8103.75[0.00-Inf];0.9962 | 8103.76[0.00-Inf];0.9944 | 8103.75[0.00-Inf];0.9963 |
| Antiplatelets | 2.19[1.93-2.49];<0.0001*** | 0.79[0.58-1.07];0.1257 | 0.94[0.80-1.11];0.4706 | 1.34[0.94-1.93];0.1100 | 1.13[0.90-1.42];0.2825 | 0.29[0.16-0.52];<0.0001*** | 0.83[0.56-1.23];0.3532 | 0.75[0.40-1.40];0.3618 |
| Statins and fibrates | 1.13[0.97-1.32];0.1067 | 0.88[0.64-1.19];0.3948 | 1.04[0.87-1.24];0.6920 | 0.97[0.64-1.46];0.8822 | 1.49[1.12-1.99];0.0067** | 0.45[0.30-0.67];0.0001*** | 1.13[0.73-1.74];0.5787 | 0.66[0.36-1.19];0.1663 |
| Nitrates | 1.84[1.59-2.14];<0.0001*** | 0.80[0.53-1.21];0.2880 | 0.95[0.76-1.18];0.6209 | 0.88[0.52-1.48];0.6224 | 1.26[0.95-1.68];0.1144 | 0.31[0.13-0.76];0.0104* | 1.03[0.63-1.70];0.9013 | 0.81[0.34-1.89];0.6196 |
| Non-steroidal anti-inflammatory drugs | 2.15[1.90-2.44];<0.0001*** | 0.76[0.56-1.04];0.0862 | 0.93[0.78-1.09];0.3642 | 1.40[0.97-2.01];0.0693 | 1.10[0.87-1.39];0.4117 | 0.22[0.11-0.43];<0.0001*** | 0.86[0.58-1.28];0.4657 | 0.78[0.41-1.46];0.4311 |
| Diuretics | 2.78[2.45-3.15];<0.0001*** | 1.17[0.88-1.56];0.2822 | 1.17[1.00-1.37];0.0568 | 1.22[0.84-1.77];0.2919 | 1.19[0.95-1.50];0.1334 | 0.68[0.43-1.08];0.1032 | 1.56[1.09-2.24];0.0161* | 1.53[0.87-2.69];0.1408 |
| Beta-blockers | 1.98[1.74-2.25];<0.0001*** | 0.87[0.63-1.20];0.3871 | 0.88[0.74-1.06];0.1804 | 0.90[0.59-1.36];0.6105 | 0.93[0.72-1.20];0.5676 | 0.68[0.41-1.12];0.1341 | 0.97[0.64-1.46];0.8763 | 0.83[0.42-1.62];0.5811 |
| Calcium channel blockers | 1.82[1.60-2.07];<0.0001*** | 1.43[1.09-1.87];0.0103* | 1.52[1.30-1.76];<0.0001*** | 1.86[1.29-2.66];0.0008*** | 1.67[1.34-2.08];<0.0001*** | 0.94[0.63-1.39];0.7422 | 1.41[0.98-2.01];0.0620 | 1.03[0.59-1.80];0.9151 |
| SUbclinical biomarkers |  |  |  |  |  |  |  |  |
| Abbreviated MDRD, mL/min/1.73m^2 | 0.971[0.968-0.974];<0.0001*** | 0.99[0.98-0.99];0.0001*** | 0.989[0.985-0.993];<0.0001*** | 0.99[0.98-1.00];0.1430 | 0.99[0.98-0.99];<0.0001*** | 1.00[0.99-1.01];0.4011 | 0.98[0.97-0.99];<0.0001*** | 0.97[0.96-0.99];0.0004*** |
| Most severe renal damage (<15 mL/min/1.73m^2) | 5.93[2.66-13.24];<0.0001*** | 0.00[0.00-Inf];0.9928 | 4.35[1.40-13.52];0.0111* | 9.23[1.28-66.30];0.0272* | 0.00[0.00-Inf];0.9906 | 0.00[0.00-Inf];0.9949 | 17.78[4.38-72.23];0.0001*** | 0.00[0.00-Inf];0.9968 |
| Severe renal damage ([15, 30) mL/min/1.73m^2) | 8.85[5.85-13.41];<0.0001*** | 1.76[0.25-12.59];0.5718 | 2.13[0.80-5.70];0.1314 | 3.40[0.47-24.39];0.2244 | 2.06[0.51-8.29];0.3079 | 0.00[0.00-Inf];0.9945 | 3.19[0.44-22.92];0.2484 | 0.00[0.00-Inf];0.9965 |
| Moderate to severe renal damage ([30, 45) mL/min/1.73m^2) | 4.62[3.63-5.89];<0.0001*** | 1.41[0.58-3.44];0.4471 | 1.30[0.78-2.18];0.3126 | 1.68[0.53-5.32];0.3767 | 1.53[0.79-2.97];0.2107 | 1.71[0.54-5.43];0.3596 | 1.04[0.26-4.23];0.9555 | 0.00[0.00-Inf];0.9964 |
| Mild to moderate renal damage ([45, 60) mL/min/1.73m^2) | 2.53[2.10-3.05];<0.0001*** | 1.61[1.00-2.59];0.0520 | 1.55[1.18-2.03];0.0015** | 1.50[0.75-3.00];0.2486 | 1.41[0.96-2.09];0.0835 | 0.80[0.33-1.98];0.6348 | 2.54[1.46-4.42];0.0010** | 4.00[1.80-8.87];0.0006*** |
| Mild renal damage ([60, 90] mL/min/1.73m^2) | 1.07[0.93-1.23];0.3405 | 1.48[1.10-2.01];0.0109* | 1.21[1.02-1.43];0.0300* | 0.68[0.43-1.06];0.0888 | 1.55[1.22-1.97];0.0003*** | 0.85[0.55-1.32];0.4600 | 1.44[0.95-2.18];0.0875 | 1.36[0.69-2.70];0.3725 |
| Chronic kidney disease (>90 mL/min/1.73m^2) | 0.39[0.33-0.45];<0.0001*** | 0.53[0.39-0.74];0.0002*** | 0.67[0.56-0.80];<0.0001*** | 1.11[0.72-1.71];0.6276 | 0.53[0.41-0.68];<0.0001*** | 1.19[0.78-1.83];0.4220 | 0.38[0.24-0.62];0.0001*** | 0.37[0.17-0.83];0.0153* |
| Neutrophil-to-lymphocyte ratio | 1.04[1.03-1.05];<0.0001*** | 1.02[0.97-1.06];0.4716 | 1.00[0.97-1.04];0.9293 | 0.96[0.83-1.10];0.5438 | 1.02[0.98-1.05];0.3410 | 0.88[0.71-1.10];0.2596 | 1.01[0.94-1.09];0.7561 | 0.94[0.72-1.23];0.6567 |
| Platelet-to-lymphocyte ratio | 1.000[1.000-1.001];0.0009*** | 1.000[0.999-1.001];0.5884 | 1.000[0.999-1.001];0.9522 | 1.00[0.99-1.00];0.7390 | 0.999[0.997-1.002];0.5756 | 1.000[0.998-1.002];0.9249 | 1.000[0.999-1.001];0.4607 | 1.00[0.99-1.01];0.5106 |
| Neutrophil-to-high-density lipoprotein ratio | 1.84[1.45-2.34];<0.0001*** | 1.11[0.27-4.48];0.8876 | 0.57[0.21-1.56];0.2764 | 1.14[0.17-7.48];0.8910 | 0.80[0.21-2.96];0.7336 | 0.10[0.00-2.35];0.1539 | 0.74[0.07-8.14];0.8091 | 0.42[0.00-34.98];0.6975 |
| Low density lipoprotein ratio-to-high density lipoprotein ratio | 1.03[0.95-1.13];0.4674 | 0.89[0.73-1.08];0.2304 | 0.84[0.75-0.94];0.0019** | 0.90[0.68-1.19];0.4585 | 0.79[0.68-0.93];0.0039** | 0.94[0.71-1.23];0.6381 | 0.77[0.58-1.02];0.0704 | 0.86[0.56-1.34];0.5194 |
| Triglyceride-glucose index | 0.95[0.85-1.07];0.4086 | 0.73[0.57-0.93];0.0119* | 0.81[0.70-0.92];0.0017** | 0.82[0.58-1.16];0.2685 | 0.76[0.63-0.92];0.0040** | 1.10[0.80-1.51];0.5596 | 0.72[0.51-1.02];0.0672 | 0.70[0.38-1.28];0.2467 |
| Protein-to-creatinine ratio | 0.66[0.61-0.71];<0.0001*** | 0.93[0.80-1.08];0.3140 | 0.93[0.85-1.01];0.0921 | 1.06[0.87-1.28];0.5598 | 0.72[0.63-0.84];<0.0001*** | 1.30[1.15-1.48];<0.0001*** | 0.83[0.66-1.04];0.1053 | 0.81[0.54-1.21];0.3000 |
| Aspartate aminotransferase-to-alanine transaminase ratio | 1.06[0.97-1.14];0.1810 | 1.02[0.69-1.51];0.9266 | 1.07[0.97-1.17];0.1807 | 0.80[0.25-2.57];0.7121 | 1.07[0.94-1.21];0.3370 | 1.07[0.82-1.41];0.6117 | 1.08[0.95-1.24];0.2420 | 1.10[0.90-1.33];0.3438 |
| Complete blood counts, renal and liver functions |  |  |  |  |  |  |  |  |
| Mean corpuscular volume, fL | 1.01[1.00-1.02];0.0826 | 1.02[0.99-1.06];0.1566 | 1.02[1.00-1.03];0.0758 | 1.04[0.99-1.09];0.1171 | 1.01[0.99-1.03];0.4330 | 1.04[0.99-1.09];0.0804 | 1.00[0.96-1.05];0.8295 | 1.01[0.94-1.09];0.7520 |
| Eosinophil, x10^9/L | 0.54[0.32-0.93];0.0266* | 0.51[0.12-2.16];0.3578 | 1.47[1.03-2.10];0.0320* | 2.02[1.44-2.84];<0.0001*** | 0.42[0.13-1.39];0.1562 | 0.10[0.01-1.28];0.0762 | 1.99[1.41-2.82];0.0001*** | 2.20[1.48-3.29];0.0001*** |
| Lymphocyte, x10^9/L | 0.54[0.47-0.61];<0.0001*** | 0.78[0.57-1.08];0.1395 | 0.79[0.66-0.95];0.0105* | 1.03[0.70-1.50];0.8877 | 0.65[0.50-0.85];0.0016** | 0.96[0.63-1.44];0.8299 | 0.94[0.61-1.45];0.7852 | 1.23[0.83-1.83];0.3094 |
| Neutrophil, x10^9/L | 1.07[1.05-1.08];<0.0001*** | 1.01[0.93-1.10];0.7868 | 0.96[0.90-1.02];0.1483 | 1.00[0.88-1.14];0.9603 | 0.97[0.90-1.06];0.5227 | 0.87[0.72-1.05];0.1380 | 0.96[0.83-1.12];0.6247 | 0.95[0.74-1.23];0.7068 |
| White cell count, x10^9/L | 1.02[1.01-1.03];0.0010** | 1.01[0.97-1.06];0.5387 | 0.97[0.93-1.02];0.2882 | 1.02[0.99-1.06];0.2274 | 0.94[0.88-1.01];0.1187 | 0.88[0.77-1.01];0.0796 | 1.02[0.98-1.06];0.3821 | 1.02[0.97-1.08];0.4042 |
| Platelet, x10^9/L | 0.997[0.996-0.998];<0.0001*** | 1.000[0.997-1.003];0.8176 | 0.999[0.997-1.001];0.2420 | 1.00[1.00-1.01];0.0741 | 0.99[0.99-1.00];<0.0001*** | 1.00[1.00-1.01];0.0576 | 1.00[1.00-1.01];0.3911 | 1.00[0.99-1.01];0.9758 |
| Red cell count, x10^12/L | 0.49[0.42-0.56];<0.0001*** | 0.61[0.43-0.86];0.0048** | 0.52[0.43-0.63];<0.0001*** | 0.52[0.32-0.86];0.0115* | 0.53[0.40-0.70];<0.0001*** | 0.34[0.21-0.53];<0.0001*** | 0.69[0.43-1.12];0.1364 | 1.38[0.64-3.00];0.4127 |
| Potassium, mmol/L | 1.13[0.96-1.32];0.1388 | 0.98[0.69-1.39];0.9152 | 1.05[0.86-1.27];0.6465 | 1.51[0.95-2.41];0.0819 | 1.16[0.89-1.52];0.2598 | 0.68[0.41-1.14];0.1443 | 0.93[0.58-1.49];0.7617 | 1.38[0.65-2.92];0.4049 |
| Albumin, g/L | 0.85[0.83-0.86];<0.0001*** | 0.91[0.87-0.95];0.0001*** | 0.94[0.91-0.96];<0.0001*** | 0.96[0.89-1.02];0.1989 | 0.91[0.88-0.95];<0.0001*** | 1.03[0.95-1.11];0.5412 | 0.96[0.89-1.03];0.2343 | 0.99[0.87-1.11];0.8196 |
| Sodium, mmol/L | 0.95[0.93-0.97];<0.0001*** | 1.01[0.96-1.07];0.6161 | 1.01[0.97-1.04];0.7326 | 0.95[0.88-1.02];0.1476 | 1.02[0.98-1.07];0.3778 | 1.05[0.97-1.14];0.2413 | 1.00[0.93-1.08];0.9649 | 1.12[0.99-1.27];0.0833 |
| Urea, mmol/L | 1.12[1.10-1.13];<0.0001*** | 1.06[1.01-1.12];0.0105* | 1.07[1.04-1.10];<0.0001*** | 1.04[0.95-1.13];0.3899 | 1.07[1.04-1.11];0.0001*** | 0.98[0.87-1.09];0.6639 | 1.11[1.07-1.15];<0.0001*** | 1.11[1.05-1.18];0.0007*** |
| Protein, g/L | 0.97[0.96-0.99];0.0012** | 1.00[0.97-1.04];0.8575 | 1.02[1.00-1.04];0.0826 | 0.99[0.95-1.04];0.8201 | 1.03[1.00-1.06];0.0606 | 1.07[1.02-1.13];0.0107* | 0.98[0.94-1.03];0.5068 | 0.99[0.91-1.08];0.8466 |
| Creatinine, umol/L | 1.003[1.003-1.004];<0.0001*** | 1.001[0.998-1.004];0.4922 | 1.002[1.001-1.003];0.0006*** | 1.002[1.000-1.005];0.0268* | 1.002[1.000-1.004];0.0354* | 0.96[0.95-0.97];<0.0001*** | 1.003[1.002-1.004];<0.0001*** | 1.00[1.00-1.01];0.0462* |
| Alkaline phosphatase, U/L | 1.00[1.00-1.01];<0.0001*** | 1.00[0.99-1.01];0.9085 | 1.00[1.00-1.01];0.0003*** | 1.00[1.00-1.01];0.0838 | 1.00[1.00-1.01];<0.0001*** | 1.00[1.00-1.01];0.1598 | 0.99[0.98-1.00];0.0684 | 0.97[0.95-1.00];0.0222* |
| Aspartate transaminase, U/L | 0.99[0.99-1.00];0.1371 | 0.99[0.97-1.01];0.2961 | 1.000[0.995-1.005];0.9683 | 1.00[0.99-1.01];0.6548 | 1.00[1.00-1.01];0.3885 | 0.98[0.95-1.01];0.2623 | 0.99[0.97-1.01];0.3658 | 0.97[0.93-1.02];0.2243 |
| Alanine transaminase, U/L | 0.990[0.986-0.995];<0.0001*** | 0.99[0.98-1.00];0.1952 | 1.00[0.99-1.00];0.1041 | 1.00[0.99-1.01];0.9848 | 1.00[0.99-1.01];0.7932 | 0.99[0.97-1.00];0.1384 | 0.98[0.96-1.00];0.0256* | 0.95[0.91-1.00];0.0545 |
| Bilirubin, umol/L | 0.99[0.97-1.00];0.1427 | 0.98[0.95-1.02];0.3739 | 1.00[0.98-1.02];0.7280 | 0.88[0.83-0.94];0.0003*** | 1.02[1.01-1.03];0.0008*** | 0.97[0.91-1.02];0.2281 | 0.97[0.92-1.02];0.1953 | 1.01[0.96-1.07];0.6573 |
| Lipid profiles |  |  |  |  |  |  |  |  |
| Triglyceride, mmol/L | 0.96[0.91-1.01];0.1238 | 0.78[0.65-0.93];0.0068** | 0.91[0.84-0.98];0.0124* | 0.88[0.71-1.08];0.2155 | 0.87[0.77-0.97];0.0175* | 0.98[0.85-1.13];0.7986 | 0.98[0.85-1.13];0.7462 | 0.87[0.62-1.22];0.4128 |
| SD of triglyceride | 0.91[0.79-1.04];0.1537 | 0.22[0.09-0.58];0.0021** | 0.44[0.30-0.64];<0.0001*** | 0.81[0.47-1.41];0.4614 | 0.25[0.13-0.48];<0.0001*** | 0.34[0.11-1.11];0.0734 | 0.80[0.47-1.38];0.4297 | 0.35[0.07-1.84];0.2144 |
| Low-density lipoprotein, mmol/L | 0.96[0.88-1.06];0.4565 | 0.84[0.68-1.04];0.1094 | 0.80[0.71-0.90];0.0002*** | 0.83[0.62-1.11];0.2012 | 0.74[0.63-0.88];0.0005*** | 1.08[0.83-1.42];0.5592 | 0.69[0.51-0.93];0.0163* | 0.65[0.40-1.06];0.0851 |
| SD of low-density lipoprotein | 1.30[0.97-1.74];0.0761 | 0.70[0.32-1.54];0.3722 | 0.78[0.52-1.18];0.2415 | 0.47[0.14-1.57];0.2188 | 0.79[0.45-1.37];0.3948 | 0.59[0.17-2.03];0.4075 | 0.84[0.31-2.29];0.7347 | 0.37[0.05-2.56];0.3143 |
| High-density lipoprotein, mmol/L | 0.79[0.62-1.01];0.0597 | 0.91[0.55-1.52];0.7273 | 1.10[0.84-1.45];0.4790 | 0.99[0.49-1.98];0.9712 | 1.10[0.75-1.61];0.6177 | 1.52[0.79-2.94];0.2067 | 1.00[0.50-1.96];0.9900 | 0.60[0.18-1.99];0.3991 |
| SD of high-density lipoprotein | 3.64[1.11-11.99];0.0334* | 0.00[0.00-0.31];0.0118* | 0.31[0.05-1.83];0.1961 | 0.18[0.00-17.27];0.4606 | 0.99[0.11-9.04];0.9901 | 1.33[0.02-97.25];0.8952 | 0.00[0.00-0.24];0.0158* | 0.01[0.00-43.49];0.2742 |
| Total cholesterol, mmol/L | 0.92[0.85-0.99];0.0309* | 0.78[0.65-0.93];0.0058** | 0.84[0.76-0.92];0.0004*** | 0.82[0.65-1.05];0.1183 | 0.78[0.68-0.89];0.0003*** | 1.13[0.94-1.36];0.2003 | 0.79[0.62-1.00];0.0499* | 0.62[0.41-0.95];0.0284* |
| SD of total cholesterol | 1.19[1.01-1.40];0.0365* | 0.51[0.24-1.07];0.0768 | 0.74[0.53-1.05];0.0923 | 0.69[0.28-1.70];0.4156 | 0.75[0.47-1.19];0.2195 | 0.42[0.14-1.30];0.1317 | 0.87[0.39-1.91];0.7228 | 0.19[0.03-1.41];0.1054 |
| Hemoglobin A1C, % | 1.05[1.01-1.09];0.0086** | 0.97[0.87-1.07];0.5439 | 0.91[0.86-0.97];0.0037** | 0.95[0.82-1.10];0.4877 | 0.95[0.88-1.03];0.2179 | 0.94[0.81-1.10];0.4503 | 0.78[0.66-0.92];0.0040** | 0.70[0.53-0.94];0.0180* |
| SD of hemoglobin A1C | 1.14[1.09-1.20];<0.0001*** | 1.02[0.80-1.29];0.8879 | 0.94[0.79-1.11];0.4574 | 0.91[0.58-1.42];0.6812 | 0.84[0.63-1.12];0.2246 | 1.07[0.83-1.37];0.6075 | 0.96[0.65-1.43];0.8453 | 0.80[0.35-1.83];0.6009 |
| Fasting glucose, mmol/L | 1.06[1.05-1.07];<0.0001*** | 1.00[0.95-1.06];0.9353 | 0.99[0.96-1.03];0.7538 | 0.95[0.87-1.05];0.3575 | 1.01[0.96-1.05];0.7599 | 1.00[0.92-1.08];0.9954 | 0.96[0.86-1.06];0.3921 | 0.92[0.76-1.12];0.4213 |
| SD of fasting glucose | 1.12[1.09-1.14];<0.0001*** | 1.03[0.93-1.13];0.6100 | 1.01[0.96-1.07];0.6787 | 1.06[0.95-1.19];0.3102 | 1.02[0.95-1.10];0.5688 | 1.00[0.86-1.16];0.9858 | 0.86[0.68-1.09];0.2043 | 0.95[0.66-1.36];0.7862 |

**Supplementary Table 5. Sensitivity analyses for SGLT2I v.s. DPP4I exposure effects predict primary and secondary cancer outcomes before and after propensity score matching (1:1).**

*** for p≤ 0.05, ** for p ≤ 0.01, *** for p ≤ 0.001; SGLT2I: Sodium-glucose cotransporter-2 inhibitors; DPP4I: Dipeptidyl peptidase-4 inhibitors; HR: hazard ratio; CI: confidence interval; PS: propensity score; IPTW: inverse probability of treatment weighting, SIPTW: stable inverse probability of treatment weighting.**

| **Before matching** | | | | | | | | |
| --- | --- | --- | --- | --- | --- | --- | --- | --- |
| **Characteristics** | **All.cause.mortality HR [95% CI];P value** | **Cancer.related.mortality HR [95% CI];P value** | **New.onset.cancer HR [95% CI];P value** | **New.onset.lung.cancer HR [95% CI];P value** | **New.onset.gastrointestinal.cancer HR [95% CI];P value** | **New.onset.breast.cancer HR [95% CI];P value** | **New.onset.bladder.cancer HR [95% CI];P value** | **New.onset.genitourinary.cancer HR [95% CI];P value** |
| SGLT2I v.s. DPP4I.1 | 0.44[0.40-0.48];<0.0001*** | 0.66[0.54-0.82];0.0001*** | 0.56[0.49-0.63];<0.0001*** | 0.54[0.40-0.74];0.0001*** | 0.51[0.43-0.61];<0.0001*** | 0.64[0.46-0.89];0.0082** | 0.75[0.47-1.18];0.2152 | 0.64[0.47-0.85];0.0025** |
| Dapagliflozin | 0.50[0.44-0.56];<0.0001*** | 0.77[0.60-0.98];0.0359* | 0.61[0.53-0.72];<0.0001*** | 0.68[0.47-0.99];0.0447* | 0.54[0.44-0.68];<0.0001*** | 0.54[0.34-0.85];0.0077** | 0.92[0.54-1.57];0.7486 | 0.77[0.54-1.09];0.1339 |
| Empagliflozin | 0.42[0.34-0.52];<0.0001*** | 0.63[0.41-0.98];0.0413* | 0.59[0.45-0.76];0.0001*** | 0.49[0.24-0.99];0.0459* | 0.56[0.39-0.81];0.0018** | 0.61[0.30-1.24];0.1703 | 0.63[0.23-1.72];0.3694 | 0.71[0.40-1.27];0.2445 |
| Canagliflozin | 0.48[0.40-0.58];<0.0001*** | 0.60[0.40-0.91];0.0148* | 0.56[0.44-0.71];<0.0001*** | 0.40[0.20-0.81];0.0113* | 0.57[0.41-0.80];0.0010** | 0.91[0.53-1.56];0.7284 | 0.52[0.19-1.42];0.2019 | 0.48[0.26-0.91];0.0239* |
| Ertugliflozin | 0.55[0.44-0.70];<0.0001*** | 0.54[0.31-0.97];0.0377* | 0.48[0.34-0.68];<0.0001*** | 0.46[0.19-1.12];0.0871 | 0.51[0.32-0.81];0.0043** | 0.81[0.38-1.73];0.5899 | 0.23[0.03-1.68];0.1491 | 0.26[0.08-0.82];0.0210* |
| **After matching** | | | | | | | | |
| **Characteristics** | **All.cause.mortality HR [95% CI];P value** | **Cancer.related.mortality HR [95% CI];P value** | **New.onset.cancer HR [95% CI];P value** | **New.onset.lung.cancer HR [95% CI];P value** | **New.onset.gastrointestinal.cancer HR [95% CI];P value** | **New.onset.breast.cancer HR [95% CI];P value** | **New.onset.bladder.cancer HR [95% CI];P value** | **New.onset.genitourinary.cancer HR [95% CI];P value** |
| SGLT2I v.s. DPP4I.1 | 0.80[0.70-0.90];0.0004*** | 0.54[0.41-0.71];<0.0001*** | 0.56[0.48-0.65];<0.0001*** | 0.51[0.35-0.74];0.0004*** | 0.65[0.52-0.82];0.0002*** | 0.46[0.30-0.70];0.0003*** | 0.28[0.14-0.55];0.0002*** | 0.47[0.32-0.69];0.0001*** |
| Dapagliflozin | 0.82[0.71-0.94];0.0060** | 0.60[0.43-0.85];0.0034** | 0.61[0.50-0.73];<0.0001*** | 0.64[0.42-0.99];0.0467* | 0.61[0.46-0.80];0.0003*** | 0.42[0.24-0.73];0.0021** | 0.40[0.18-0.88];0.0230* | 0.70[0.46-1.07];0.1000 |
| Empagliflozin | 0.73[0.58-0.93];0.0095** | 0.52[0.29-0.93];0.0277* | 0.73[0.55-0.97];0.0312* | 0.59[0.29-1.22];0.1543 | 0.84[0.58-1.24];0.3839 | 0.64[0.30-1.38];0.2567 | 0.55[0.17-1.77];0.3151 | 0.69[0.35-1.36];0.2879 |
| Canagliflozin | 0.94[0.78-1.15];0.5612 | 0.82[0.53-1.27];0.3697 | 0.71[0.55-0.92];0.0102* | 0.48[0.24-0.99];0.0475* | 0.90[0.64-1.27];0.5540 | 1.04[0.58-1.86];0.8990 | 0.14[0.02-1.04];0.0545 | 0.30[0.12-0.74];0.0089** |
| Ertugliflozin | 1.09[0.86-1.38];0.4830 | 0.81[0.45-1.45];0.4714 | 0.58[0.40-0.84];0.0045** | 0.56[0.23-1.38];0.2077 | 0.78[0.49-1.26];0.3154 | 0.41[0.13-1.29];0.1279 | 0.27[0.04-1.98];0.1989 | 0.34[0.11-1.07];0.0651 |

**Supplementary Table 6. Annualized incidence rate (IR) per 1000 person-year of primary and secondary cancer outcomes, all-cause mortality and cancer related mortality in the cohort before and after 1:1 propensity score matching of SGLT2I and DPP4 cohort**

| **Before matching** | | | | **After 1:1 propensity score matching** | | |
| --- | --- | --- | --- | --- | --- | --- |
| **All-cause mortality** | | | | | | |
| Overall | Person-year | Events | IR [95% CI] | Person-year | Events | IR [95% CI] |
|  | 3.29 x 10^5 | 3033 | 9.2 [8.6-9.6] | 2.01 x 10^5 | 970 | 4.8 [4.5-5.1] |
| DPP4I |  | | | | | |
| (0 - 1] | 41932.4 | 36 | 0.9 [0.6-1.2] | 18165.6 | 4 | 0.2 [0.1-0.6] |
| (1 - 2] | 41808.1 | 237 | 5.7 [5-6.4] | 18155.9 | 23 | 1.3 [0.8-1.9] |
| (2 - 3] | 41438.9 | 546 | 13.2 [12.1-14.3] | 18103.5 | 104 | 5.7 [4.7-7] |
| (3 - 4] | 40735 | 780 | 19.1 [17.9-20.5] | 17948.4 | 169 | 9.4 [8.1-10.9] |
| (4 - 5] | 39955.4 | 747 | 18.7 [17.4-20.1] | 17762.8 | 192 | 10.8 [9.4-12.5] |
| (5 - 6] | 22471.1 | 195 | 8.7 [7.5-10] | 10243.2 | 47 | 4.6 [3.4-6.1] |
| > 6 | 0.1 | 0 | 0 | 0 | 0 | 0 |
| SGLT2I |  | | | | | |
| (0 - 1] | 18167 | 0 | 0 | 18167 | 0 | 0 |
| (1 - 2] | 18157.7 | 28 | 1.5 [1.1-2.2] | 18159 | 24 | 1.3 [0.9-2] |
| (2 - 3] | 18111.5 | 66 | 3.6 [2.9-4.6] | 18119.3 | 57 | 3.1 [2.4-4.1] |
| (3 - 4] | 18017.5 | 129 | 7.2 [6-8.5] | 18037 | 114 | 6.3 [5.3-7.6] |
| (4 - 5] | 17841.9 | 209 | 11.7 [10.2-13.4] | 17882.2 | 181 | 10.1 [8.7-11.7] |
| (5 - 6] | 10117.4 | 60 | 5.9 [4.6-7.6] | 10154.9 | 55 | 5.4 [4.2-7.1] |
| >6 | 0.1 | 0 | 0 | 0 | 0 | 0 |
| **Cancer-caused mortality** | | | | | | |
| Overall | Person-year | Events | IR [95% CI] | Person-year | Events | IR [95% CI] |
|  | 3.29 x 10^5 | 506 | 1.5 [1.4-1.7] | 2.01 x 10^5 | 211 | 1.1 [0.9-1.2] |
| DPP4I |  |  |  |  |  |  |
| (0 - 1] | 41932.4 | 1 | 0 [0-0.2] | 18165.6 | 0 | 0 |
| (1 - 2] | 41808.1 | 35 | 0.8 [0.6-1.2] | 18155.9 | 7 | 0.4 [0.2-0.8] |
| (2 - 3] | 41438.9 | 93 | 2.2 [1.8-2.8] | 18103.5 | 25 | 1.4 [0.9-2] |
| (3 - 4] | 40735 | 136 | 3.3 [2.8-3.9] | 17948.4 | 58 | 3.2 [2.5-4.2] |
| (4 - 5] | 39955.4 | 110 | 2.8 [2.3-3.3] | 17762.8 | 41 | 2.3 [1.7-4.2] |
| (5 - 6] | 22471.1 | 16 | 0.7 [0.4-1.2] | 10243.2 | 6 | 0.6 [0.3-1.3] |
| > 6 | 0.1 | 0 | 0 | 0 | 0 | 0 |
| SGLT2I |  |  |  |  |  |  |
| (0 - 1] | 18167 | 0 | 0 | 18167 | 0 | 0 |
| (1 - 2] | 18157.7 | 4 | 0.2 [0.1-0.6] | 18159 | 2 | 0.1 [0-0.4] |
| (2 - 3] | 18111.5 | 16 | 0.9 [0.5-1.4] | 18119.3 | 9 | 0.5 [0.3-1] |
| (3 - 4] | 18017.5 | 33 | 1.8 [1.3-2.6] | 18037 | 22 | 1.2 [0.8-1.9] |
| (4 - 5] | 17841.9 | 51 | 2.9 [2.2-3.8] | 17882.2 | 33 | 1.8 [1.3-2.6] |
| (5 - 6] | 10117.4 | 11 | 1.1 [0.6-2] | 10154.9 | 8 | 0.8 [0.4-1.6] |
| >6 | 0.1 | 0 | 0 | 0 | 0 | 0 |
| **New-onset all-cause cancer** | | | | | | |
| Overall | Person-year | Events | IR [95% CI] | Person-year | Events | IR [95% CI] |
|  | 3.26 x 10^5 | 1533 | 4.7 [4.5-4.9] | 2.00 x 10^5 | 674 | 3.4 [3.1-3.6] |
| DPP4I |  |  |  |  |  |  |
| (0 - 1] | 41932.4 | 0 | 0 | 18165.6 | 0 | 0 |
| (1 - 2] | 41681.8 | 287 | 6.9 [6.1-7.7] | 18119 | 81 | 4.5 [3.6-5.6] |
| (2 - 3] | 41061.1 | 360 | 8.8 [7.9-9.7] | 17969.6 | 162 | 9 [7.7-10.5] |
| (3 - 4] | 40110 | 393 | 9.8 [9.9-10.8] | 17709.5 | 137 | 7.7 [6.5-9.1] |
| (4 - 5] | 39149.9 | 190 | 4.9 [4.2-5.6] | 17462.4 | 52 | 3 [2.3-3.9] |
| (5 - 6] | 22013.3 | 0 | 0 | 10082.4 | 0 | 0 |
| > 6 | 0.1 | 0 | 0 | 0 | 0 | 0 |
| SGLT2I |  | | | | | |
| (0 - 1] | 18167 | 0 | 0 | 18167 | 0 | 0 |
| (1 - 2] | 18129.9 | 54 | 3 [2.3-3.9] | 18132.7 | 45 | 2.5 [1.9-3.3] |
| (2 - 3] | 18030.1 | 86 | 4.8 [3.9-5.9] | 18053.8 | 63 | 3.5 [2.7-4.5] |
| (3 - 4] | 17852.3 | 103 | 5.8 [4.8-7] | 17903.8 | 85 | 4.7 [3.8-5.9] |
| (4 - 5] | 17623.7 | 60 | 3.4 [2.6-4.4] | 17700.4 | 49 | 2.8 [2.1-3.7] |
| (5 - 6] | 9985.7 | 0 | 0 | 10043.6 | 0 | 0 |
| > 6 | 0.1 | 0 | 0 | 0 | 0 | 0 |
| **New-onset lung cancer** | | | | | | |
| Overall | Person-year | Events | IR [95% CI] | Person-year | Events | IR [95% CI] |
|  | 3.28 x 10^5 | 249 | 0.8 [0.7-0.9] | 2.01 x 10^5 | 124 | 0.6 [0.5-0.7] |
| DPP4I |  |  |  |  |  |  |
| (0 - 1] | 41932.4 | 0 | 0 | 18165.6 | 0 | 0 |
| (1 - 2] | 41795.7 | 38 | 0.9 [0.7-1.2] | 18148.1 | 17 | 0.9 [0.6-1.5] |
| (2 - 3] | 41400.5 | 55 | 1.3 [1-1.7] | 18083.3 | 29 | 1.6 [1.1-2.3] |
| (3 - 4] | 40658.9 | 72 | 1.8 [1.4-2.2] | 17913.7 | 26 | 1.5 [1-2.1] |
| (4 - 5] | 39856.7 | 36 | 0.9 [0.7-1.3] | 17725.2 | 10 | 0.6 [0.3-1] |
| (5 - 6] | 22418.3 | 0 | 0 | 10224.4 | 0 | 0 |
| > 6 | 0.1 | 0 | 0 | 0 | 0 | 0 |
| SGLT2I |  |  |  |  |  |  |
| (0 - 1] | 18167 | 0 | 0 | 18167 | 0 | 0 |
| (1 - 2] | 18152.4 | 12 | 0.7 [0.4-1.2] | 18153.8 | 11 | 0.6 [0.3-1.1] |
| (2 - 3] | 18097.3 | 11 | 0.6 [0.3-1.1] | 18106.4 | 9 | 0.5 [0.3-1] |
| (3 - 4] | 17991.7 | 15 | 0.8 [0.5-1.4] | 18014 | 12 | 0.7 [0.4-1.2] |
| (4 - 5] | 17810.8 | 10 | 0.6 [0.3-1] | 17852.1 | 10 | 0.6 [0.3-1] |
| (5 - 6] | 10097.8 | 0 | 0 | 10135.3 | 0 | 0 |
| > 6 | 0.1 | 0 | 0 | 0 | 0 | 0 |
| **New-onset gastrointestinal cancer** | | | | | | |
| Overall | Person-year | Events | IR [95% CI] | Person-year | Events | IR [95% CI] |
|  | 3.27 x 10^5 | 817 | 2.5 [2.3-2.7] | 2.00 x 10^5 | 325 | 1.6 [1.5-1.8] |
| DPP4I |  |  |  |  |  |  |
| (0 - 1] | 41932.4 | 0 | 0 | 18165.6 | 0 | 0 |
| (1 - 2] | 41741.4 | 158 | 3.8 [3.2-4.4] | 18135.7 | 43 | 2.4 [1.8-3.2] |
| (2 - 3] | 41234 | 202 | 4.9 [4.3-5.6] | 18038.7 | 72 | 4 [3.2-5] |
| (3 - 4] | 40401.7 | 206 | 5.1 [4.4-5.8] | 17832.4 | 65 | 3.6 [2.9-4.6] |
| (4 - 5] | 39537 | 100 | 2.5 [2.1-3.1] | 17631.9 | 16 | 0.9 [0.6-1.5] |
| (5 - 6] | 22236.4 | 0 | 0 | 10176.1 | 0 | 0 |
| > 6 | 0.1 | 0 | 0 | 0 | 0 | 0 |
| SGLT2I |  |  |  |  |  |  |
| (0 - 1] | 18167 | 0 | 0 | 18167 | 0 | 0 |
| (1 - 2] | 18148.6 | 20 | 1.1 [0.7-1.7] | 18150.6 | 16 | 0.9 [0.5-1.4] |
| (2 - 3] | 18075.8 | 49 | 2.7 [2-3.6] | 18090.9 | 39 | 2.2 [1.6-3] |
| (3 - 4] | 17939.5 | 54 | 3 [2.3-3.9] | 17970.5 | 50 | 2.8 [2.1-3.7] |
| (4 - 5] | 17742.7 | 28 | 1.6 [1.1-2.3] | 17792.1 | 24 | 1.3 [0.9-2] |
| (5 - 6] | 10057.2 | 0 | 0 | 10098.4 | 0 | 0 |
| > 6 | 0.1 | 0 | 0 | 0 | 0 | 0 |
| **New-onset bladder cancer** | | | | | | |
| Overall | Person-year | Events | IR [95% CI] | Person-year | Events | IR [95% CI] |
|  | 3.29 x 10^5 | 97 | 0.3 [0.2-0.4] | 2.01 x 10^5 | 50 | 0.2 [0.2-0.3] |
| DPP4I |  |  |  |  |  |  |
| (0 - 1] | 41932.4 | 0 | 0 | 18165.6 | 0 | 0 |
| (1 - 2] | 41801.8 | 15 | 0.4 [0.2-0.6] | 18154.1 | 4 | 0.2 [0.1-0.6] |
| (2 - 3] | 41417.1 | 20 | 0.5 [0.3-0.7] | 18089.6 | 24 | 1.3 [0.9-2] |
| (3 - 4] | 40698.4 | 24 | 0.6 [0.4-0.9] | 17916.8 | 9 | 0.5 [0.3-1] |
| (4 - 5] | 39901.6 | 14 | 0.4 [0.2-0.6] | 17726.1 | 2 | 0.1 [0-0.5] |
| (5 - 6] | 22440.3 | 0 | 0 | 10223 | 0 | 0 |
| > 6 | 0.1 | 0 | 0 | 0 | 0 | 0 |
| SGLT2I |  |  |  |  |  |  |
| (0 - 1] | 18167 | 0 | 0 | 18167 | 0 | 0 |
| (1 - 2] | 18156 | 5 | 0.3 [0.1-0.7] | 18157.6 | 3 | 0.2 [0.1-0.5] |
| (2 - 3] | 18104.2 | 7 | 0.4 [0.2-0.8] | 18115.7 | 3 | 0.2 [0.1-0.5] |
| (3 - 4] | 18000.7 | 8 | 0.4 [0.2-0.9] | 18029.2 | 3 | 0.2 [0.1-0.5] |
| (4 - 5] | 17819.2 | 4 | 0.2 [0.1-0.6] | 17872.2 | 2 | 0.1 [0-0.4] |
| (5 - 6] | 10102.7 | 0 | 0 | 10148 | 0 | 0 |
| > 6 | 0.1 | 0 | 0 | 0 | 0 | 0 |
| **New-onset genitourinary cancer** | | | | | | |
| Overall | Person-year | Events | IR [95% CI] | Person-year | Events | IR [95% CI] |
|  | 3.28 x 10^5 | 261 | 0.8 [0.7-0.9] | 2.01 x 10^5 | 121 | 0.6 [0.5-0.7] |
| DPP4I |  |  |  |  |  |  |
| (0 - 1] | 41932.4 | 0 | 0 | 18165.6 | 0 | 0 |
| (1 - 2] | 41781.6 | 50 | 1.2 [0.9-1.6] | 18149.1 | 12 | 0.7 [0.4-1.2] |
| (2 - 3] | 41364.8 | 65 | 1.6 [1.2-2] | 18065.8 | 49 | 2.7 [2-3.6] |
| (3 - 4] | 40618.9 | 60 | 1.5 [1.1-1.9] | 17884.1 | 17 | 1 [0.6-1.5] |
| (4 - 5] | 39803.2 | 29 | 0.7 [0.5-1] | 17689.3 | 4 | 0.2 [0.1-0.6] |
| (5 - 6] | 22384.7 | 0 | 0 | 10203.1 | 0 | 0 |
| > 6 | 0.1 | 0 | 0 | 0 | 0 | 0 |
| SGLT2I |  |  |  |  |  |  |
| (0 - 1] | 18167 | 0 | 0 | 18167 | 0 | 0 |
| (1 - 2] | 18151.9 | 12 | 0.7 [0.4-1.2] | 18153.5 | 10 | 0.6 [0.3-1] |
| (2 - 3] | 18093.7 | 15 | 0.8 [0.5-1.4] | 18106.5 | 9 | 0.5 [0.3-0.1] |
| (3 - 4] | 17982.3 | 16 | 0.9 [0.5-1.5] | 18013.1 | 10 | 0.6 [0.3-1] |
| (4 - 5] | 17792.4 | 14 | 0.8 [0.5-1.3] | 17849.8 | 10 | 0.6 [0.3-1] |
| (5 - 6] | 10084.1 | 0 | 0 | 10132.5 | 0 | 0 |
| > 6 | 0.1 | 0 | 0 | 0 | 0 | 0 |
| **New-onset breast cancer** | | | | | | |
| Overall | Person-year | Events | IR [95% CI] | Person-year | Events | IR [95% CI] |
|  | 3.28 x 10^5 | 201 | 0.6 [0.5-0.7] | 2.01 x 10^5 | 101 | 0.5 [0.4-0.6] |
| DPP4I |  |  |  |  |  |  |
| (0 - 1] | 41932.4 | 0 | 0 | 18165.6 | 0 | 0 |
| (1 - 2] | 41787.2 | 41 | 1 [0.7-1.3] | 18153.8 | 9 | 0.5 [0.3-1] |
| (2 - 3] | 41376.2 | 40 | 1 [0.7-1.3] | 18091.5 | 13 | 0.7 [0.4-1.2] |
| (3 - 4] | 40634.7 | 50 | 1.2 [0.9-1.6] | 17923.2 | 25 | 1.4 [0.9-2.1] |
| (4 - 5] | 39816.1 | 26 | 0.7 [0.4-1] | 17703.5 | 22 | 1.2 [0.8-1.9] |
| (5 - 6] | 22386.6 | 0 | 0 | 10207.1 | 0 | 0 |
| > 6 | 0.1 | 0 | 0 | 0 | 0 | 0 |
| SGLT2I |  |  |  |  |  |  |
| (0 - 1] | 18167 | 0 | 0 | 18167 | 0 | 0 |
| (1 - 2] | 18150.9 | 10 | 0.6 [0.3-1] | 18152.6 | 8 | 0.4 [0.2-0.9] |
| (2 - 3] | 18097.9 | 10 | 0.6 [0.3-1] | 18108 | 6 | 0.3 [0.1-0.7] |
| (3 - 4] | 17992.1 | 15 | 0.8 [0.5-1.4] | 18017.9 | 11 | 0.6 [0.3-1.1] |
| (4 - 5] | 17803.6 | 9 | 0.5 [0.3-1] | 17853.1 | 7 | 0.4 [0.2-0.8] |
| (5 - 6] | 10097.6 | 0 | 0 | 10140.9 | 0 | 0 |
| > 6 | 0.1 | 0 | 0 | 0 | 0 | 0 |
